# A Systematic Evaluation of Machine Learning-based Biomarkers for Major Depressive Disorder across Modalities

**DOI:** 10.1101/2023.02.27.23286311

**Authors:** Nils R. Winter, Julian Blanke, Ramona Leenings, Jan Ernsting, Lukas Fisch, Kelvin Sarink, Carlotta Barkhau, Katharina Thiel, Kira Flinkenflügel, Alexandra Winter, Janik Goltermann, Susanne Meinert, Katharina Dohm, Jonathan Repple, Marius Gruber, Elisabeth J. Leehr, Nils Opel, Dominik Grotegerd, Ronny Redlich, Robert Nitsch, Jochen Bauer, Walter Heindel, Joachim Groß, Till F. M. Andlauer, Andreas J. Forstner, Markus M. Nöthen, Marcella Rietschel, Stefan G. Hofmann, Julia-Katharina Pfarr, Lea Teutenberg, Paula Usemann, Florian Thomas-Odenthal, Adrian Wroblewski, Katharina Brosch, Frederike Stein, Andreas Jansen, Hamidreza Jamalabadi, Nina Alexander, Benjamin Straube, Igor Nenadić, Tilo Kircher, Udo Dannlowski, Tim Hahn

## Abstract

**Background:** Biological psychiatry aims to understand mental disorders in terms of altered neurobiological pathways. However, for one of the most prevalent and disabling mental disorders, Major Depressive Disorder (MDD), patients only marginally differ from healthy individuals on the group-level. Whether Precision Psychiatry can solve this discrepancy and provide specific, reliable biomarkers remains unclear as current Machine Learning (ML) studies suffer from shortcomings pertaining to methods and data, which lead to substantial over-as well as underestimation of true model accuracy.

**Methods:** Addressing these issues, we quantify classification accuracy on a single-subject level in N=1,801 patients with MDD and healthy controls employing an extensive multivariate approach across a comprehensive range of neuroimaging modalities in a well-curated cohort, including structural and functional Magnetic Resonance Imaging, Diffusion Tensor Imaging as well as a polygenic risk score for depression.

**Findings:** Training and testing a total of 2.4 million ML models, we find accuracies for diagnostic classification between 48.1% and 62.0%. Multimodal data integration of all neuroimaging modalities does not improve model performance. Similarly, training ML models on individuals stratified based on age, sex, or remission status does not lead to better classification. Even under simulated conditions of perfect reliability, performance does not substantially improve. Importantly, model error analysis identifies symptom severity as one potential target for MDD subgroup identification.

**Interpretation:** Although multivariate neuroimaging markers increase predictive power compared to univariate analyses, single-subject classification – even under conditions of extensive, best-practice Machine Learning optimization in a large, harmonized sample of patients diagnosed using state-of-the-art clinical assessments – does not reach clinically relevant performance. Based on this evidence, we sketch a course of action for Precision Psychiatry and future MDD biomarker research.

**Funding:** The German Research Foundation, and the Interdisciplinary Centre for Clinical Research of the University of Münster.

## Introduction

Overcoming Cartesian mind-body dualism was the pivotal achievement of biological psychiatry in the 20th century, enabling the treatment of mental disorders as disorders of the brain.[1] Since the effectiveness of physical interventions such as neuropsychopharmacological treatments as well as the substantial heritability of many psychiatric disorders in principle support this dogma, hopes are high for biomarkers to inform diagnosis and treatment. However, identifying specific, reliable neurobiological deviations informative on the level of the individual patient has proven elusive even after decades of intense research, with the clinical reality of patients remaining largely unchanged.[2, 3] For Major Depressive Disorder (MDD) mounting evidence suggests that group-level, univariate neuroimaging or genetic markers only marginally differ between healthy controls and patients with MDD, with the distributions of patients and controls overlapping more than 85% even under optimal conditions.[4–6] Fuelled by the availability of large-scale datasets as well as substantial improvements regarding Machine Learning (ML) software and hardware, the field of Precision Psychiatry has gained increasing traction over the last decade. Precision Psychiatry aims to build models which allow for individual predictions, thereby moving from the investigation of univariate statistical group differences towards multivariate neurobiological patterns of individual patients. This focus on prediction and prognosis instead of group-level inference as well as the ability for a direct assessment of clinical utility renders Precision Psychiatry essential in all translational efforts.[7–11] While a consensus on best-practice guidelines for Precision Psychiatry and ML has been emerging[7, 10, 11], four broad issues in MDD biomarker research remain which may lead to substantial over-as well as underestimation of the true predictive performance: First, methodological shortcomings in predictive model validation (e.g. data leakage between training and test set) lead to an overestimation of predictive performance in many publications.[12] Strikingly, about a quarter of all published studies using predictive models in psychiatry do not provide any kind of model validation and thus do not provide any information regarding predictive performance in new patients.[13] In the same vein, small sample sizes for model evaluation, such as those most common in the literature today, often result in unreliable and eventually inflated estimates of predictive performance.[14] Second, many published studies rely on a single ML algorithm; often without optimizing model performance through hyperparameter tuning, thereby running the risk of greatly underestimating true predictive performance.[15] Third, current studies almost exclusively focus on a single data modality and studies integrating multiple modalities to increase predictive performance are rare.[7, 15] Fourth, clinical assessment of MDD diagnosis across studies is inconsistent and especially for larger studies often relies on self-report questionnaires rather than clinical interviews by a trained clinician, thus rendering diagnostic labels more heterogeneous and less reliable.[16, 17] Similarly, a lack of harmonization of study protocols, resulting in clinical heterogeneity of patient samples and recruitment modalities, quality control, and neuroimaging data acquisition in multi-site analyses has previously been used to explain small effect sizes and inconsistent results.[18] In summary, the existing literature on multivariate biomarker discovery in MDD does not allow for a conclusive evaluation of clinical utility of ML approaches. Here, we explicitly address these previous shortcomings to systematically evaluate ML-based multivariate biomarkers for MDD across neuroimaging modalities: We performed nested cross-validation to separate the model optimization step from the estimation of general-izability and ensured adequate test sets by using one of the largest single-study MDD cohorts for which multimodal data and in-depth diagnostic assessment is available (N=1,801 MDD patients and controls).[19, 20] Next, we did not rely on a single predictive algorithm, but capitalized on the advances in ML software[21, 22] and computational capabilities to combine multiple classifiers from complementary algorithmic categories including feature selection, dimensionality reduction, and extensive tuning of model hyperparameters, resulting in a total of 2.4 million machine learning models trained and evaluated in this study. Expanding previous work, we drew upon a comprehensive set of neuroimaging modalities including structural Magnetic Resonance Imaging (MRI), task-based and resting-state functional MRI (fMRI), Diffusion Tensor Imaging (DTI) as well as an MDD polygenic risk score and several environmental risk factors. This allowed us to directly compare predictive performance across modalities in the same sample and enabled us to quantify the potential benefit of multimodal data integration. In addition, clinical assessment of patients in our data was based on structured clinical interviews (SCID) which provided standardized DSM-based MDD diagnosis and therefore reduced the diagnostic uncertainty often hampering model performance in large-scale, multi-site data today. Likewise, methodological heterogeneity due to, e.g. differing exclusion criteria, recruitment modalities, clinical phenotyping, or MRI scanning protocols, could be alleviated in this well-curated, harmonized sample.[20] Finally, the low reliability of neuroimaging data and psychiatric diagnosis is being discussed as one of the major drivers for small effect sizes currently reported in the literature.[17, 23–26] To address this hypothesis, we systematically simulated classification performance in scenarios of optimal reliability and quantified expected improvements. Considering the substantial heterogeneity of patients with MDD, we finally conducted in-depth analyses of model errors to uncover characteristics of patients that contribute to misclassification, thereby shedding light on subgroups for which neuroimaging-based predictive models are successful or might fail.[27–29]

## Methods

### Study design and participants

The data used in this study are part of the Marburg-Münster Affective Disorders Cohort Study (MACS).[19, 20] Data were collected at two sites (Marburg and Münster, Germany) using identical study protocols and harmonized scanner settings.[20] The study was approved by the ethics committees of the medical faculties of the University of Marburg, Germany, and the University of Münster, Germany. Participants received financial compensation and gave written and informed consent. At the time of data analysis, a sample of N=2,036 healthy participants and patients with major depression were recruited as part of the MACS cohort (*eMethods 1-3*). Clinical diagnosis was assessed using the Structured Clinical Interview for DSM-IV, axis 1 disorders (SCID-I).[30] Patients were recruited from local in- and outpatient services and either fulfilled the DSM-IV criteria for an acute major depressive episode or had a lifetime history of a major depressive episode. Individuals with any history of neurological or medical conditions were excluded, resulting in a final sample of N=1,801. See *eMethods 1* for further information on exclusion criteria. Participants were recruited from September 11, 2014, to September 26, 2018. For every neuroimaging data modality, all participants for whom data of the specific modality were available and passed quality checks were used in subsequent analyses (see *eMethods 1 and 4-12*). This study followed Strengthening the Reporting of Observational Studies in Epidemiology (STROBE) reporting guidelines.[31]

### Procedures and neuroimaging data modalities

The neuroimaging, genetic and behavioural data used in this study have been described previously.[5] Detailed information is available in *eMethods 4-12*. In short, voxel-based morphometry (VBM, CAT12 toolbox) and region-based surface, thickness and volume (FreeSurfer) were extracted from T_1_-weighted structural MRI.[32, 33] Structural connectomes were derived from DTI as fractional anisotropy (FA) and mean diffusivity (MD).[34] Functional connectomes were derived from resting-state functional MRI (rsfMRI). Voxel-based local correlation (LCOR), the amplitude of low-frequency fluctuations (ALFF) as well as the fractional amplitude of low-frequency fluctuations (fALFF) were also computed from rsfMRI.[35] For both structural and functional connectomes commonly used graph network parameters such as betweenness centrality, degree centrality, or global efficiency were calculated.[36] Task-based fMRI was based on an established emotional face matching paradigm and a faces versus shapes contrast was used.[37, 38] In addition, we compared results to a commonly used polygenic risk score for depression (PRS, *eMethods 5*)[39, 40] as well as questionnaire data on adverse experiences during childhood (Childhood Trauma Questionnaire; CTQ) and current social support (F-SozU), since these variables are established risk or protective factors in the aetiology of major depression.[39, 41, 42] A medication load index was calculated expressing the current psychiatric medication. Current depressive symptoms were assessed using the Beck Depression Inventory (BDI) and Hamilton Depression Rating Scale (HAMD).[43, 44]

### Choice of the primary measures

Accuracy of predicted diagnostic labels in all machine learning models was calculated using the widely used balanced classification accuracy (BACC), sensitivity, specificity, and area under the receiver operating characteristic curve (AUC), following STAR*D guidelines for reporting predictive accuracy. In addition, we calculated Matthew’s correlation coefficient (MCC, Equation 1). For all metrics, mean and standard deviation across the 10 outer cross-validation splits were reported to assess the generalizability of the predictive models.

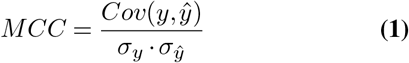

### Machine Learning analyses

A total of 2.4 million machine learning models to classify healthy participants and patients with MDD were trained, optimized and evaluated (see Figure 1, *eMethods 14*). A single ML pipeline consisted of a sequence of data transformation steps and a final classification algorithm. Data transformations included an imputation of missing data, a feature normalization, selection of a percentage of univariate features with the highest effect size, and a principal component analysis (PCA) to reduce the dimensionality of the brain data. Subsequently, a classification algorithm was trained to predict diagnosis, including support vector machines, random forests, logistic regression, k-nearest neighbour, Gaussian naive Bayes, and boosting classifiers. A nested cross-validation scheme with 10 inner validation and 10 outer test splits was used to optimize hyper-parameters and assess final generalizability. These primary ML analyses were complemented by analyses for subgroups of acutely depressed patients (omitting remitted patients) or recurrently depressed patients (omitting single episode patients), males and females, as well as a homogeneous age group (age range 24 to 28). For more details see *eMethods 3*.

**Fig. 1.**
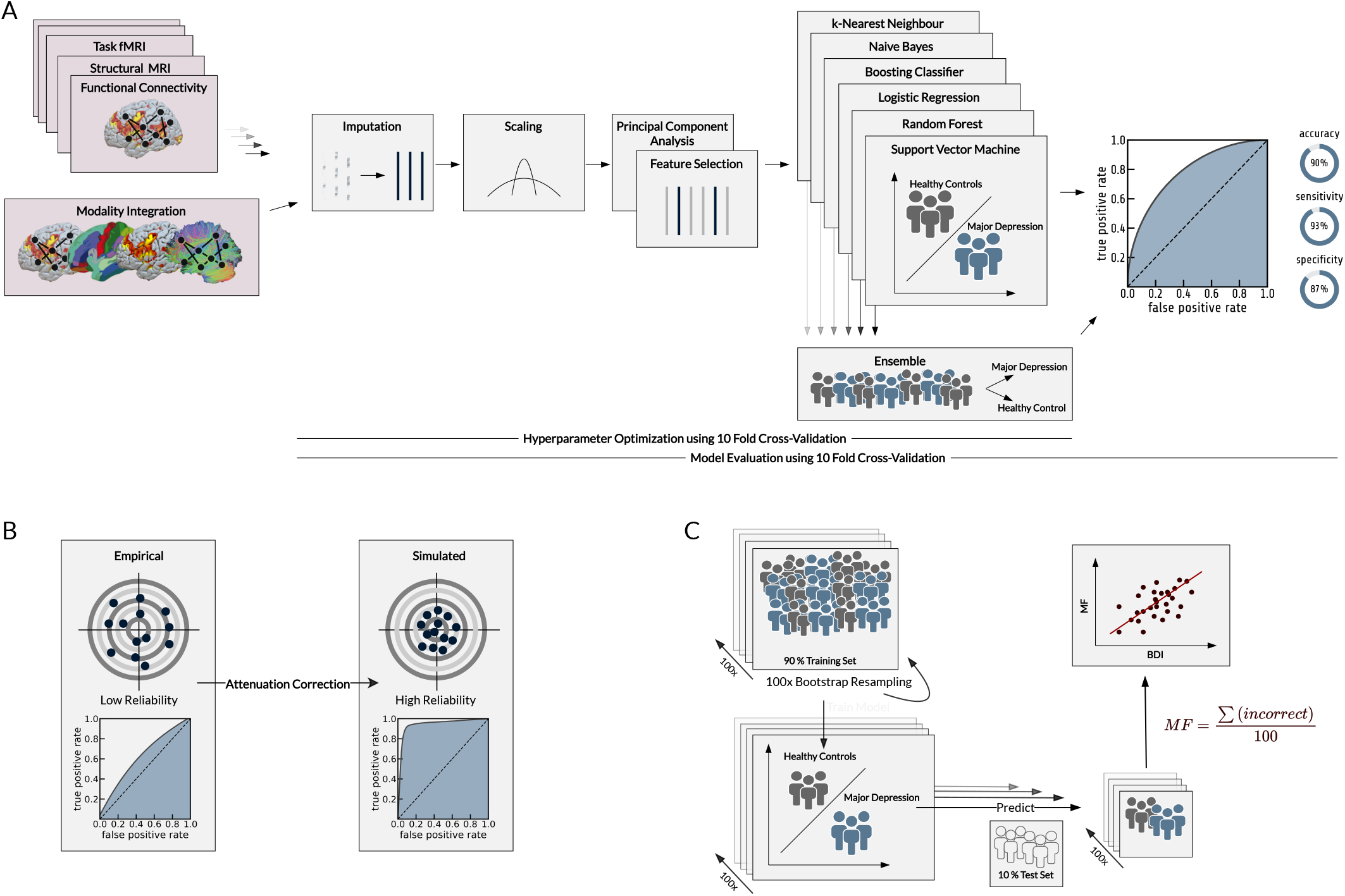
Overview of all analyses. (A) illustrates steps of the Machine Learning pipeline. (B) illustrates reliability correction and its effect on classification accuracy. (C) illustrates model error analysis using misclassification frequency (MF) through repeated bootstrapping.

### Modality integration

Brain modality integration was accomplished using two strategies. First, a PCA was performed for every data modality separately and the resulting components were then concatenated and used as input to the ML pipelines. Second, a voting ensemble strategy was used combining all diagnosis predictions from the unimodal models. Final predictions were calculated using a majority vote. All ML analyses were performed using PHOTONAI.[22] Scripts are available at https://github.com/wwu-mmll.

## Simulation of perfect reliability

To quantify the effect of reliability on classification performance, we performed exploratory analyses using attenuation correction from classical test theory to simulate the true classification accuracy occurring if the reliability of the data was perfect.[45] We first computed MCC from the model predictions *ŷ* and the actual diagnostic labels *y* (Equation 1).[46] This correlation was then corrected for an assumed reliability *ρ* using the attenuation formula (Equation 2).[47]

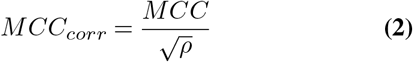

We conducted two separate attenuation correction analyses. First, we assume a reliability of *ρ*_*y*_ = 0.28 for an MDD diagnosis, which is based on the current literature on the interrater reliability of DSM-5 diagnoses.[17, 26] Second, we assumed reliabilities for neuroimaging data ranging from 0.1 to 1. The resulting corrected correlations were then converted back to BACC using prevalence *ϕ* and bias *β* with equations 15 and 21 in [46] (Equation 3, *eMethods 13*).

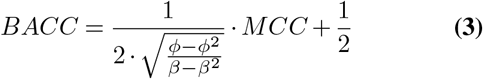

### Analysis of systematic model error

Identifying subgroups of individuals for whom brain-based ML models routinely fail has been shown to improve the development of generalizable predictive models.[27] In order to quantify this tendency for misclassification in every individual, we performed 100 bootstrap resampling runs on the training set of the best performing neuroimaging modality. One ML pipeline for every bootstrap training set was then trained and diagnostic labels for the participants in the test set were collected, resulting in 100 predictions (healthy, depressed) for every participant. The sum over incorrect classifications then leads to the frequency of misclassification (MF).[27] Finally, MF was correlated with external measures describing depressive symptom severity and demographic or environmental characteristics using Spearman rank correlation.

## Results

A total of 1,801 individuals (856 patients [47.5%] and 945 healthy controls [52.5%]) were included in the analyses (mean [SD] age, 36.1 [13.1] years; 555 female patients [64.8%] and 607 female healthy controls [64.2%], see Table 1 for details).

**Table 1.**
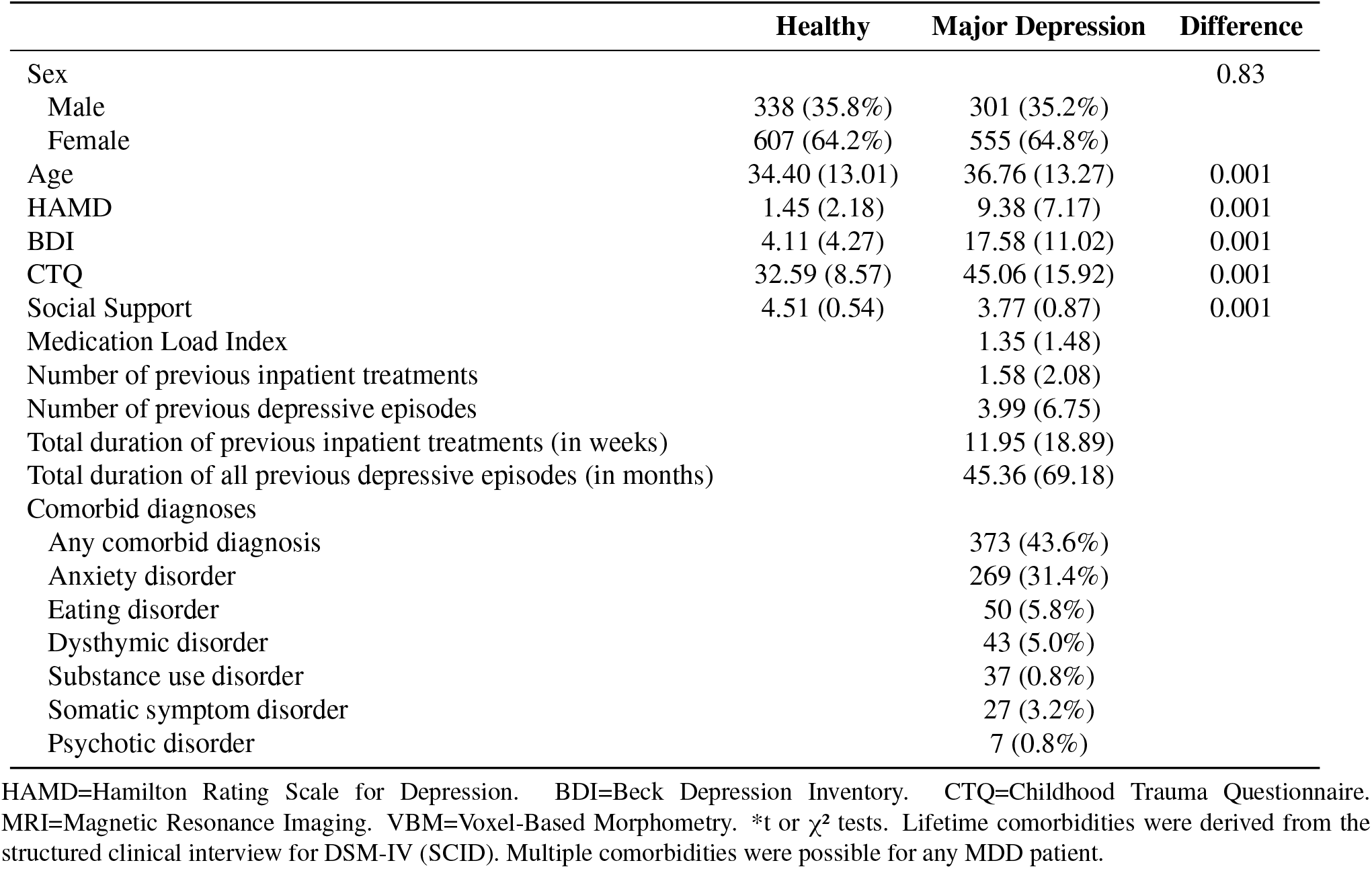
Socio-demographics and clinical characteristics of all participants.

|  | Healthy | Major Depression | Difference |
| --- | --- | --- | --- |
| Sex |  |  | 0.83 |
| Male | 338 (35.8%) | 301 (35.2%) |  |
| Female | 607 (64.2%) | 555 (64.8%) |  |
| Age | 34.40 (13.01) | 36.76 (13.27) | 0.001 |
| HAMD | 1.45 (2.18) | 9.38 (7.17) | 0.001 |
| BDI | 4.11 (4.27) | 17.58 (11.02) | 0.001 |
| CTQ | 32.59 (8.57) | 45.06 (15.92) | 0.001 |
| Social Support | 4.51 (0.54) | 3.77 (0.87) | 0.001 |
| Medication Load Index |  | 1.35 (1.48) |  |
| Number of previous inpatient treatments |  | 1.58 (2.08) |  |
| Number of previous depressive episodes |  | 3.99 (6.75) |  |
| Total duration of previous inpatient treatments (in weeks) |  | 11.95 (18.89) |  |
| Total duration of all previous depressive episodes (in months) |  | 45.36 (69.18) |  |
| Comorbid diagnoses |  |  |  |
| Any comorbid diagnosis |  | 373 (43.6%) |  |
| Anxiety disorder |  | 269 (31.4%) |  |
| Eating disorder |  | 50 (5.8%) |  |
| Dysthymic disorder |  | 43 (5.0%) |  |
| Substance use disorder |  | 37 (0.8%) |  |
| Somatic symptom disorder |  | 27 (3.2%) |  |
| Psychotic disorder |  | 7 (0.8%) |  |

HAMD=Hamilton Rating Scale for Depression. BDI=Beck Depression Inventory. CTQ=Childhood Trauma Questionnaire. MRI=Magnetic Resonance Imaging. VBM=Voxel-Based Morphometry. \*t or $\chi^2$ tests. Lifetime comorbidities were derived from the structured clinical interview for DSM-IV (SCID). Multiple comorbidities were possible for any MDD patient.

### Multivariate classification accuracy

Across neuroimaging modalities and ML algorithms, BACC ranged between 48.1% and 61.5% (see eTable 1-2 detailed results and *eMethods* for neuroimaging feature descriptions). Results for the single best ML algorithm in each modality are shown in Figure 2. Highest BACC was found for resting-state connectivity, with mean [SD] BACC ranging between 51.5% [7.1%] and 61.5% [3.4%]. Structural MRI as well as task-based fMRI showed lower BACC compared to all resting-state fMRI modalities. Calculating graph network parameters from DTI or resting-state fMRI did not improve overall BACC compared to using the functional or structural connectome directly. To investigate the effect of remission status and chronicity of the MDD population, we performed additional analyses limited to, first, MDD patients with acute symptoms (N=599) thus excluding remitted patients and, second, MDD patients with recurrent episodes (N=297). Overall, ML pipelines on subgroups did not outperform the analysis containing all MDD patients (BACC_*max*_=61.7%). Likewise, restricting analyses to male or female individuals or a more homogeneous age range of 24 to 28 did not change the overall results (BACC_*max*_=61.6%, see eFigure 1-5 and eTables 5-19).

**Fig. 2.**
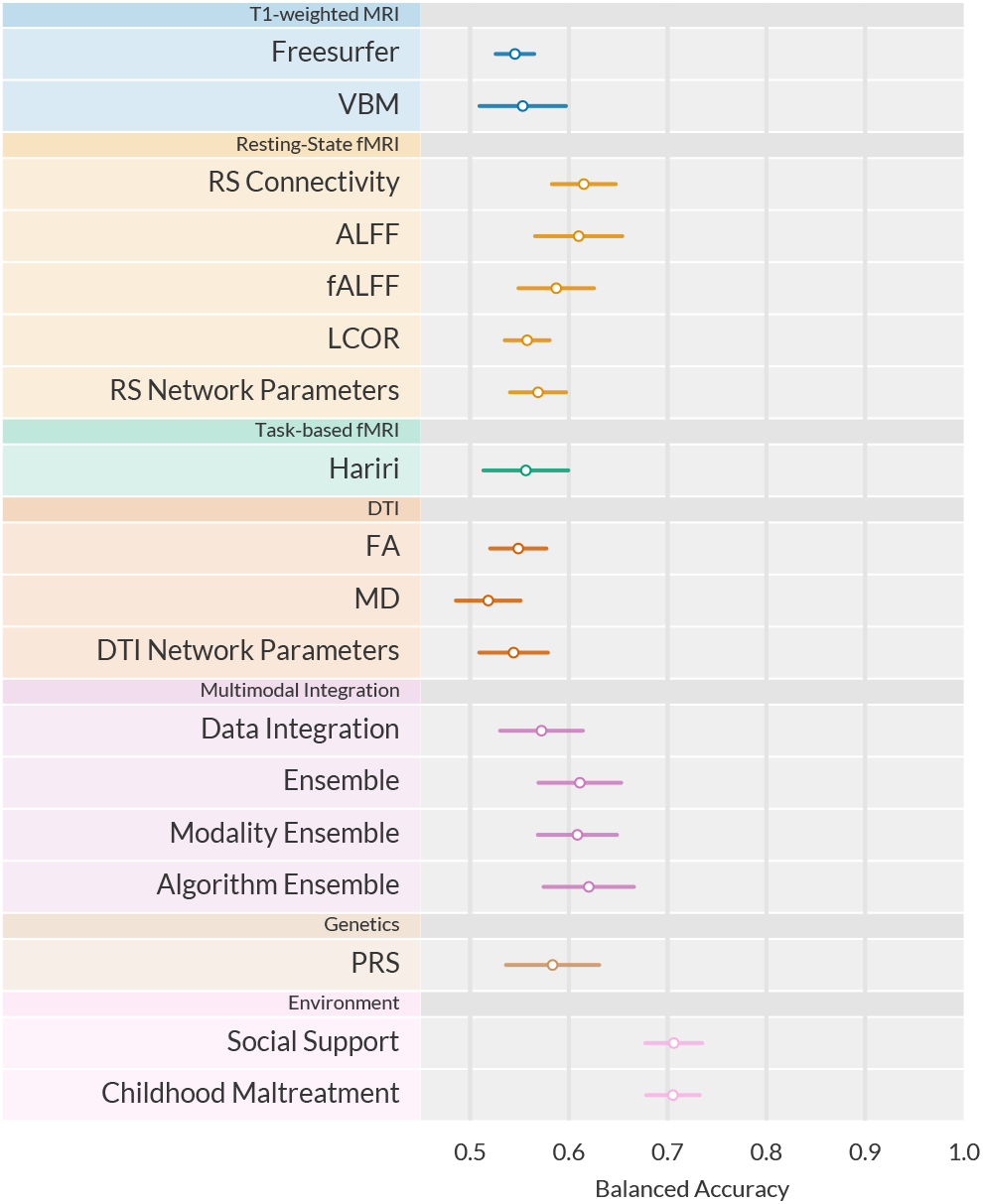
Balanced accuracy for best machine learning pipeline in every modality. Error bars display +-1 standard deviation calculated across the 10 outer cross-validation folds. VBM=Voxel-based morphometry, ALFF=Amplitude of low-frequency fluctuations, fALFF=fractional ALFF, LCOR=Local correlation, FA=Fractional anisotropy, MD=Mean diffusivity, PRS=Polygenic risk score.

### Multimodal Data Integration

Integration of neuroimaging modalities was evaluated using two alternative approaches. First, principal components from modality specific PCAs were concatenated and used as input to the previously described ML pipelines. This modality integration analysis achieved BACCs between 50.1% [4.0%] and 57.2% [4.4%] (eTable 3, Figure 2). Second, predicted labels from the unimodal models (across algorithm, across modalities, or across both) were combined into a majority-vote ensemble classifier. The voting ensemble classifier achieved a BACC of 61.1% [4.4%]. Both multimodal data integration methods did not improve the 61.5% accuracy reached in the best unimodal model. Combining predictions from all ALFF models achieved the highest BACC of 62.0% [4.8%].

### Comparison with Genetic and Environmental Variables

We next compared the neuroimaging-based ML models to the predictive performance of univariate approaches using genetic and environmental variables. While the Howard et al. depression PRS[39] achieved similar results to neuroimaging (BACC = 58.4% [5.0%]), both self-reported childhood maltreatment and social support outperformed brain-based and PRS-based models, achieving a BACC of 70.5% [2.9%] and 70.6% [3.0%], respectively.

### Effects of Reliability of Diagnosis and Neuroimaging Data

To investigate to what extent the reliability of neuroimaging data and diagnosis affect classification accuracy, we first converted BACC to MCC as a measure of the association between the actual and predicted diagnostic label. This correlation coefficient could then be corrected using the attenuation correction formula, estimating classification performance given perfect reliability. Second, we corrected for the lower bound of the MDD diagnosis reliability of *ρ* = 0.28 as reported in the literature (Figure 3A). With this approach, BACC for the best machine learning algorithm on resting-state connectivity increased to 71.8% [6.4%]. BACC for the voting ensemble increased to 73.4% [7.4%]. Third, we assumed reliability coefficients of neuroimaging modalities between 0.1 and 1 (Figure 3B). For the best unimodal analysis (resting-state connectivity), BACC increases to 66.3% for an assumed reliability of 0.5. These reliability correction analyses suggest that improving reliability might only have a minor positive effect on classification accuracy.

**Fig. 3.**
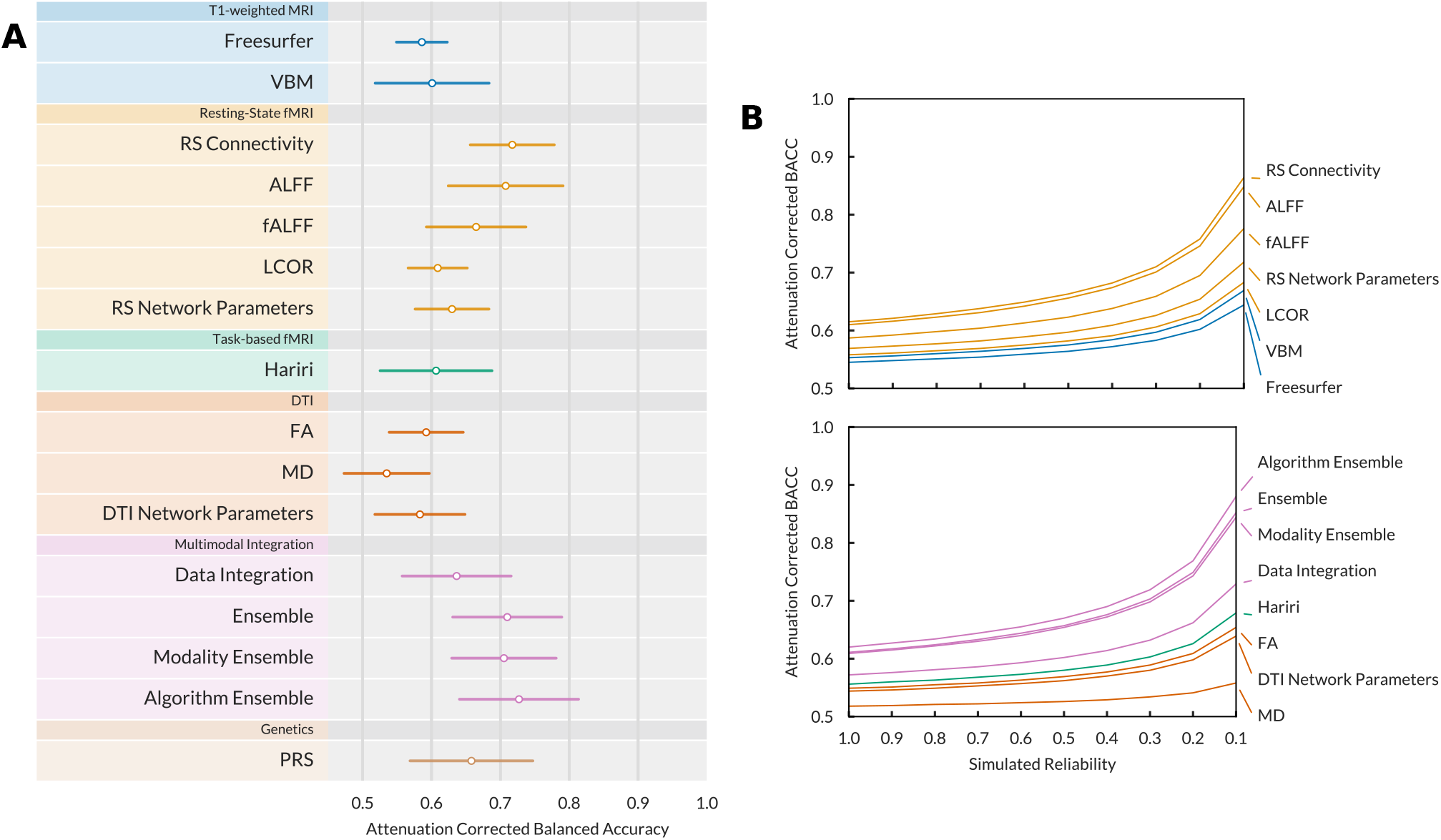
(A) Balanced accuracy for best machine learning pipeline in every modality after performing an attenuation correction for the empirical reliability of the MDD diagnosis. Error bars display +-1 standard deviation calculated across the 10 outer cross-validation folds. (B) Balanced accuracy for best machine learning pipeline in every modality after performing an attenuation correction for simulated reliability of the neuroimaging data. A simulated reliability of 1 corresponds to the empirical results achieved in the unimodal analyses. Decreasing the simulated reliability results in a corrected BACC. VBM=Voxel-based morphometry, ALFF=Amplitude of low-frequency fluctuations, fALFF=fractional ALFF, LCOR=Local correlation, FA=Fractional anisotropy, MD=Mean diffusivity.

### Analysis of Systematic Model Errors

Recent work has shown that brain-based predictive models do not work equally well for all individuals, potentially displaying substantial bias.[27] Therefore, identifying patients for which ML models repeatedly fail will be informative for the development of clinical useful predictive models. The frequency with which each individual was incorrectly classified as either healthy or depressed was measured using the misclassification frequency (MF) based on the modality which achieved the highest performance in the unimodal analyses (rsfMRI connectivity). MF was significantly correlated with symptom severity in patients with depression (eTable 4). A higher score in current depressive symptom levels (BDI, HAMD) as well as a higher number of previous hospitalizations were associated with fewer misclassifications (BDI: n=621, *r*=-0.15, *p*<0.001; HAMD: n=628, *r*=-0.20, *p*<0.001, number of hospitalizations: n=622, *r*=-.10, *p*=0.01), showing that patients with more severe current depressive symptoms and a more unfavourable previous disease course were correctly classified as patients more often. Likewise, a higher score in global assessment of functioning (GAF) in patients and a lower GAF score in healthy controls was associated with more misclassifications (HC: n=690, *r*=-.10, *p*=0.007; MDD: n=620, *r*=.17, *p*<0.001). A higher medication load in patients was also associated with fewer misclassifications (n=631, *r*=-.21, *p*<0.001). Furthermore, a higher number of misclassifications was apparent in remitted patients compared to patients with acute depressive symptoms (*F*_1,628_=7.24, *p*=0.007) and in patients without comorbidities compared to patients with comorbidities (*F*_1,628_=7.79, *p*=0.005).

## Discussion

Extending recent evidence in neuroimaging and other neurobiological research domains showing that univariate group-level differences between patients with MDD and healthy controls are small[5], we aimed to systematically evaluate ML approaches classifying patients and healthy controls on the basis of multivariate neuroimaging signatures. Importantly, we directly addressed the limitations of existing ML studies which have led to over- and underestimation of model performance, providing a much more accurate assessment of the potential of current predictive models in MDD diagnosis. In summary, training and testing a total of 2.4 million ML models on a large, harmonized sample, accuracy for predicting MDD diagnosis did not exceed 62%. Although slightly improving the 56-58% classification accuracy achieved using univariate neuroimaging and genetic markers[5], this systematic evaluation of multivariate methods revealed a disconcerting discrepancy to existing proof-of-concept studies, yielding considerably lower predictive accuracy than previously expected.[12] Our study provides four main improvements over existing ML studies: First, we reduced the common risk of producing systematically inflated predictive performance estimates due to data leakage and/or small test set size[14] by using nested cross-validation in a large sample of N=1,801 patients and controls, ensuring independent and sufficiently large test sets. Our results thus point towards small (test) sample sizes as a major driver in the current overestimation of neuroimaging-based predictability of MDD diagnosis.[12] Second, whereas previous studies mostly relied on single ML models, e.g., Support Vector Machines only, and did thus not systematically explore the space of possible ML pipelines, we employed an extensive multivariate approach providing substantially improved coverage of algorithmic search space. In addition, we also extensively sampled the hyperparameters for each algorithm. Despite these considerable efforts, classification accuracy still falls short of expectations. Note that we focused on classical ML algorithms and did not investigate more complex models e.g. based on Deep Learning.[48] Although deep learning (DL) has revolutionized ML applications, model performance will only improve if the data have nonlinear relationships exploitable at the available sample sizes, yet linear models have shown to perform on par with more complex DL approaches for structural and functional MRI up to sample sizes >10,000 subjects.[49] Future studies should, however, combine clinical samples with modality-specific, large-scale data of healthy controls (as available e.g. from the UK Biobank or ENIGMA) using for example self-supervised learning, transfer learning or semi-supervised learning approaches to increase sample size to tens of thousands to enable the exploitation of non-linear associations. Third, capitalizing on our multimodal dataset comprising structural and functional MRI as well as DTI, we tested if classification accuracy can be boosted by integrating data from neuroimaging modalities. However, even integrating all 11 modalities using different strategies did not increase performance. This suggests that modality specific models either learn so little that combining them is irrelevant or that model predictions are so highly correlated as to render their integration redundant. The latter seems plausible given that modalities with higher accuracy also show considerable correlation among each other (*r*_*max*_ = 0.47, eFigure 6). Fourth, we addressed two major shortcomings of large, multi-site ML studies, i.e. between-site variability due to data pooling and clinical heterogeneity due to unstandardized diagnostic procedures.[50] Our harmonized sample made it possible to run ML analyses on a large sample without the need of data pooling across multiple studies and acquisition processes, ef-fectively minimizing methodological heterogeneity resulting from multiple scanning sites, neuroimaging preprocessing pipelines and population differences. In addition, we were able to reduce diagnostic uncertainty by relying upon structured clinical SCID interviews for MDD diagnostics. Thus, we provide evidence that low predictive performance cannot be explained by a lack of harmonization of studies or unstandardized diagnoses as previously suggested.[50]

Aiming to explain the apparent discrepancy between the popular belief in mainstream biological psychiatry that mental disorders are in fact brain disorders[2] and a lack of neurobiological manifestations of MDD informative on the level of the individual across the most commonly investigated modalities in research today, we will discuss a number of viewpoints concerning the reliability and validity of both the neuroimaging data and the conceptualization of MDD as well as the current research design.

Addressing the debate around reliability[17, 27, 51], we show that even under conditions of perfect reliability of diagnosis or neuroimaging data, clinically useful prediction on the level of the individual patient still remains elusive. Note that this approach can, by design, only simulate perfect reliability with regard to final model predictions and thus does not speak directly to the effect different data or pre-processing pipelines might have on model training.[51] Although improved reliability of neuroimaging data could potentially lead to more stable ML models, this seems unlikely given the complete lack of correlation between known reliability estimates of MRI data and our classification results.

Apart from concerns about reliability, we may also question the validity of neuroimaging data in terms of its ability to capture the neurobiological information necessary for explaining the MDD phenotype. If we assume current methods fall short in this regard, there are several research directions that could enhance our understanding of the disorder. These include higher spatial or temporal resolution, more advanced experimental paradigms or data preprocessing techniques, as well as longitudinal research designs that can model changes in an individual’s neurobiology associated with current symptoms and episodes.[52, 53] Additionally, the complexity of the MDD phenotype might require a more comprehensive approach that incorporates interactions between neurobiology, the entire body, as well as the environment.[54] However, since there is no established formal theory of the neurobiology of depression, it is uncertain which neuroimaging methods will be best suited to capture clinically relevant information.[55]

On the other hand, if we assume that the information relevant for explaining behaviour and mental processes is present in current neuroimaging modalities, issues of biological validity of the MDD construct appear plausible. Since clinical heterogeneity in MDD has been extensively described[28], focusing on clinically relevant outcomes and longitudinal data, even across diagnoses, rather than MDD diagnosis itself might be better suited to yield high-accuracy predictions, e.g. associating neuroimaging markers with long-term disease trajectories.[8, 56–58] Likewise, investigating symptoms rather than syndromes has been promoted lately, with network theory of psychopathology providing one conceptual framework possibly able to model symptom dynamics independent of psychiatric category.[59] Indeed, our results regarding correlations of misclassification frequency provide support for associations between symptom severity and neurobiological markers, suggesting that patients with higher levels of current symptoms, lower global functioning and more unfavourable disease courses in the past are easier to detect and correctly classify. Although providing a potential target for MDD subgroup identification beyond a more general MDD category, our complementary subgroup analyses focusing on acutely and recurrently depressed patients, respectively, did not increase predictive performance. This, however, might be due to the reduced sample sizes available during model training of depressive subgroups. Other research directions such as the Research Domain Criteria aim at identifying biologically motivated descriptions of mental disorders for which a direct link between neurobiology and cognitive processes is a necessary requirement.[60] However, the current results indicate that it might be difficult to find biological predictors for all patients currently covered under the umbrella of the MDD diagnosis.

In the same vein, a strictly reductionist case-control design in neuroimaging might be too simplistic to adequately model the complex relationship between brain and behaviour.[1, 61] Modelling complexity could be increased using e.g. normative modelling approaches that capture deviations of the individual patient, overcoming the necessity for a common biological cause across all MDD patients.[62] Similarly, identifying biotypes of mental disorders through clustering across DSM diagnoses might constitute a promising way forward.[40, 56, 63] Furthermore, given the complex, nonlinear dynamics of brain processes and symptom interactions, dynamical systems theory within computational psychiatry provides another conceptual framework that could be able to overcome simplistic reductionism and model the neurobiological complexity of MDD.[64] It also provides one way of moving towards quantitative theories of depression, e.g. network theory of psychopathology.[59] However, more research is needed to investigate whether these approaches are actually able to increase clinically relevant predictions on the level of the individual patient.

## Conclusions

In summary, we show that although multivariate neuroimaging markers increase predictive performance compared to univariate analyses, classification on the level of the individual patient – even under optimal conditions – does not reach clinically relevant levels. How biological Precision Psychiatry can deliver more accurate individualized prediction to improve treatment and patient care remains a central open question at this point.

## Data Availability

All data produced in the present study are available upon reasonable request to the authors

## ACKNOWLEDGEMENTS

This work was funded by the German Research Foundation (DFG grants FOR2107 KI588/14-1, and KI588/14-2, and KI588/20-1, KI588/22-1 to Tilo Kircher, Marburg,

Germany; STR1146/18-1 to Benjamin Straube, Marburg, Germany; HA7070/2-2, HA7070/3, and HA7070/4 to Tim Hahn, Münster, Germany; Dan3/012/17 to Udo Dannlowski) and MzH 3/020/20 from the Interdisciplinary Center for Clinical Research of the medical faculty of Münster to Tim Hahn. The project was further supported by the cluster project “The Adaptive Mind”, funded by the Excellence Program of the Hessian Ministry of Higher Education, Science, Research and Art to Tilo Kircher and Benjamin Straube.

This work is part of the German multicenter consortium “Neurobiology of Affective Disorders. A translational perspective on brain structure and function”, funded by the German Research Foundation (Deutsche Forschungsgemeinschaft DFG; Forschungsgruppe/Research Unit FOR2107).

Principal investigators (PIs) with respective areas of responsibility in the FOR2107 consortium are: Work Package WP1, FOR2107/MACS cohort and brainimaging: Tilo Kircher (speaker FOR2107; DFG grant numbers KI 588/14-1, KI 588/14-2), Udo Dannlowski (co-speaker FOR2107; DA 1151/5-1, DA 1151/5-2), Axel Krug (KR 3822/5-1, KR 3822/7-2), Igor Nenadic (NE 2254/1-2), Carsten Konrad (KO 4291/3-1). WP2, animal phenotyping: Markus Wöhr (WO 1732/4-1, WO 1732/4-2), Rainer Schwarting (SCHW 559/14-1, SCHW 559/14-2). WP3, miRNA: Gerhard Schratt (SCHR 1136/3-1, 1136/3-2). WP4, immunology, mitochondriae: Judith Alferink (AL 1145/5-2), Carsten Culmsee (CU 43/9-1, CU 43/9-2), Holger Garn (GA 545/5-1, GA 545/7-2). WP5, genetics: Marcella Rietschel (RI 908/11-1, RI 908/11-2), Markus Nöthen (NO 246/10-1, NO 246/10-2), Stephanie Witt (WI 3439/3-1, WI 3439/3-2). WP6, multi-method data analytics: Andreas Jansen (JA 1890/7-1, JA 1890/7-2), Tim Hahn (HA 7070/2-2), Bertram Müller-Myhsok (MU1315/8-2), Astrid Dempfle (DE 1614/3-1, DE 1614/3-2). CP1, biobank: Petra Pfefferle (PF 784/1-1, PF 784/1-2), Harald Renz (RE 737/20-1, 737/20-2). CP2, administration. Tilo Kircher (KI 588/15-1, KI 588/17-1), Udo Dannlowski (DA 1151/6-1), Carsten Konrad (KO 4291/4-1).

Data access and responsibility: All PIs take responsibility for the integrity of the respective study data and their components. All authors and coauthors had full access to all study data.

Acknowledgements and members by Work Package (WP): WP1: Henrike Bröhl, Katharina Brosch, Bruno Dietsche, Rozbeh Elahi, Jennifer Engelen, Sabine Fischer, Jessica Heinen, Svenja Klingel, Felicitas Meier, Tina Meller, Julia-Katharina Pfarr, Kai Ringwald, Torsten Sauder, Simon Schmitt, Frederike Stein, Annette Tittmar, Dilara Yüksel (Dept. of Psychiatry, Marburg University). Mechthild Wallnig, Rita Werner (Core-Facility Brainimaging, Marburg University). Carmen Schade-Brittinger, Maik Hahmann (Coordinating Centre for Clinical Trials, Marburg). Michael Putzke (Psychiatric Hospital, Friedberg). Rolf Speier, Lutz Lenhard (Psychiatric Hospital, Haina). Birgit Köhnlein (Psychiatric Practice, Marburg). Peter Wulf, Jürgen Kleebach, Achim Becker (Psychiatric Hospital Hephata, Schwalmstadt-Treysa). Ruth Bär (Care facility Bischoff, Neukirchen). Matthias Müller, Michael Franz, Siegfried Scharmann, Anja Haag, Kristina Spenner, Ulrich Ohlenschläger (Psychiatric Hospital Vitos, Marburg). Matthias Müller, Michael Franz, Bernd Kundermann (Psychiatric Hospital Vitos, Gießen). Christian Bürger, Katharina Dohm, Fanni Dzvonyar, Verena Enneking, Stella Fingas, Katharina Förster, Janik Goltermann, Dominik Grotegerd, Hannah Lemke, Susanne Meinert, Nils Opel, Ronny Redlich, Jonathan Repple, Katharina Thiel, Kordula Vorspohl, Bettina Walden, Lena Waltemate, Alexandra Winter, Dario Zaremba (Dept. of Psychiatry, University of Münster). Harald Kugel, Jochen Bauer, Walter Heindel, Birgit Vahrenkamp (Dept. of Clinical Radiology, University of Münster). Gereon Heuft, Gudrun Schneider (Dept. of Psychosomatics and Psychotherapy, University of Münster). Thomas Reker (LWL-Hospital Münster). Gisela Bartling (IPP Münster). Ulrike Buhlmann (Dept. of Clinical Psychology, University of Münster).

WP2: Marco Bartz, Miriam Becker, Christine Blöcher, Annuska Berz, Moria Braun, Ingmar Conell, Debora dalla Vecchia, Darius Dietrich, Ezgi Esen, Sophia Estel, Jens Hensen, Ruhkshona Kayumova, Theresa Kisko, Rebekka Obermeier, Anika Pützer, Nivethini Sangarapillai, Özge Sungur, Clara Raithel, Tobias Redecker, Vanessa Sandermann, Finnja Schramm, Linda Tempel, Natalie Vermehren, Jakob Vörckel, Stephan Weingarten, Maria Willadsen, Cüneyt Yildiz (Faculty of Psychology, Marburg University).

WP4: Jana Freff (Dept. of Psychiatry, University of Münster). Susanne Michels, Goutham Ganjam, Katharina Elsässer (Faculty of Pharmacy, Marburg University). Felix Ruben Picard, Nicole Löwer, Thomas Ruppersberg (Institute of Laboratory Medicine and Pathobiochemistry, Marburg University).

WP5: Helene Dukal, Christine Hohmeyer, Lennard Stütz, Viola Lahr, Fabian Streit, Josef Frank, Lea Sirignano (Dept. of Genetic Epidemiology, Central Institute of Mental Health, Medical Faculty Mannheim, Heidelberg University). Stefanie Heilmann-Heimbach, Stefan Herms, Per Hoffmann (Institute of Human Genetics, University of Bonn, School of Medicine & University Hospital Bonn). Andreas J. Forstner (Institute of Human Genetics, University of Bonn, School of Medicine & University Hospital Bonn).

WP6: Anastasia Benedyk, Miriam Bopp, Roman Keßler, Maximilian Lückel, Verena Schuster, Christoph Vogelbacher (Dept. of Psychiatry, Marburg University). Jens Sommer, Olaf Steinsträter (Core-Facility Brainimaging, Marburg University). Thomas W.D. Möbius (Institute of Medical Informatics and Statistics, Kiel University).

CP1: Julian Glandorf, Fabian Kormann, Arif Alkan, Fatana Wedi, Lea Henning, Alena Renker, Karina Schneider, Elisabeth Folwarczny, Dana Stenzel, Kai Wenk, Felix Picard, Alexandra Fischer, Sandra Blumenau, Beate Kleb, Doris Finholdt, Elisabeth Kinder, Tamara Wüst, Elvira Przypadlo, Corinna Brehm (Comprehensive Biomaterial Bank Marburg, Marburg University).

The FOR2107 cohort project (WP1) was approved by the Ethics Committees of the Medical Faculties, University of Marburg (AZ: 07/14) and University of Münster (AZ: 2014-422-b-S).

Biosamples and corresponding data were sampled, processed and stored in the Marburg Biobank CBBMR.

Biomedical financial interests or potential conflicts of interest: Tilo Kircher received unrestricted educational grants from Servier, Janssen, Recordati, Aristo, Otsuka, neuraxpharm. Markus Wöhr is scientific advisor of Avisoft Bioacoustics.

