## Supplementary Materials for "A Systematic Evaluation of Machine Learning-based Biomarkers for Major Depressive Disorder across Modalities"


eMethods 1. Sample exclusion criteria **3**

eMethods 2. Diagnosis, remission status, and medication index **3**

eMethods 3. Definition of subgroups **3**

eMethods 4. Assessment of childhood maltreatment and social support **3**

eMethods 5. Polygenic risk score for depression **4**

eMethods 6. Magnetic Resonance Imaging **4**

eMethods 7. T1-weighted MRI **4**

eMethods 8. Functional MRI image acquisition **5**

eMethods 9. Resting-State fMRI **5**

eMethods 10. Task-based fMRI **5**

eMethods 11. DTI image acquisition and preprocessing **6**

eMethods 12. Graph network parameters **6**

eMethods 13. Reliability Analysis **7**

eMethods 14. Machine Learning Analysis **7**

eFigure 1. Classification results for healthy participants versus patients with acute MDD **9**

eFigure 2. Classification results for healthy participants versus patients with recurrent MDD **10**

eFigure 3. Classification results for male participants only (HC versus MDD) **11**

eFigure 4. Classification results for female participants only (HC versus MDD) **12**

eFigure 5. Classification results for participants within an age range of 24 to 28 years (HC versus MDD) **13**

eFigure 6. Correlations of model predictions across modalities for HC versus MDD. **14**

eTable 1. Classification accuracy for multivariate biomarkers based on structural MRI. **16**

eTable 2. Classification accuracy for multivariate biomarkers based on functional MRI. **18**

eTable 3. Classification accuracy for multivariate biomarkers using neuroimaging modality integration. **20**

eTable 4. Associations of symptom severity and misclassification frequency. **22**

eTable 5. Classification accuracy based on structural MRI for HC vs acute MDD. **23**

eTable 6. Classification accuracy based on functional MRI for HC vs acute MDD. **25**

eTable 7. Classification accuracy using neuroimaging modality integration for HC vs acute MDD. **27**

eTable 8. Classification accuracy based on structural MRI for HC vs recurrent MDD. **29**

eTable 9. Classification accuracy based on functional MRI for HC vs recurrent MDD. **31** eTable 10. Classification accuracy using neuroimaging modality integration for HC vs recurrent MDD. **33**

eTable 11. Classification accuracy based on structural MRI for HC vs MDD (male). **35**

eTable 12. Classification accuracy based on functional MRI for HC vs MDD (male). **37**

eTable 13. Classification accuracy using neuroimaging modality integration for HC vs MDD (male). **39**

eTable 14. Classification accuracy based on structural MRI for HC vs MDD (female). **41**

eTable 15. Classification accuracy based on functional MRI for HC vs MDD (female). **43**

eTable 16. Classification accuracy using neuroimaging modality integration for HC vs MDD (female). **45**

eTable 17. Classification accuracy based on structural MRI for HC vs MDD (age 24 - 28). **47**

eTable 18. Classification accuracy based on functional MRI for HC vs MDD (age 24 - 28). **49**

eTable 19. Classification accuracy using neuroimaging modality integration for HC vs MDD (age 24 - 28). **51**

eReferences **54**

**eMethods 1. Sample exclusion criteria**

Participants with a history of neurological (e.g., concussion, stroke, tumor, neuro-inflammatory diseases) or medical conditions (e.g., cancer, chronic inflammatory or autoimmune diseases, heart diseases, diabetes mellitus, infections), as well as those who self-identified as non-Caucasian, were excluded from the analysis. Non-Caucasian participants were excluded since the FOR2107 MACS cohort was originally focused on genetic and neuroimaging analyses, thus providing greater genetic homogeneity. The exclusion criteria were the same for both the healthy and depressive participants. Additionally, healthy participants were further excluded if they had a current or past history of psychiatric illness. A sample of 1,801 participants remained for the analyses. Please note that the number of participants finally used in the machine learning models is reduced due to data availability and quality checks within the specific neuroimaging modality. Please see below for a detailed description of the number of participants used in each analysis.

**eMethods 2. Diagnosis, remission status, and medication index**

Each participant underwent a structured clinical interview for DSM-IV (SCID-I) to evaluate their current and lifetime psychopathological diagnoses.[1] For a Major Depressive Disorder (MDD) diagnosis, the participant had to meet the DSM-IV criteria, which includes experiencing at least five of nine core symptoms most of the day nearly every day for two weeks or more within the last four weeks, and must cause significant distress or impairment. At least one of the symptoms must either be a depressed mood or a markedly diminished interest or pleasure. Other symptoms include significant weight loss or gain, insomnia or hypersomnia, psychomotor agitation or retardation, fatigue or loss of energy, feelings of worthlessness or excessive guilt, diminished ability to concentrate or think clearly, and recurrent thoughts of death or suicide. Only participants with a primary MDD diagnosis were included in the MDD sample. MDD patients were not excluded if they fulfilled the criteria of an additional comorbid psychopathological diagnosis. Partial remission was defined either by (1) a presence of some major depressive symptoms, but full criteria are no longer met or (2) no major depressive symptoms, but the period of remission has been less than two months. Complete remission was defined as the absence of diagnostic criteria for depression for at least two consecutive months.

A medication load index was calculated expressing the combined dosage level of all current psychiatric medication. Trained personnel conducted interviews to assess current psychiatric medication, which were then classified categorized based on active ingredients (e.g. SSRIs, benzodiazepines). Psychiatric medication was coded based on established dosage-dependent cut-offs, with each active ingredient given a score of 0, 1, or 2.[2] The scores for all psychiatric medication were added together to create the final medication index. A similar procedure has been used in previous publications.[3,4]

**eMethods 3. Definition of subgroups**

In the secondary analyses that focused on acutely depressed patients, we excluded individuals with fully remitted MDD from the sample. In the secondary analyses that focused on recurrently depressed patients, we only included MDD subjects who had a history of at least two inpatient stays related to their depressive disorder. In addition, we ran analyses using only female or only male participants to reduce heterogeneity of the sample. We ran a final analysis using participants within a 5-year age range to reduce potential age effects confounding the classification analyses. To maximize the number of participants within a small age bin, we used an age range of 24 to 28, which showed the highest density of patients and controls.

**eMethods 4. Assessment of childhood maltreatment and social support**

The childhood trauma questionnaire (CTQ), a well-established tool, was used to evaluate childhood maltreatment in both patients and controls.[5] A sum score based on the five maltreatment subscales was computed and used in all analyses that included childhood maltreatment. No CTQ sum score was available for 13 individuals, resulting in a sample of N=1788 that could be used for the CTQ analysis.
The Social Support Questionnaire (F-SozU), an established German self-report measure, was used to evaluate perceived social support across three subscales (perceived emotional support, instrumental support, and social integration), with a sum score used in all analyses.[6] No social support sum score was available for 10 individuals, resulting in a sample of N=1791 that could be used for the social support analysis.

**eMethods 5. Polygenic risk score for depression**

Genetic data and preprocessing procedures have been described in previous publications.[7,8] Genome-wide genotyping was performed using the PsychArray BeadChip, followed by quality control (QC) and imputation using PLINK v1.9.[7,9–11] The genotype data were imputed to the 1000 Genomes phase 3 reference panel using SHAPEIT and IMPUTE2.[12,13] Genetic data were available for a total of n=1683 individuals. Related participants were identified using the PLINK command –genome and one individual of each related pair (PI-HAT ≥12.5) was excluded for a specific analysis. A total of 67 related participants were excluded for the main HC versus MDD analysis, resulting in a final sample of n=1616. Finally, a polygenic risk score (PRS) was calculated from the genetic data using the PRS-CS method and summary statistics from a recent depression GWAS, with the global shrinkage parameter phi estimated automatically using the PRS-CS-auto method ($\varphi=1 \cdot 30\times10^{-4}$).[14,15] This resulted in only a single PRS that was used in the present analysis. Note that the GWAS used for our PRS calculation investigated a wider depression phenotype which was not restricted to participants with a DSM MDD diagnosis. Therefore, we use the term 'depression PRS' throughout the manuscript. The FOR2107 MACS data used in this study were independent of the depression GWAS.

**eMethods 6. Magnetic Resonance Imaging**

Magnetic resonance imaging (MRI) data, protocol and preprocessing procedures (eMethods 6-12) have been described in a previous publication but are described here again to provide direct access to the relevant information.[8]

Data for all brain-based modalities using MRI were obtained through the use of two 3T whole body MRI scanners. The first scanner, located in Marburg, Germany, is the MAGNETOM Trio Tim with software version Syngo MR B17 (Siemens, Erlangen, Germany), equipped with a 12-channel head matrix Rx-coil. The second scanner, located in Münster, Germany, is the MAGNETOM Prisma with software version Syngo MR D13D (Siemens, Erlangen, Germany), equipped with a 20-channel head matrix Rx-coil. A GRAPPA acceleration factor of 2 was used during the acquisition process. Pulse sequence parameters were standardized across both sites to the extent possible (see below for differences between sites). More information on pulse sequence parameters, as well as quality assurance protocols, can be found in reference [16].

**eMethods 7. T1-weighted MRI**

The following parameters were used to acquire the structural MRI data: TE = 2.26ms (Marburg), TE = 2.28ms (Münster), TR = 1,900ms (Marburg), TR = 2,130ms (Münster), FoV = 256 mm, matrix = 256 × 256, slice thickness = 1 mm, distance factor = 50%, phase encoding direction anterior >> posterior, flip angle = 8°, bandwidth 200 Hz/Px, ascending acquisition, axial acquisition, 192 slices. Of the original sample of 1,801 subjects, 28 did not have a T1 scan available and 31 were excluded due to low image quality or artifacts, leaving a remaining sample of 1,742 subjects. Additionally, nine subjects were excluded from the Freesurfer analysis due to poor segmentation quality, leaving a final sample of 1,733 subjects. The quality of the images was assessed through visual inspection by a trained expert and by checking image homogeneity using the CAT12 toolbox. The voxel-based morphometry (VBM) was preprocessed using the default parameters in the CAT12 MATLAB toolbox (version r1450), which included bias correction, tissue segmentation, and normalization to MNI-space using linear and non-linear transformations.[17] Normalization was performed using a pre-computed high-dimensional DARTEL template, and the modulated gray matter images were smoothed with a Gaussian kernel of 8 mm FWHM. An absolute threshold masking value of 0.1 was used for all analyses, as recommended in the CAT12 manual (http://www.neuro.uni-jena.de/cat/).

Automated segmentation was performed using Freesurfer's (Version 5.3) cortical and subcortical parcellation stream based on the Desikan-Killiany atlas.[18] This allowed for the extraction of measures for 68 cortical regions (34 on each hemisphere), 14 subcortical regions (7 on each hemisphere), 4 ventricles (2 on each hemisphere), and total intracranial volume (ICV) for each participant. Global measures of cortical and subcortical surface, thickness, and volume were also calculated both per and across hemispheres, resulting in a total of 166 parameters that were used in the statistical analyses. Default parameters were used for the segmentation (https://surfer.nmr.mgh.harvard.edu/), and segmentation quality was reviewed visually and based on statistical outlier analysis following standardized protocols by the ENIGMA consortium.[19]

**eMethods 8. Functional MRI image acquisition**

The two functional MRI paradigms (face matching and resting-state) were based on a $T_2$*-weighted echo-planar imaging (EPI) sequence that was sensitive to blood oxygen level-dependent (BOLD) contrast. The following parameters were used: TE = 30 ms (Marburg), TE = 29ms (Münster), TR = 2,000 ms, FoV = 210 mm, matrix = 64 × 64, slice thickness = 3.8 mm, distance factor = 10%, phase encoding direction anterior >> posterior, flip angle = 90°, no parallel imaging, bandwidth 2,232 Hz/Px, ascending acquisition, axial acquisition, 33 slices, slice alignment parallel to AC-PC line tilted 20° in the dorsal direction. For resting-state fMRI, 237 interleaved and ascending measurements (8 minutes) were acquired, tilted with -20° against the anterior and posterior commission alignment (AC-PC alignment) and the following parameters: Base resolution=64, Bandwith=2,232 Hz/Px, Echo spacing=0.51ms, EPI factor=64, TR=2s). Participants were asked to keep their eyes closed until the end of the resting state session.

**eMethods 9. Resting-State fMRI**

Resting-state data was preprocessed using the CONN (v18b) MATLAB toolbox and its default volume-based MNI preprocessing pipeline.[20] The functional and structural images underwent several preprocessing steps, including functional realignment and unwarp, slice-timing correction, outlier identification based on the ART method, direct segmentation, and normalization of functional and structural images, as well as functional smoothing using an 8mm FWHM kernel. To regress out potential noise artefacts in the functional data, CONN's denoising step was used with default parameters. This process involved an anatomical component-based noise correction procedure (aCompCor), where cerebral white matter and cerebrospinal areas' noise components (5 PCA components), estimated subject-motion parameters (12 parameters including 6 motion parameters and their associated first-order derivatives), and identified outlier scans were regressed out.[21] Additionally, temporal band-pass filtering was applied to remove low frequencies under 0.008 Hz and high frequencies above 0.09 Hz.

Resting-state and $T_1$ data from 1372 subjects were available for the analyses. 14 subjects were excluded due to recent use of tranquilizers such as benzodiazepine or Z-drugs, which are known to affect functional MRI data. 16 subjects were excluded after visual quality checks revealed poor quality of the functional or structural segmentation and normalization or distribution of correlation values after denoising did not follow a normal distribution, as determined by quality assurance plots in the CONN toolbox. 11 subjects were excluded due to significant motion artifacts that resulted in fewer than 5 minutes of usable resting-state image time points after scrubbing. As a result, data from a total of 1331 subjects was available for the resting-state analyses.

The connectivity matrices for each participant were generated by computing the bivariate Pearson's correlation coefficient between the time-series of each region of the 17 networks Schaefer atlas (100 parcels). The Amplitude of Low-Frequency Fluctuations (ALFF) maps were computed from the resting-state time-series. The ALFF represents a measure of the BOLD signal power within the frequency band of interest, defined as the root mean square of the BOLD signal at each voxel after filtering.[22] To obtain a relative measure of BOLD signal power, the fractional ALFF (fALFF) was computed as the ratio of the root mean square of the BOLD signal at each voxel after versus before low- or band-pass filtering. Regional homogeneity was measured using the Local Correlation (LCOR) method, which is a more robust version of the original ReHo definition by Zang et al. (2004) across different data resolutions and neighboring sizes. LCOR is defined as the average of correlation coefficients between each individual voxel and a region of neighboring voxels.[20] For a more detailed description of the Integrated Local Correlation and a comparison to ReHo, see Deshpande et al. (2009).[23]

**eMethods 10. Task-based fMRI**

Functional MRI data from a well-established emotional face matching paradigm was used.[24] In summary, the experimental design involved presenting images of faces depicting fear or anger during the task, while geometric shapes served as the control condition. Each trial consisted of a target image presented at the top and two additional images displayed at the bottom left and right, one of which matched the target image. Participants were instructed to indicate which of the bottom images was identical to the target image by pressing the corresponding button. The preprocessing steps for this dataset have also been described previously.[25]

The same image acquisition parameters used for the resting-state scans were used for the emotional face matching paradigm. Subjects whose overall movement exceeded 2mm were excluded from the final sample to avoid motion artifacts, and a visual quality check was performed to exclude subjects with visible artifacts. Subjects under acute medication with tranquilizers were also excluded from the analyses. The fMRI responses for both the faces and shapes conditions were modeled using a block design and the canonical hemodynamic response function in SPM8, with high-pass filtering (cut-off frequency of 1/128 Hz) applied to attenuate low-frequency components. Contrast images were created by contrasting beta images of the faces and shapes conditions. Of the 1364 participants with available face matching fMRI data, 7 subjects under acute medication with tranquilizers were excluded, leaving a sample of 1357. An additional 109 subjects with more than 2mm of movement were excluded, leaving a sample of 1248. 14 subjects with low image quality or artifacts, as determined by visual inspection by a trained expert, were also excluded. Thus, a final sample of 1234 subjects was available for the face matching task-based fMRI analyses.

**eMethods 11. DTI image acquisition and preprocessing**

DTI data were acquired using a GRAPPA acceleration factor of two. 56 axial slices with no gap were measured with an isotropic voxel size of 2.5×2.5×2.5 mm3 (TE = 90 ms, TR = 7300 ms). Five non-DW images (*b* = 0 s/mm2) and 2 × 30 DW images with a *b*-value of 1000 s/mm2 were acquired.

The preprocessing procedure of the DTI images is described in more detail in a previous publication.[26] In summary, realignment and eddy correction of the Diffusion-weighted images (DWI) was done using FSL eddy.[27–30] Next, the reconstruction of the anatomical connectome was achieved using the CATO toolbox, which models the measured signal of a single voxel by a tensor describing the preferred diffusion direction per voxel.[31] CATO uses the RESTORE algorithm, which estimates the diffusion tensor while simultaneously identifying and removing outliers, thereby reducing the impact of physiological noise artifacts.[29,30] Deterministic tractography was used to reconstruct white matter paths. To this end, eight seeds were started per voxel, and for each seed, a tractography streamline was constructed by following the main diffusion direction from voxel to voxel. Stop criteria included reaching a voxel with a fractional anisotropy <0.1, making a sharp turn of >45°, reaching a gray matter voxel, or exiting the brain mask. Given the poorer DWI signal-to-noise ratio in subcortical regions and the dominant effect of subcortical regions on network properties, we decided to use the Lausanne parcellation including 114 cortical brain regions, a subdivision of FreeSurfer's Desikan Killiany Atlas, as we have done in previous work.[26,32–34] Matrix entries represent the weights of the graph edges. Network edges were weighted according to fractional anisotropy (FA), mean diffusivity (MD), and number of streamlines (NOS). In total, DTI data was available for 1558 participants. After quality control (see below), 55 subjects were excluded, resulting in a final sample of 1503 participants for all DTI-based analyses.

Outlier detection during quality checks were based on the following four metrics: the average number of streamlines, the average fractional anisotropy, the average prevalence of each subject's connections (low value if the subject has "odd" connections), and the average prevalence of each subjects connected brain regions (high value, if the participant misses commonly found connections). Then, quartiles (Q1, Q2, Q3) and the interquartile range (IQR=Q3-Q1) was computed for every metric across the group. A datapoint was declared an outlier if its value was below Q1-1.5*IQR or above Q3+1.5*IQR on any of the four metrics.

**eMethods 12. Graph network parameters**

To create the DTI and resting-state fMRI connectivity matrices, a binary adjacency matrix was produced using different methods. For DTI, a binary adjacency matrix was computed based on the number of streamlines, where all edges with less than three number of streamlines were set to 0 and all other edges were set to 1. For resting-state fMRI, a binary adjacency matrix was created by setting the top 15 percent of connections (highest correlation coefficient) to 1, and all other edges were set to 0. These adjacency matrices were then used to derive a variety of graph parameters using PHOTONAI Graph (https://github.com/wwu-mmll/photonai_graph). The selected graph metrics were global efficiency, local efficiency, clustering coefficient, degree centrality, betweenness centrality, and degree assortativity (for an introduction of graph metrics for brain connectivity, see [35]). Global efficiency was computed as the average inverse shortest path length between all node pairs, while local efficiency was computed on node neighborhoods. Clustering coefficient was defined as the average probability that the neighbors of a node are also mutually connected. Degree centrality was defined as the number of nodes connected to the node of interest, while betweenness centrality was defined as the proportion of shortest paths in the network that pass through the node of interest. Clustering coefficient, degree centrality, and betweenness centrality were calculated per node and additionally averaged across nodes, resulting in a total of 348 network parameters. For further information on graph metrics for brain connectivity, see (36).

**eMethods 13. Reliability Analysis**

The calculation of BACC from MCC requires information on prevalence $\phi$ and bias $\beta$ as is described in [36]. In this context, prevalence measures the frequency of MDD cases in the test set while bias refers to how likely the classifier predicts an MDD label in the test set. We corrected MCC independently for all test sets of the 10 folds and converted it back to BACC using the frequency of MDD cases and frequency of MDD predictions in the corresponding test sets.

**eMethods 14. Machine Learning Analysis**

*General*
All ML analyses were done using Python 3.8, scikit-learn version 0.24.2 and photonai version 2.2.0. For all analyses, a strict nested cross-validation scheme with 10 inner and 10 outer folds was used to train, optimize, and evaluate the ML pipeline. Imputation, scaling, feature selection, and dimensionality reduction was always done on the training set and then applied to the validation or test set to ensure the independence assumption of the cross-validation. The inner cross-validation was used to optimize all hyperparameters. Once an optimal configuration based on mean balanced accuracy across the 10 validation sets was found, an ML pipeline with this configuration was trained on the complete training set and evaluated on the 10% test set. This was done for all 10 outer folds.

*Imputation*
The first step of the ML pipeline was an imputation of missing values. We used sklearn's SimpleImputer with default parameters. Missing values for a single feature of the training and test data were imputed with the mean of the training set. No hyperparameter tuning was performed for this pipeline element.

*Scaling*
The second step of the ML pipeline was an individual scaling of every feature using sklearn's RobustScaler method with default settings. This scaling method removes the median and scales the data according to the interquartile range. The interquartile is the range between the 1st quartile (25th quantile) and the 3rd quartile (75th quantile). Scaling of the features in the test set is done using the median and interquartile range of the training set to avoid data leakage.

*Feature Selection and Dimensionality Reduction*
The third step of the ML pipeline was a photonai Switch, an element that evaluates two or more other elements at the same position in the complete pipeline. In this case, the photonai Switch chose between a feature selection or a dimensionality reduction element. The selection of an ML element within a photonai Switch itself can be seen as a hyperparameter that is optimized by photonai. The feature selection was done using sklearn's f_classif() method that computes a statistical F-value from an ANOVA model differentiating between healthy participants and patients with MDD. This was combined with sklearn's selection method SelectPercentile, selecting a predefined percentage of features with the F-values. For our analyses, we used 5, 10, and 50 percent of the features as selection criterion. The dimensionality reduction as an alternative pipeline element was implemented using a Principal Component Analysis (PCA) from sklearn. A PCA is a linear dimensionality reduction method using Singular Value Decomposition to project the data into a lower dimensional space of orthogonal components. We used sklearn's default parameters to perform a full variance decomposition using all possible principal components. This will reduce the dimensionality of the data to either the number of features or the number of samples, depending on what is lower. The only hyperparameter of the PCA step was therefore whether to do the dimensionality reduction or not. In combination, this resulted in 6 possible hyperparameter configurations (4 for feature selection, 2 for dimensionality reduction).

*Classification Algorithms*
The last step of the ML pipeline consisted of one of six classification algorithms from different categories. First, a boosting ensemble method was evaluated using sklearn's GradientBoostingClassifier. It uses a decision tree classifier as base estimator and builds an additive model in a forward stage-wise fashion. As a hyperparameter, the number of boosting stages (n_estimators) was set to 10, 25, or 50. Second, a bagging ensemble implemented as a Random Forest was evaluated using sklearn's RandomForestClassifier. Two hyperparameters were optimized to control the amount of overfitting. The maximum number of features used for each split (max_features) was set to either the square root or log2 of the total number of features. The minimum samples required in a leaf of the individual decision trees was set to either 1%, 10%, or 20% of the total number of samples in the training set. Third, to evaluate the potential of linear models, we used a logistic regression as

implemented in sklearn's LogisticRegression class. To control the amount of overfitting, we optimized the penalty added to the cost function using either L1 (lasso), L2 (ridge), or elastic net. In addition, we set the regularization parameter C to 0.0001, 0.01, 1, 100, or 10,000. Fourth, representing kernel methods which are very popular in multivariate neuroimaging data analysis, a support vector machine was evaluated using either the LinearSVM or SVM classes from sklearn. We used either a linear, polynomial, or radial basis function kernel with a C value of $10^{-8}$, $10^{-6}$, $10^{-4}$, $10^{-2}$, 1, $10^{2}$, $10^{4}$, $10^{6}$, or $10^{8}$. Fifth, a Gaussian naive Bayes classifier was evaluated as implemented in sklearn's GaussianNB class with default hyperparameters. Sixth, a k-nearest neighbour classification algorithm was evaluated as implemented in sklearn's KNeighborsClassifier class, optimizing the number of possible neighbours (5, 10, 15).

*Number of Evaluated ML Pipelines*

As reported in the main article, a total of about 2.4 million ML pipelines were trained in this project. The total number of pipelines are calculated below. Number of possible hyperparameter configurations are listed in parentheses next to the corresponding pipeline step.

1. [Imputation (1)] x [Scaling (1)] x [Feature Selection (4) + Dimensionality Reduction (2)] x [BC (3) + RF (6) + LR (15) + SVM (27) + NB (1) + kNN (3)] = 330 ML pipelines
2. 330 pipelines x 10 inner folds x 10 outer folds + 6 optimal configurations (one per classifier) x 10 outer folds = 33,060 pipelines
3. 33,060 pipelines x 12 modalities (11 unimodal and 1 PCA-based modality integration) = 396,720 pipelines
4. 396,720 pipelines x 6 subgroup analyses (all, acute, recurrent, male, female, age bin) = 2,380,320 pipelines

*Modality Integration and Voting Ensemble*

Modality integration was achieved using two different methods. First, the neuroimaging data was compressed and concatenated into one data matrix. Dimensionality reduction was implemented using a principal component analysis on every neuroimaging modality separately. All possible principal components were used to create the resulting data matrix, i.e. n_components of sklearn's PCA() method was set to "None" to allow for a full variance decomposition. The number of principal components was thus limited by either the training set sample size or the number of initial features, depending on the specific modality. The resulting data matrix was used as input to the ML pipelines described above. Importantly, this PCA-based modality integration was also done within the nested-cross validation, i.e. the PCA's were trained on the training set and applied to the test set. This was implemented using photonai's Stack and Branch pipeline elements. Second, an ensemble voting algorithm was used to combine predictions of all unimodal models. In order to build the voting ensemble, the optimized ML pipelines in every outer fold across neuroimaging modalities were used to classify between healthy participants and patients with MDD in the corresponding test set. For one participant, this resulted in 11 (modalities) x 6 (classification algorithm pipelines described above) predicted labels (HC or MDD). If data for a specific neuroimaging modality was not available for a participant, the number of usable predictions within the ensemble was thus reduced. A final prediction for every participant was calculated using a majority vote method. Simply put, the final prediction was determined by the most frequent prediction (HC or MDD). Finally, the classification metrics were calculated exactly as in the unimodal models and mean and standard deviation across the 10 outer folds are reported. Depending on which model predictions were used in the ensemble, different voting strategies could be evaluated. One ensemble model contained predictions from all algorithms and all modalities. Another aggregated across modalities only, keeping the classification algorithms separately which resulted in a final classification for every algorithm. A final ensemble model aggregated across classification algorithms but keeping the neuroimaging modalities separately. Note that this final method actually did not combine predictions across neuroimaging modalities and thus cannot be considered a modality integration analysis. Still, it evaluates the use of an algorithm ensemble for every modality.

**eFigure 1. Classification results for healthy participants versus patients with acute MDD**

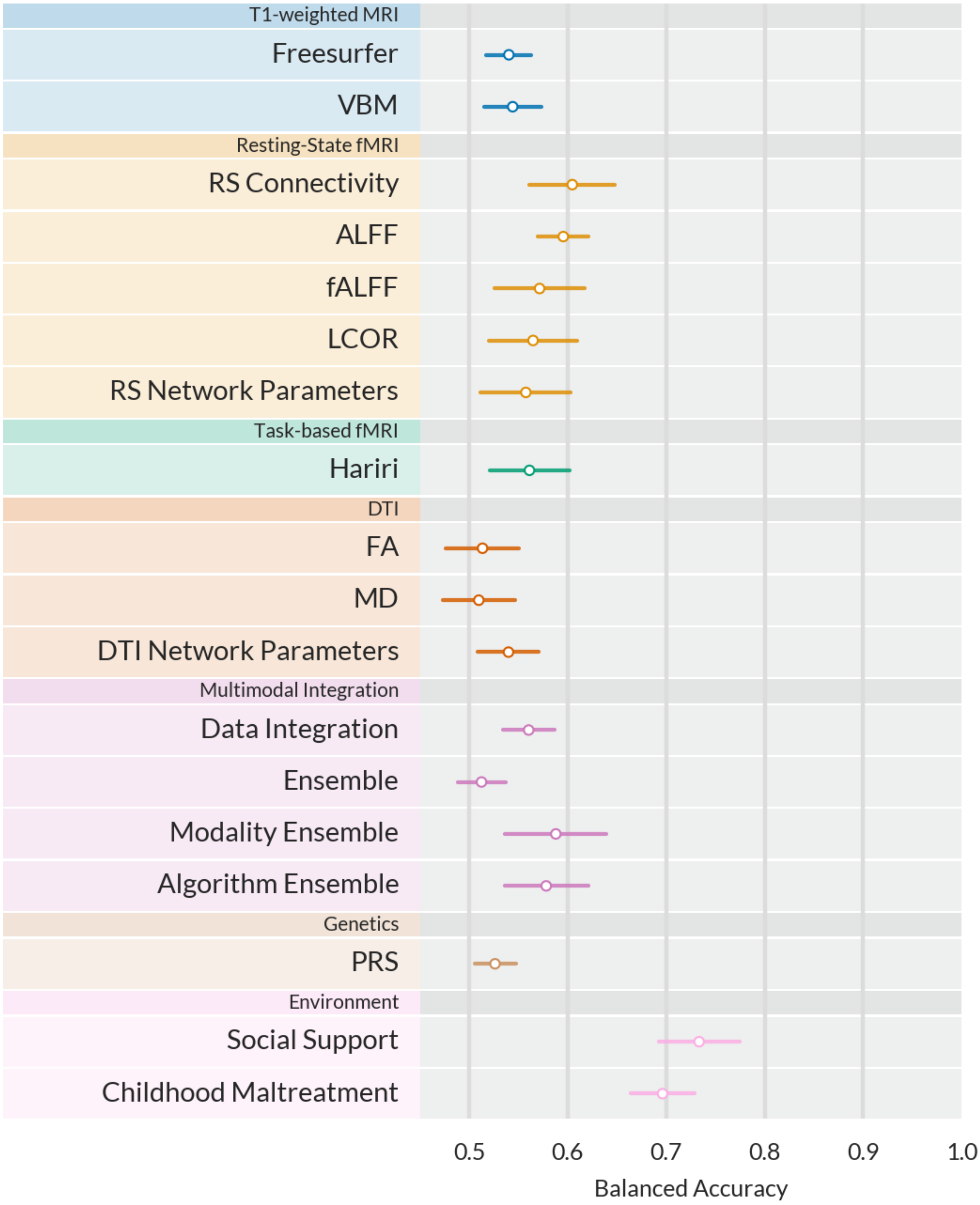


**eFigure 1**. Balanced accuracy for best machine learning pipeline in every modality. Error bars display +-1 standard deviation calculated across the 10 outer cross-validation folds. VBM=Voxel-based morphometry, ALFF=Amplitude of low-frequency fluctuations, fALFF=fractional ALFF, LCOR=Local correlation, FA=Fractional anisotropy, MD=Mean diffusivity, PRS=Polygenic risk score.

**eFigure 2. Classification results for healthy participants versus patients with recurrent MDD**

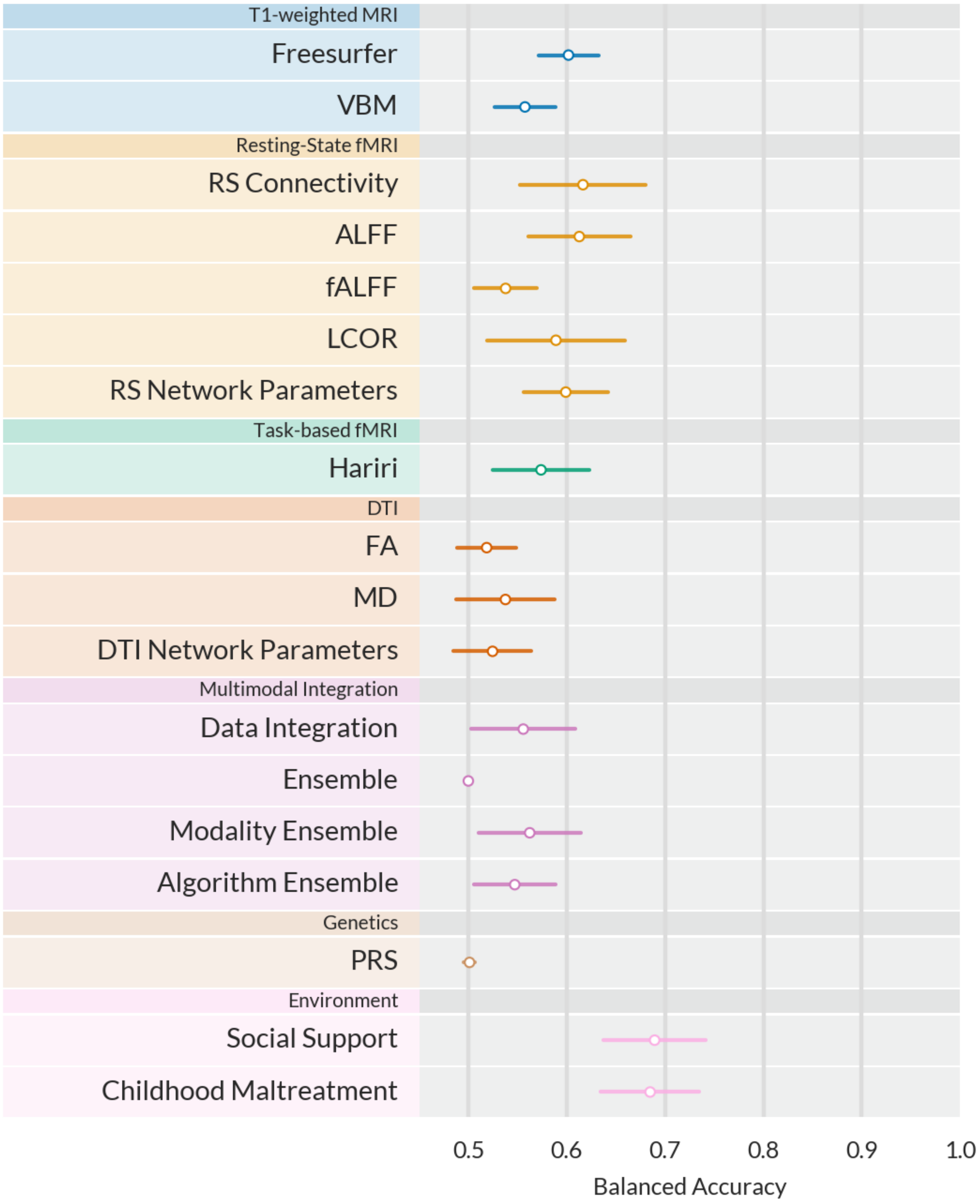


**eFigure 2.** Balanced accuracy for best machine learning pipeline in every modality. Error bars display +-1 standard deviation calculated across the 10 outer cross-validation folds. VBM=Voxel-based morphometry, ALFF=Amplitude of low-frequency fluctuations, fALFF=fractional ALFF, LCOR=Local correlation, FA=Fractional anisotropy, MD=Mean diffusivity, PRS=Polygenic risk score.

**eFigure 3. Classification results for male participants only (HC versus MDD)**

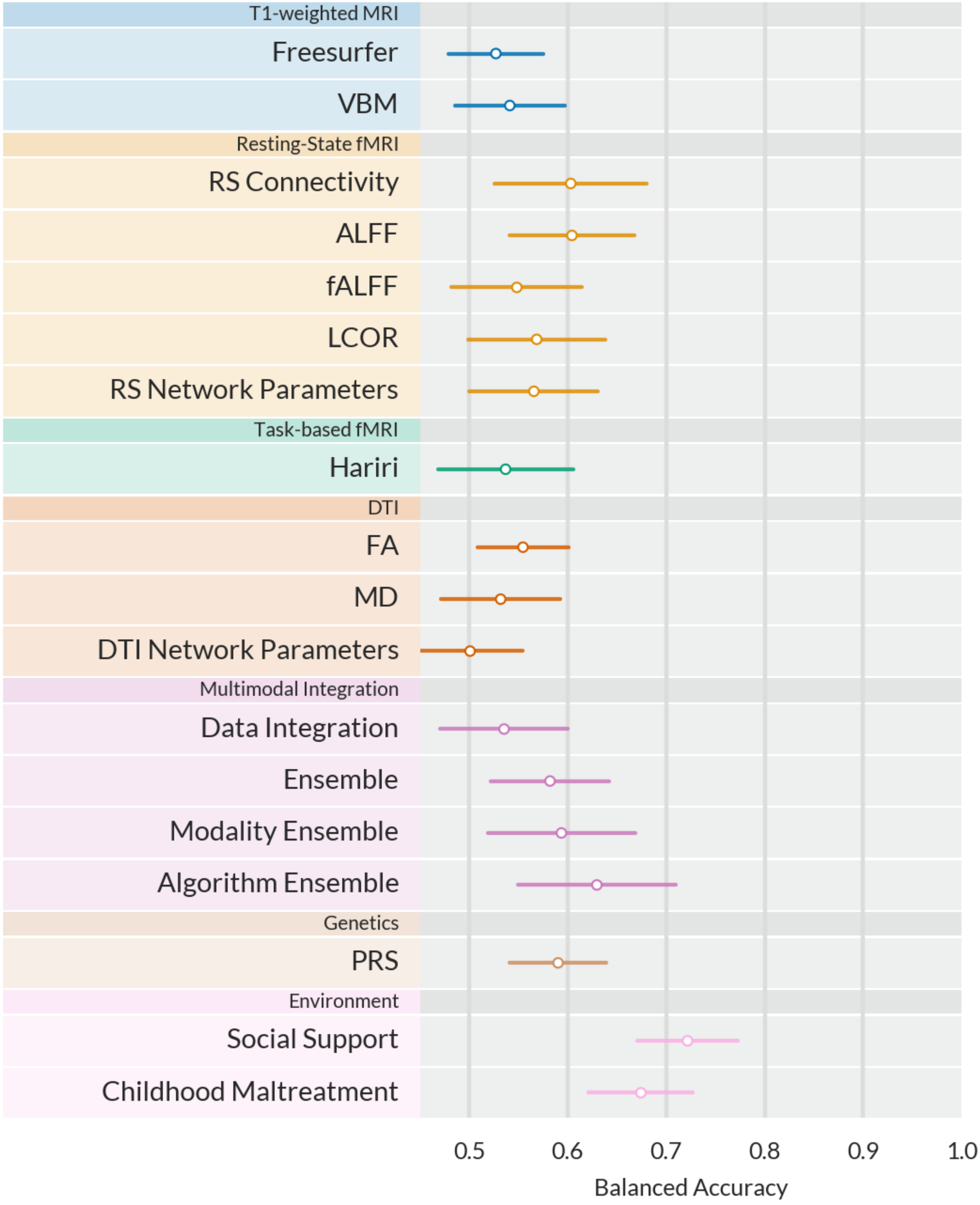


**eFigure 3.** Balanced accuracy for best machine learning pipeline in every modality. Error bars display +-1 standard deviation calculated across the 10 outer cross-validation folds. VBM=Voxel-based morphometry, ALFF=Amplitude of low-frequency fluctuations, fALFF=fractional ALFF, LCOR=Local correlation, FA=Fractional anisotropy, MD=Mean diffusivity, PRS=Polygenic risk score.

**eFigure 4. Classification results for female participants only (HC versus MDD)**

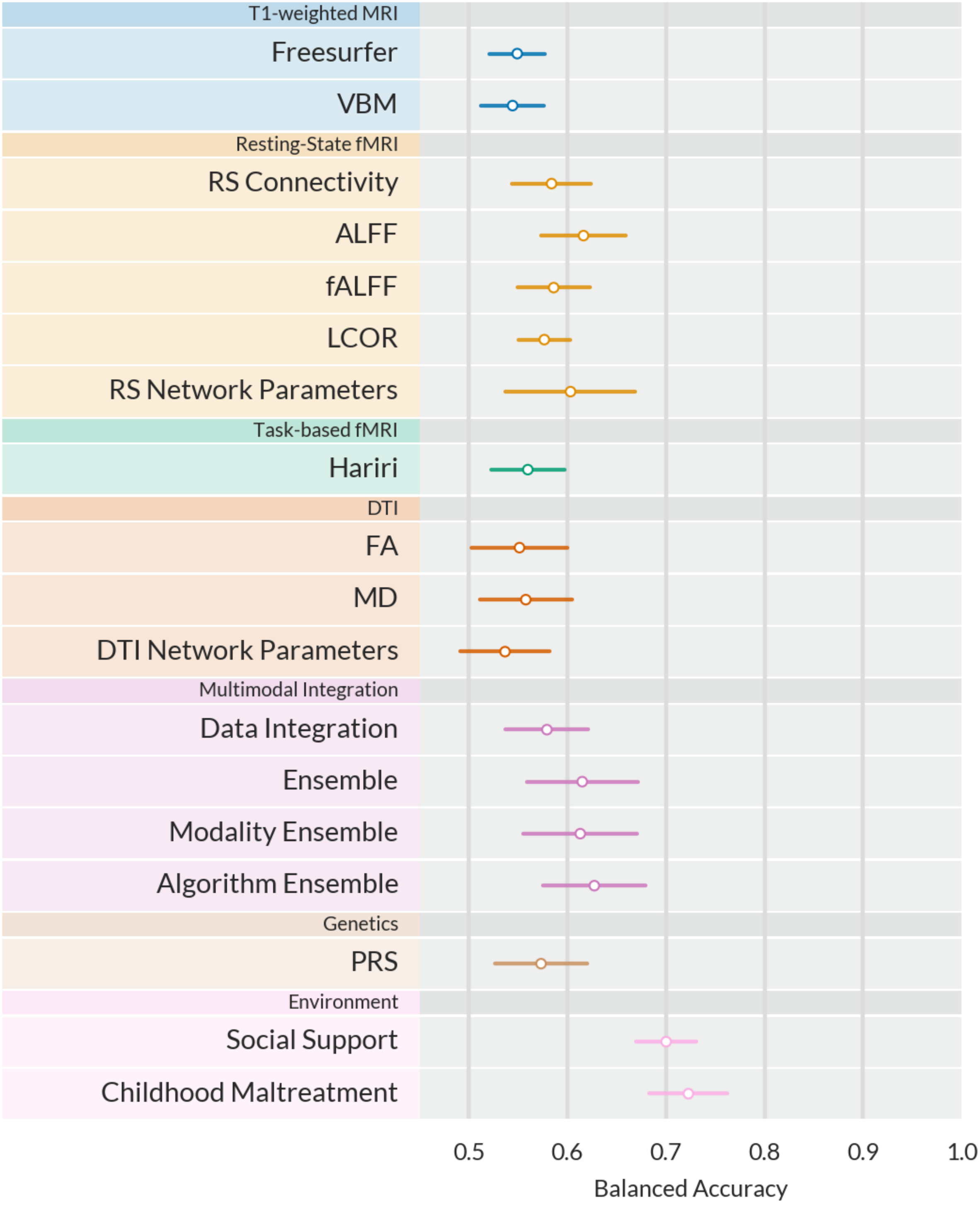


**eFigure 4.** Balanced accuracy for best machine learning pipeline in every modality. Error bars display +-1 standard deviation calculated across the 10 outer cross-validation folds. VBM=Voxel-based morphometry, ALFF=Amplitude of low-frequency fluctuations, fALFF=fractional ALFF, LCOR=Local correlation, FA=Fractional anisotropy, MD=Mean diffusivity, PRS=Polygenic risk score.

**eFigure 5. Classification results for participants within an age range of 24 to 28 years (HC versus MDD)**

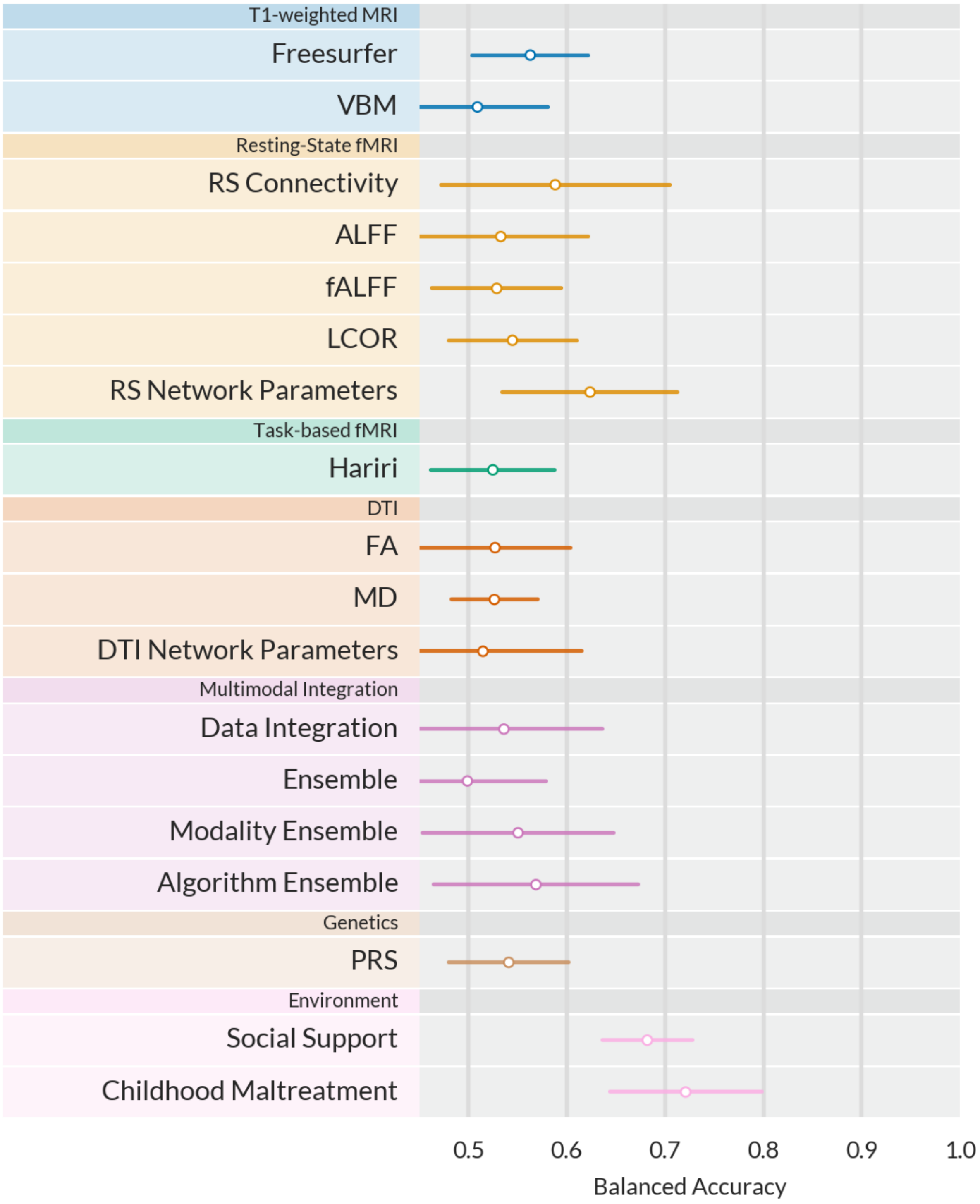


**eFigure 5.** Balanced accuracy for best machine learning pipeline in every modality. Error bars display +-1 standard deviation calculated across the 10 outer cross-validation folds. VBM=Voxel-based morphometry, ALFF=Amplitude of low-frequency fluctuations, fALFF=fractional ALFF, LCOR=Local correlation, FA=Fractional anisotropy, MD=Mean diffusivity, PRS=Polygenic risk score**.**

**eFigure 6. Correlations of model predictions across modalities for HC versus MDD.**

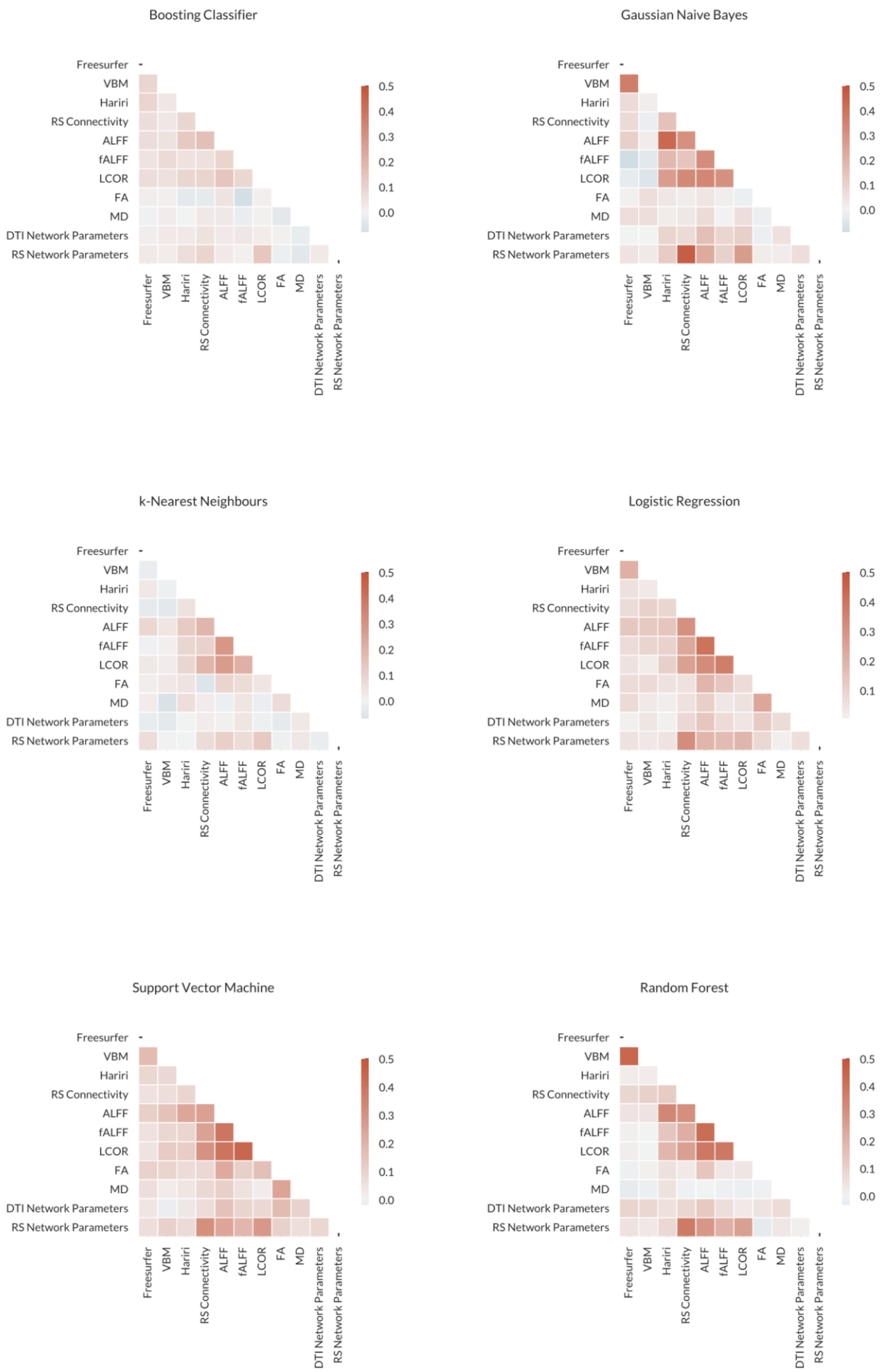


**eFigure 6.** Correlation plots correlating model predictions for all six algorithms across neuroimaging modalities.

**eTable 1. Classification accuracy for multivariate biomarkers based on structural MRI.**

| *eTable 1. Classification accuracy for multivariate biomarkers based on structural MRI.* | | | | | | | | |
|---|---|---|---|---|---|---|---|---|
| **Modality** | **Algorithm** | **BACC** | **ACC** | **Sensitivity** | **Specificity** | **AUC** | **MCC** | **n (HC/MDD)** |
| Freesurfer | Boosting Classifier | 51.9% (2.8%) | 52.6% (2.9%) | 41.1% (8.2%) | 62.8% (8.6%) | 52.9% (3.7%) | 0.04 (0.06) | 920/813 |
| | k-Nearest Neighbours | 51.2% (4.8%) | 51.8% (4.9%) | 42.8% (6.2%) | 59.7% (8.3%) | 52.4% (5.2%) | 0.03 (0.1) | 920/813 |
| | Logistic Regression | 53.7% (3.1%) | 54.5% (2.9%) | 41.0% (9.2%) | 66.4% (7.0%) | 55.7% (3.8%) | 0.08 (0.06) | 920/813 |
| | Gaussian Naive Bayes | 54.5% (2.1%) | 55.1% (2.1%) | 46.4% (5.1%) | 62.7% (4.7%) | 56.0% (3.5%) | 0.09 (0.04) | 920/813 |
| | Random Forest | 53.5% (2.3%) | 54.6% (2.3%) | 34.8% (4.9%) | 72.2% (4.7%) | 55.1% (3.4%) | 0.08 (0.05) | 920/813 |
| | Support Vector Machine | 53.4% (2.7%) | 54.4% (2.5%) | 37.8% (9.6%) | 69.0% (7.4%) | 53.4% (2.7%) | 0.07 (0.06) | 920/813 |
| VBM | Boosting Classifier | 53.2% (3.6%) | 53.6% (3.9%) | 45.5% (4.3%) | 60.8% (8.1%) | 53.9% (3.6%) | 0.07 (0.07) | 924/817 |
| | k-Nearest Neighbours | 50.5% (2.5%) | 51.4% (2.5%) | 35.5% (6.1%) | 65.5% (5.5%) | 50.0% (3.2%) | 0.01 (0.05) | 924/817 |
| | Logistic Regression | 51.4% (3.2%) | 49.7% (3.2%) | 80.0% (8.3%) | 22.8% (8.4%) | 51.4% (4.1%) | 0.04 (0.07) | 924/817 |
| | Gaussian Naive Bayes | 52.5% (3.4%) | 53.6% (2.8%) | 34.4% (23.1%) | 70.6% (18.6%) | 54.8% (2.4%) | 0.05 (0.07) | 924/817 |
| | Random Forest | 55.3% (4.6%) | 56.4% (4.7%) | 37.9% (7.4%) | 72.7% (7.8%) | 57.4% (4.0%) | 0.12 (0.1) | 924/817 |
| | Support Vector Machine | 51.4% (3.3%) | 51.2% (3.6%) | 54.0% (6.7%) | 48.8% (10.8%) | 51.4% (3.3%) | 0.03 (0.07) | 924/817 |
| DTI FA | Boosting Classifier | 49.5% (3.5%) | 50.6% (4.2%) | 37.1% (14.3%) | 61.9% (16.7%) | 50.0% (4.3%) | -0.01 (0.07) | 818/685 |
| | k-Nearest Neighbours | 51.8% (3.5%) | 52.2% (3.5%) | 47.7% (10.6%) | 55.9% (9.4%) | 51.8% (4.4%) | 0.04 (0.07) | 818/685 |
| | Logistic Regression | 51.0% (4.9%) | 52.0% (4.7%) | 39.7% (8.9%) | 62.2% (6.8%) | 53.0% (5.5%) | 0.02 (0.1) | 818/685 |
| | Gaussian Naive Bayes | 48.1% (3.6%) | 46.8% (3.1%) | 62.8% (28.5%) | 33.5% (24.7%) | 48.7% (3.8%) | -0.03 (0.08) | 818/685 |
| | Random Forest | 49.5% (3.4%) | 51.8% (4.0%) | 23.8% (6.6%) | 75.2% (10.7%) | 49.7% (5.6%) | -0.01 (0.08) | 818/685 |

| | | | | | | | | |
|---|---|---|---|---|---|---|---|---|
| | Support Vector Machine | 54.9% (3.0%) | 56.0% (2.9%) | 41.9% (6.2%) | 67.9% (4.4%) | 54.9% (3.0%) | 0.1 (0.06) | 818/685 |
| DTI MD | Boosting Classifier | 49.4% (3.4%) | 50.6% (3.8%) | 36.5% (13.1%) | 62.3% (14.2%) | 47.4% (3.0%) | -0.01 (0.07) | 818/685 |
| | k-Nearest Neighbours | 49.9% (4.8%) | 51.1% (4.8%) | 37.2% (8.5%) | 62.7% (7.8%) | 49.2% (5.8%) | -0.0 (0.1) | 818/685 |
| | Logistic Regression | 50.8% (4.1%) | 52.2% (4.6%) | 35.4% (13.7%) | 66.3% (16.2%) | 51.3% (4.9%) | 0.02 (0.09) | 818/685 |
| | Gaussian Naive Bayes | 51.2% (2.4%) | 52.5% (2.9%) | 36.2% (9.7%) | 66.2% (11.8%) | 49.8% (4.0%) | 0.03 (0.05) | 818/685 |
| | Random Forest | 50.1% (2.6%) | 52.6% (2.9%) | 21.8% (5.5%) | 78.5% (8.0%) | 50.3% (4.9%) | 0.01 (0.07) | 818/685 |
| | Support Vector Machine | 51.8% (3.5%) | 52.8% (3.7%) | 40.3% (7.9%) | 63.3% (9.4%) | 51.8% (3.5%) | 0.04 (0.07) | 818/685 |
| DTI Network Parameters | Boosting Classifier | 52.8% (3.2%) | 53.8% (3.4%) | 40.5% (5.2%) | 65.1% (7.7%) | 54.0% (3.6%) | 0.06 (0.07) | 818/685 |
| | k-Nearest Neighbours | 51.1% (3.3%) | 52.8% (3.3%) | 32.8% (7.2%) | 69.5% (7.2%) | 51.4% (5.1%) | 0.02 (0.07) | 818/685 |
| | Logistic Regression | 54.4% (3.7%) | 55.0% (3.6%) | 48.2% (5.0%) | 60.6% (4.1%) | 57.8% (4.5%) | 0.09 (0.07) | 818/685 |
| | Gaussian Naive Bayes | 54.0% (4.9%) | 54.0% (4.8%) | 53.9% (8.8%) | 54.0% (7.6%) | 56.0% (4.3%) | 0.08 (0.1) | 818/685 |
| | Random Forest | 53.7% (1.8%) | 56.0% (2.1%) | 28.6% (7.1%) | 78.9% (8.3%) | 54.4% (3.8%) | 0.09 (0.05) | 818/685 |
| | Support Vector Machine | 53.3% (2.3%) | 54.6% (1.9%) | 39.3% (8.5%) | 67.4% (5.7%) | 53.3% (2.3%) | 0.07 (0.05) | 818/685 |

Note: HC = Healthy Controls, MDD = Major Depressive Disorder, BACC = Balanced Accuracy, ACC = Accuracy, AUC = Area Under The Receiver Operating Characteristic Curve, MCC = Matthew’s Correlation Coefficient.

**eTable 2. Classification accuracy for multivariate biomarkers based on functional MRI.**

| *eTable 2. Classification accuracy for multivariate biomarkers based on functional MRI.* | | | | | | | | |
|---|---|---|---|---|---|---|---|---|
| **Modality** | **Algorithm** | **BACC** | **ACC** | **Sensitivity** | **Specificity** | **AUC** | **MCC** | **n (HC/MDD)** |
| Face Matching Task | Boosting Classifier | 53.0% (4.1%) | 53.3% (4.0%) | 48.8% (7.5%) | 57.2% (4.8%) | 52.9% (5.6%) | 0.06 (0.08) | 654/580 |
| | k-Nearest Neighbours | 51.5% (5.7%) | 52.5% (5.4%) | 34.7% (12.7%) | 68.3% (5.9%) | 51.9% (5.6%) | 0.03 (0.12) | 654/580 |
| | Logistic Regression | 53.3% (6.0%) | 53.5% (5.9%) | 49.1% (10.6%) | 57.4% (7.8%) | 54.8% (5.8%) | 0.07 (0.12) | 654/580 |
| | Gaussian Naive Bayes | 54.9% (4.7%) | 54.8% (4.7%) | 56.4% (7.9%) | 53.4% (8.2%) | 55.6% (4.9%) | 0.1 (0.09) | 654/580 |
| | Random Forest | 55.6% (4.5%) | 56.5% (4.5%) | 41.9% (6.3%) | 69.4% (5.6%) | 59.1% (5.7%) | 0.12 (0.09) | 654/580 |
| | Support Vector Machine | 54.0% (5.3%) | 54.2% (5.5%) | 50.0% (15.5%) | 58.0% (16.0%) | 54.0% (5.3%) | 0.08 (0.11) | 654/580 |
| RS Connectivity | Boosting Classifier | 51.5% (7.1%) | 51.7% (7.1%) | 48.5% (10.1%) | 54.6% (8.5%) | 54.5% (7.4%) | 0.03 (0.14) | 700/631 |
| | k-Nearest Neighbours | 56.1% (6.3%) | 56.9% (6.1%) | 40.3% (11.5%) | 71.9% (7.2%) | 59.1% (7.3%) | 0.13 (0.13) | 700/631 |
| | Logistic Regression | 59.7% (3.7%) | 60.2% (3.8%) | 49.5% (7.1%) | 69.9% (7.8%) | 62.7% (4.4%) | 0.2 (0.08) | 700/631 |
| | Gaussian Naive Bayes | 59.0% (3.6%) | 59.1% (3.7%) | 56.6% (6.4%) | 61.4% (8.2%) | 60.4% (2.9%) | 0.18 (0.07) | 700/631 |
| | Random Forest | 59.1% (3.1%) | 59.6% (2.9%) | 49.3% (11.0%) | 68.9% (7.2%) | 61.3% (3.1%) | 0.19 (0.06) | 700/631 |
| | Support Vector Machine | 61.5% (3.4%) | 61.8% (3.3%) | 55.3% (7.2%) | 67.7% (5.0%) | 61.5% (3.4%) | 0.23 (0.07) | 700/631 |
| ALFF | Boosting Classifier | 56.6% (4.2%) | 56.8% (4.4%) | 52.8% (7.5%) | 60.5% (10.5%) | 60.4% (6.0%) | 0.13 (0.08) | 701/631 |
| | k-Nearest Neighbours | 60.3% (4.3%) | 60.7% (4.4%) | 53.4% (5.8%) | 67.2% (6.4%) | 63.2% (4.6%) | 0.21 (0.09) | 701/631 |
| | Logistic Regression | 61.0% (4.7%) | 61.3% (4.6%) | 55.8% (8.7%) | 66.2% (6.1%) | 64.3% (4.2%) | 0.22 (0.09) | 701/631 |
| | Gaussian Naive Bayes | 59.1% (4.3%) | 59.2% (4.3%) | 55.8% (7.7%) | 62.4% (8.7%) | 59.7% (4.6%) | 0.18 (0.09) | 701/631 |
| | Random Forest | 59.8% (3.5%) | 60.4% (3.5%) | 49.9% (7.4%) | 69.8% (6.5%) | 63.9% (4.1%) | 0.2 (0.07) | 701/631 |
| | Support Vector Machine | 60.2% (3.4%) | 60.7% (3.3%) | 52.1% (8.2%) | 68.3% (6.0%) | 60.2% (3.4%) | 0.21 (0.07) | 701/631 |
| fALFF | Boosting Classifier | 52.7% (4.4%) | 52.9% (4.3%) | 47.7% (9.9%) | 57.6% (9.2%) | 53.3% (5.7%) | 0.05 (0.09) | 701/631 |

| | | | | | | | | |
|---|---|---|---|---|---|---|---|---|
| | k-Nearest Neighbours | 55.6% (3.2%) | 56.6% (3.2%) | 36.6% (6.7%) | 74.6% (5.7%) | 57.3% (4.7%) | 0.12 (0.07) | 701/631 |
| | Logistic Regression | 58.7% (4.0%) | 59.1% (4.0%) | 51.8% (6.2%) | 65.6% (6.1%) | 61.6% (5.5%) | 0.18 (0.08) | 701/631 |
| | Gaussian Naive Bayes | 54.7% (4.4%) | 54.8% (4.3%) | 52.5% (6.8%) | 56.9% (4.7%) | 54.9% (4.5%) | 0.09 (0.09) | 701/631 |
| | Random Forest | 54.4% (3.0%) | 55.1% (3.0%) | 40.3% (6.4%) | 68.5% (5.3%) | 55.9% (3.5%) | 0.09 (0.06) | 701/631 |
| | Support Vector Machine | 56.5% (3.3%) | 56.9% (3.3%) | 48.8% (5.6%) | 64.2% (4.5%) | 56.5% (3.3%) | 0.13 (0.07) | 701/631 |
| LCOR | Boosting Classifier | 51.5% (4.3%) | 51.9% (4.2%) | 44.4% (8.3%) | 58.6% (5.8%) | 53.2% (3.4%) | 0.03 (0.09) | 701/631 |
| | k-Nearest Neighbours | 55.7% (3.6%) | 56.2% (3.5%) | 45.3% (10.5%) | 66.1% (7.9%) | 57.1% (3.0%) | 0.12 (0.08) | 701/631 |
| | Logistic Regression | 55.8% (2.4%) | 56.2% (2.3%) | 48.6% (4.4%) | 62.9% (3.3%) | 58.4% (3.9%) | 0.12 (0.05) | 701/631 |
| | Gaussian Naive Bayes | 55.6% (4.7%) | 55.7% (4.7%) | 54.0% (6.8%) | 57.2% (6.6%) | 56.0% (4.7%) | 0.11 (0.09) | 701/631 |
| | Random Forest | 53.8% (5.1%) | 54.4% (5.2%) | 42.3% (6.8%) | 65.2% (7.7%) | 56.5% (5.4%) | 0.08 (0.11) | 701/631 |
| | Support Vector Machine | 55.7% (2.2%) | 56.2% (2.1%) | 47.5% (4.8%) | 63.9% (2.6%) | 55.7% (2.2%) | 0.12 (0.04) | 701/631 |
| RS Network Parameters | Boosting Classifier | 53.5% (3.6%) | 54.0% (3.5%) | 44.2% (6.8%) | 62.9% (5.5%) | 55.0% (4.8%) | 0.07 (0.07) | 700/631 |
| | k-Nearest Neighbours | 53.6% (4.6%) | 54.4% (4.5%) | 38.8% (8.0%) | 68.4% (5.9%) | 56.0% (5.6%) | 0.08 (0.1) | 700/631 |
| | Logistic Regression | 56.6% (2.6%) | 57.1% (2.8%) | 47.7% (6.7%) | 65.6% (8.8%) | 57.6% (3.4%) | 0.14 (0.06) | 700/631 |
| | Gaussian Naive Bayes | 56.3% (3.1%) | 56.4% (3.1%) | 54.7% (6.2%) | 57.9% (6.5%) | 57.5% (3.9%) | 0.13 (0.06) | 700/631 |
| | Random Forest | 56.8% (3.0%) | 57.6% (3.1%) | 40.6% (6.5%) | 73.0% (7.0%) | 58.3% (4.7%) | 0.15 (0.06) | 700/631 |
| | Support Vector Machine | 56.9% (3.0%) | 57.4% (3.0%) | 46.8% (7.9%) | 67.0% (7.3%) | 56.9% (3.0%) | 0.14 (0.06) | 700/631 |

Note: HC = Healthy Controls, MDD = Major Depressive Disorder, BACC = Balanced Accuracy, ACC = Accuracy, AUC = Area Under The Receiver Operating Characteristic Curve, MCC = Matthew's Correlation Coefficient.

**eTable 3. Classification accuracy for multivariate biomarkers using neuroimaging modality integration.**

| *eTable 3. Classification accuracy for multivariate biomarkers using neuroimaging modality integration.* | | | | | | | | | |
|---|---|---|---|---|---|---|---|---|---|
| **Modality Integration** | **Modality** | **Algorithm** | **BACC** | **ACC** | **Sensitivity** | **Specificity** | **AUC** | **MCC** | **n (HC/MDD)** |
| PCA-based | all | Boosting Classifier | 54.1% (4.1%) | 54.9% (4.4%) | 45.4% (11.6%) | 62.7% (12.6%) | 55.2% (5.4%) | 0.09 (0.08) | 567/471 |
| | | k-Nearest Neighbours | 50.3% (3.9%) | 51.7% (3.9%) | 35.0% (8.4%) | 65.6% (8.0%) | 50.0% (5.1%) | 0.01 (0.08) | 567/471 |
| | | Logistic Regression | 51.1% (3.0%) | 52.1% (4.5%) | 40.0% (44.2%) | 62.3% (43.0%) | 51.4% (5.2%) | 0.05 (0.12) | 567/471 |
| | | Gaussian Naive Bayes | 51.3% (3.6%) | 54.6% (3.2%) | 15.9% (24.7%) | 86.7% (20.8%) | 50.4% (4.6%) | 0.04 (0.13) | 567/471 |
| | | Random Forest | 57.2% (4.4%) | 59.8% (4.4%) | 29.1% (8.3%) | 85.4% (6.1%) | 62.0% (4.8%) | 0.18 (0.11) | 567/471 |
| | | Support Vector Machine | 50.1% (4.0%) | 49.2% (5.3%) | 60.2% (46.5%) | 40.0% (45.0%) | 50.1% (4.0%) | 0.01 (0.15) | 567/471 |
| Voting | all | all | 61.1% (4.4%) | 63.3% (4.4%) | 37.6% (6.7%) | 84.6% (5.0%) | 65.3% (4.2%) | 0.26 (0.1) | 567/471 |
| Voting | all | Boosting Classifier | 55.5% (4.2%) | 57.4% (4.3%) | 34.8% (6.9%) | 34.8% (6.9%) | 57.4% (5.3%) | 0.12 (0.1) | 567/471 |
| | | k-Nearest Neighbours | 56.3% (3.2%) | 58.9% (3.3%) | 28.7% (3.3%) | 28.7% (3.3%) | 62.2% (5.5%) | 0.16 (0.08) | 567/471 |
| | | Logistic Regression | 60.9% (4.2%) | 62.1% (4.2%) | 47.2% (8.1%) | 47.2% (8.1%) | 63.7% (4.0%) | 0.23 (0.09) | 567/471 |
| | | Gaussian Naive Bayes | 60.6% (5.8%) | 61.4% (5.9%) | 51.8% (7.2%) | 51.8% (7.2%) | 62.7% (4.1%) | 0.22 (0.12) | 567/471 |
| | | Random Forest | 59.5% (3.5%) | 62.3% (3.3%) | 29.5% (6.3%) | 29.5% (6.3%) | 63.3% (4.2%) | 0.24 (0.08) | 567/471 |
| | | Support Vector Machine | 60.3% (3.8%) | 62.0% (3.6%) | 42.1% (8.0%) | 42.1% (8.0%) | 64.9% (3.6%) | 0.22 (0.08) | 567/471 |
| Voting | Freesurfer | all | 53.1% (4.4%) | 55.0% (4.3%) | 31.9% (6.5%) | 31.9% (6.5%) | 54.3% (5.7%) | 0.07 (0.1) | 567/471 |
| | VBM | | 54.2% (4.6%) | 56.0% (4.5%) | 35.1% (8.2%) | 35.1% (8.2%) | 54.5% (6.0%) | 0.09 (0.1) | 567/471 |
| | DTI FA | | 52.0% (5.4%) | 54.5% (5.4%) | 25.1% (7.3%) | 25.1% (7.3%) | 52.4% (7.5%) | 0.05 (0.13) | 567/471 |
| | DTI MD | | 50.6% (4.1%) | 53.6% (4.1%) | 18.5% (4.3%) | 18.5% (4.3%) | 52.1% (6.3%) | 0.02 (0.11) | 567/471 |

| | | | | | | | | | |
|---|---|---|---|---|---|---|---|---|---|
| | DTI Network Parameters | | 55.7% (3.8%) | 58.1% (3.6%) | 29.5% (6.9%) | 29.5% (6.9%) | 56.3% (5.6%) | 0.13 (0.09) | 567/471 |
| | Face Matching Task | | 55.6% (5.7%) | 57.2% (5.7%) | 37.6% (7.4%) | 37.6% (7.4%) | 57.2% (6.0%) | 0.12 (0.12) | 567/471 |
| | RS Connectivity | | 60.2% (4.6%) | 61.5% (4.7%) | 46.1% (6.6%) | 46.1% (6.6%) | 62.2% (5.6%) | 0.21 (0.1) | 567/471 |
| | ALFF | | 62.0% (4.8%) | 63.3% (4.7%) | 48.2% (7.5%) | 48.2% (7.5%) | 64.7% (5.5%) | 0.25 (0.1) | 567/471 |
| | fALFF | | 55.9% (4.5%) | 57.4% (4.2%) | 39.3% (9.0%) | 39.3% (9.0%) | 58.5% (5.5%) | 0.12 (0.09) | 567/471 |
| | LCOR | | 54.7% (3.4%) | 56.3% (3.4%) | 38.2% (6.1%) | 38.2% (6.1%) | 58.2% (3.4%) | 0.1 (0.07) | 567/471 |
| | RS Network Parameters | | 57.5% (4.9%) | 59.4% (4.8%) | 37.8% (6.9%) | 37.8% (6.9%) | 60.7% (5.2%) | 0.16 (0.11) | 567/471 |

Note: HC = Healthy Controls, MDD = Major Depressive Disorder, BACC = Balanced Accuracy, ACC = Accuracy, AUC = Area Under The Receiver Operating Characteristic Curve, MCC = Matthew's Correlation Coefficient.

**eTable 4. Associations of symptom severity and misclassification frequency.**

| *eTable 4. Associations of symptom severity and misclassification frequency.* | | | | | | |
|---|---|---|---|---|---|---|
| | **MDD** | | | **HC** | | |
| | **N** | **r** | ***p*** | **N** | **r** | ***p*** |
| **Age** | 631 | -0.057 | 0.156 | 700 | 0.011 | 0.774 |
| **BDI** | 621 | -0.152 | <0.001 | | | |
| **HAMD** | 628 | -0.197 | <0.001 | | | |
| **GAF** | 620 | 0.171 | <0.001 | 690 | -0.102 | 0.007 |
| **Number of Hospitalizations** | 622 | -0.102 | 0.011 | | | |

Note: BDI = Beck's Depression Inventory, HAMD = Hamilton Depression Rating Scale, GAF = Global Assessment of Functioning.

**eTable 5. Classification accuracy based on structural MRI for HC vs acute MDD.**

| *eTable 5. Classification accuracy for multivariate biomarkers based on structural MRI for HC vs acute MDD.* | | | | | | | | |
|---|---|---|---|---|---|---|---|---|
| **Modality** | **Algorithm** | **BACC** | **ACC** | **Sensitivity** | **Specificity** | **AUC** | **MCC** | **n (HC/MDD)** |
| Freesurfer | Boosting Classifier | 52.7% (2.1%) | 58.7% (2.1%) | 27.1% (6.7%) | 78.3% (5.5%) | 54.7% (2.6%) | 0.06 (0.05) | 920/568 |
| | k-Nearest Neighbours | 51.4% (3.4%) | 57.5% (3.7%) | 25.7% (9.5%) | 77.1% (8.6%) | 55.4% (5.0%) | 0.03 (0.07) | 920/568 |
| | Logistic Regression | 53.5% (4.5%) | 55.2% (4.2%) | 46.0% (9.1%) | 61.0% (6.1%) | 54.0% (5.2%) | 0.07 (0.09) | 920/568 |
| | Gaussian Naive Bayes | 54.0% (2.4%) | 58.0% (2.2%) | 37.3% (8.0%) | 70.8% (5.9%) | 55.4% (4.2%) | 0.08 (0.05) | 920/568 |
| | Random Forest | 52.5% (1.3%) | 61.7% (1.3%) | 13.6% (3.2%) | 91.4% (2.6%) | 57.2% (3.4%) | 0.08 (0.04) | 920/568 |
| | Support Vector Machine | 51.4% (3.2%) | 57.1% (4.1%) | 27.0% (11.1%) | 75.8% (11.2%) | 51.4% (3.2%) | 0.03 (0.07) | 920/568 |
| VBM | Boosting Classifier | 54.4% (3.1%) | 58.7% (3.0%) | 36.4% (5.0%) | 72.4% (3.9%) | 56.4% (3.6%) | 0.09 (0.06) | 924/571 |
| | k-Nearest Neighbours | 49.1% (1.7%) | 55.5% (1.5%) | 21.9% (7.3%) | 76.3% (5.4%) | 49.2% (5.6%) | -0.02 (0.04) | 924/571 |
| | Logistic Regression | 51.1% (3.4%) | 45.5% (6.1%) | 74.9% (15.8%) | 27.4% (18.1%) | 52.5% (2.9%) | 0.03 (0.08) | 924/571 |
| | Gaussian Naive Bayes | 52.2% (2.7%) | 58.9% (3.0%) | 23.7% (21.4%) | 80.8% (17.3%) | 53.3% (4.4%) | 0.05 (0.07) | 924/571 |
| | Random Forest | 52.1% (3.2%) | 61.0% (3.0%) | 14.4% (4.6%) | 89.8% (3.6%) | 59.0% (2.8%) | 0.06 (0.09) | 924/571 |
| | Support Vector Machine | 49.7% (1.7%) | 49.3% (11.0%) | 51.4% (45.6%) | 48.0% (45.7%) | 49.7% (1.7%) | -0.01 (0.07) | 924/571 |
| DTI FA | Boosting Classifier | 51.0% (4.5%) | 57.7% (4.6%) | 25.0% (9.0%) | 77.0% (7.8%) | 49.1% (6.0%) | 0.02 (0.11) | 818/484 |
| | k-Nearest Neighbours | 48.8% (2.4%) | 55.4% (3.4%) | 23.1% (6.9%) | 74.5% (8.1%) | 48.7% (3.5%) | -0.03 (0.05) | 818/484 |
| | Logistic Regression | 49.1% (3.7%) | 53.9% (3.6%) | 30.6% (9.1%) | 67.7% (7.0%) | 49.6% (4.0%) | -0.02 (0.08) | 818/484 |
| | Gaussian Naive Bayes | 48.7% (3.6%) | 55.6% (3.8%) | 21.7% (11.4%) | 75.7% (9.8%) | 48.2% (6.7%) | -0.04 (0.08) | 818/484 |
| | Random Forest | 49.7% (1.4%) | 61.9% (1.3%) | 2.1% (2.6%) | 97.3% (2.0%) | 49.4% (5.0%) | -0.02 (0.07) | 818/484 |

| | | | | | | | | |
|---|---|---|---|---|---|---|---|---|
| | Support Vector Machine | 51.4% (3.9%) | 56.1% (3.9%) | 32.8% (11.7%) | 69.9% (9.2%) | 51.4% (3.9%) | 0.03 (0.08) | 818/484 |
| DTI MD | Boosting Classifier | 48.5% (3.4%) | 55.5% (3.9%) | 21.3% (11.3%) | 75.7% (10.1%) | 46.4% (4.1%) | -0.04 (0.07) | 818/484 |
| | k-Nearest Neighbours | 51.0% (3.9%) | 59.7% (3.6%) | 17.1% (7.5%) | 84.8% (5.2%) | 51.4% (5.2%) | 0.02 (0.11) | 818/484 |
| | Logistic Regression | 48.4% (4.5%) | 55.1% (5.7%) | 22.2% (12.8%) | 74.6% (13.7%) | 50.1% (5.2%) | -0.04 (0.1) | 818/484 |
| | Gaussian Naive Bayes | 48.6% (3.3%) | 56.4% (4.6%) | 18.2% (11.0%) | 79.0% (11.8%) | 49.3% (4.9%) | -0.03 (0.08) | 818/484 |
| | Random Forest | 48.9% (1.6%) | 60.7% (2.1%) | 2.7% (2.2%) | 95.1% (4.1%) | 46.9% (5.4%) | -0.04 (0.08) | 818/484 |
| | Support Vector Machine | 51.0% (4.1%) | 56.6% (5.4%) | 29.1% (8.1%) | 72.9% (11.3%) | 51.0% (4.1%) | 0.03 (0.09) | 818/484 |
| DTI Network Parameters | Boosting Classifier | 51.9% (3.6%) | 57.4% (3.0%) | 30.7% (14.3%) | 73.1% (10.0%) | 51.8% (4.7%) | 0.04 (0.08) | 818/484 |
| | k-Nearest Neighbours | 49.3% (2.4%) | 58.1% (1.9%) | 14.8% (7.1%) | 83.7% (4.5%) | 50.5% (5.5%) | -0.02 (0.07) | 818/484 |
| | Logistic Regression | 54.0% (3.3%) | 58.7% (3.1%) | 35.7% (7.6%) | 72.2% (5.5%) | 55.3% (3.4%) | 0.08 (0.07) | 818/484 |
| | Gaussian Naive Bayes | 52.0% (5.3%) | 53.4% (5.2%) | 46.8% (15.1%) | 57.2% (11.7%) | 53.9% (5.3%) | 0.04 (0.11) | 818/484 |
| | Random Forest | 51.7% (2.7%) | 61.8% (2.8%) | 12.2% (2.9%) | 91.2% (3.3%) | 53.0% (4.7%) | 0.06 (0.09) | 818/484 |
| | Support Vector Machine | 52.9% (4.7%) | 59.4% (4.3%) | 27.7% (6.8%) | 78.1% (4.1%) | 52.9% (4.7%) | 0.07 (0.11) | 818/484 |

Note: HC = Healthy Controls, MDD = Major Depressive Disorder, BACC = Balanced Accuracy, ACC = Accuracy, AUC = Area Under The Receiver Operating Characteristic Curve, MCC = Matthew's Correlation Coefficient.

**eTable 6. Classification accuracy based on functional MRI for HC vs acute MDD.**

| *eTable 6. Classification accuracy for multivariate biomarkers based on functional MRI for HC vs acute MDD.* | | | | | | | | |
|---|---|---|---|---|---|---|---|---|
| **Modality** | **Algorithm** | **BACC** | **ACC** | **Sensitivity** | **Specificity** | **AUC** | **MCC** | **n (HC/MDD)** |
| Face Matching Task | Boosting Classifier | 53.2% (3.7%) | 57.7% (3.8%) | 34.3% (5.8%) | 72.0% (5.6%) | 56.1% (3.2%) | 0.07 (0.08) | 654/402 |
| | k-Nearest Neighbours | 51.1% (2.7%) | 58.1% (2.9%) | 21.9% (8.7%) | 80.4% (7.4%) | 53.4% (5.1%) | 0.03 (0.07) | 654/402 |
| | Logistic Regression | 56.1% (4.3%) | 59.7% (3.7%) | 41.0% (9.2%) | 71.2% (5.1%) | 59.9% (4.2%) | 0.13 (0.09) | 654/402 |
| | Gaussian Naive Bayes | 55.0% (5.1%) | 56.4% (5.0%) | 49.1% (11.8%) | 60.9% (9.4%) | 55.4% (4.4%) | 0.1 (0.1) | 654/402 |
| | Random Forest | 51.6% (2.0%) | 61.5% (1.6%) | 10.5% (6.5%) | 92.8% (4.0%) | 57.7% (4.9%) | 0.06 (0.07) | 654/402 |
| | Support Vector Machine | 53.4% (5.5%) | 56.8% (6.2%) | 39.0% (11.5%) | 67.7% (12.1%) | 53.4% (5.5%) | 0.07 (0.11) | 654/402 |
| RS Connectivity | Boosting Classifier | 53.8% (5.0%) | 57.0% (4.9%) | 40.1% (7.5%) | 67.6% (5.9%) | 55.8% (5.8%) | 0.08 (0.1) | 700/439 |
| | k-Nearest Neighbours | 55.7% (3.8%) | 62.4% (3.8%) | 26.6% (5.9%) | 84.9% (5.0%) | 58.7% (5.4%) | 0.14 (0.09) | 700/439 |
| | Logistic Regression | 58.7% (5.7%) | 61.5% (5.4%) | 46.7% (10.4%) | 70.7% (7.1%) | 62.2% (6.0%) | 0.18 (0.11) | 700/439 |
| | Gaussian Naive Bayes | 59.4% (3.9%) | 60.6% (3.4%) | 54.4% (10.6%) | 64.4% (7.1%) | 61.6% (4.5%) | 0.19 (0.08) | 700/439 |
| | Random Forest | 54.4% (3.1%) | 61.5% (3.0%) | 23.5% (6.0%) | 85.3% (4.8%) | 61.3% (5.2%) | 0.11 (0.08) | 700/439 |
| | Support Vector Machine | 60.5% (4.6%) | 64.7% (5.0%) | 41.9% (5.7%) | 79.0% (7.5%) | 60.5% (4.6%) | 0.23 (0.1) | 700/439 |
| ALFF | Boosting Classifier | 54.9% (3.6%) | 59.1% (3.5%) | 36.4% (7.3%) | 73.3% (5.5%) | 58.5% (4.0%) | 0.1 (0.08) | 701/439 |
| | k-Nearest Neighbours | 55.9% (5.1%) | 60.5% (4.9%) | 35.7% (8.1%) | 76.0% (5.7%) | 59.9% (5.0%) | 0.13 (0.11) | 701/439 |
| | Logistic Regression | 59.6% (2.7%) | 61.2% (2.6%) | 52.4% (9.7%) | 66.7% (7.5%) | 63.7% (4.5%) | 0.19 (0.05) | 701/439 |
| | Gaussian Naive Bayes | 58.5% (6.0%) | 59.9% (5.3%) | 52.6% (12.4%) | 64.5% (7.6%) | 59.1% (5.8%) | 0.17 (0.12) | 701/439 |
| | Random Forest | 58.0% (2.9%) | 63.6% (3.4%) | 33.9% (4.2%) | 82.2% (5.7%) | 63.3% (4.8%) | 0.19 (0.07) | 701/439 |
| | Support Vector Machine | 57.4% (2.9%) | 61.5% (3.0%) | 39.4% (4.9%) | 75.3% (4.7%) | 57.4% (2.9%) | 0.16 (0.06) | 701/439 |
| fALFF | Boosting Classifier | 52.6% (4.5%) | 56.9% (5.3%) | 33.9% (8.2%) | 71.3% (10.2%) | 56.2% (5.2%) | 0.06 (0.1) | 701/439 |

| | | | | | | | | |
|---|---|---|---|---|---|---|---|---|
| | k-Nearest Neighbours | 53.7% (3.3%) | 62.2% (3.4%) | 16.8% (6.9%) | 90.6% (6.6%) | 57.8% (5.0%) | 0.12 (0.12) | 701/439 |
| | Logistic Regression | 57.2% (4.8%) | 60.9% (4.9%) | 41.0% (10.1%) | 73.3% (8.8%) | 60.5% (6.2%) | 0.15 (0.1) | 701/439 |
| | Gaussian Naive Bayes | 54.5% (5.3%) | 55.0% (5.1%) | 52.2% (8.2%) | 56.8% (5.9%) | 54.8% (6.1%) | 0.09 (0.1) | 701/439 |
| | Random Forest | 50.8% (2.4%) | 60.2% (2.7%) | 10.2% (3.9%) | 91.4% (4.5%) | 57.5% (5.2%) | 0.03 (0.07) | 701/439 |
| | Support Vector Machine | 54.7% (3.9%) | 59.6% (3.3%) | 33.3% (9.0%) | 76.2% (5.1%) | 54.7% (3.9%) | 0.1 (0.08) | 701/439 |
| LCOR | Boosting Classifier | 51.1% (3.8%) | 55.8% (2.8%) | 30.5% (9.5%) | 71.6% (4.1%) | 49.9% (7.2%) | 0.02 (0.08) | 701/439 |
| | k-Nearest Neighbours | 53.3% (4.6%) | 58.5% (4.0%) | 30.8% (9.6%) | 75.9% (5.4%) | 57.3% (6.0%) | 0.07 (0.1) | 701/439 |
| | Logistic Regression | 56.5% (4.8%) | 60.4% (4.6%) | 39.7% (8.3%) | 73.3% (6.1%) | 58.2% (6.0%) | 0.14 (0.1) | 701/439 |
| | Gaussian Naive Bayes | 55.9% (6.3%) | 56.6% (6.1%) | 52.9% (9.1%) | 58.9% (7.5%) | 55.9% (6.4%) | 0.12 (0.12) | 701/439 |
| | Random Forest | 50.8% (3.9%) | 58.9% (3.6%) | 16.0% (8.0%) | 85.7% (5.0%) | 56.9% (8.3%) | 0.02 (0.11) | 701/439 |
| | Support Vector Machine | 53.9% (5.3%) | 58.2% (5.2%) | 35.3% (8.6%) | 72.5% (6.3%) | 53.9% (5.3%) | 0.08 (0.12) | 701/439 |
| RS Network Parameters | Boosting Classifier | 54.9% (4.6%) | 59.1% (4.2%) | 36.7% (7.8%) | 73.1% (4.4%) | 55.5% (6.0%) | 0.1 (0.09) | 700/439 |
| | k-Nearest Neighbours | 52.0% (3.8%) | 58.0% (4.8%) | 25.5% (7.2%) | 78.4% (10.0%) | 53.9% (5.8%) | 0.05 (0.1) | 700/439 |
| | Logistic Regression | 55.5% (3.2%) | 59.9% (2.5%) | 36.2% (10.7%) | 74.7% (6.8%) | 57.2% (3.5%) | 0.12 (0.07) | 700/439 |
| | Gaussian Naive Bayes | 55.3% (7.1%) | 57.4% (7.0%) | 46.2% (10.8%) | 64.4% (9.1%) | 57.7% (6.6%) | 0.11 (0.14) | 700/439 |
| | Random Forest | 55.8% (4.8%) | 63.3% (4.5%) | 22.8% (6.9%) | 88.7% (3.9%) | 58.0% (6.7%) | 0.15 (0.13) | 700/439 |
| | Support Vector Machine | 54.8% (4.1%) | 60.8% (3.7%) | 28.5% (10.0%) | 81.1% (6.8%) | 54.8% (4.1%) | 0.11 (0.1) | 700/439 |

Note: HC = Healthy Controls, MDD = Major Depressive Disorder, BACC = Balanced Accuracy, ACC = Accuracy, AUC = Area Under The Receiver Operating Characteristic Curve, MCC = Matthew’s Correlation Coefficient.

### eTable 7. Classification accuracy using neuroimaging modality integration for HC vs acute MDD.

| *eTable 7. Classification accuracy for multivariate biomarkers using neuroimaging modality integration for HC vs acute MDD.* | | | | | | | | | |
|---|---|---|---|---|---|---|---|---|---|
| **Modality Integration** | **Modality** | **Algorithm** | **BACC** | **ACC** | **Sensitivity** | **Specificity** | **AUC** | **MCC** | **n (HC/MDD)** |
| PCA-based | all | Boosting Classifier | 56.0% (2.7%) | 62.2% (3.3%) | 32.7% (11.1%) | 79.4% (9.7%) | 58.2% (4.5%) | 0.14 (0.07) | 567/330 |
| | | k-Nearest Neighbours | 49.8% (3.8%) | 56.7% (3.6%) | 23.6% (8.4%) | 76.0% (6.2%) | 50.6% (6.8%) | -0.01 (0.09) | 567/330 |
| | | Logistic Regression | 49.3% (2.3%) | 55.0% (9.6%) | 27.6% (37.7%) | 71.0% (36.7%) | 49.2% (3.0%) | -0.01 (0.08) | 567/330 |
| | | Gaussian Naive Bayes | 50.6% (4.7%) | 60.8% (3.8%) | 12.1% (10.8%) | 89.1% (5.6%) | 49.5% (7.9%) | 0.0 (0.13) | 567/330 |
| | | Random Forest | 50.1% (2.2%) | 62.4% (3.2%) | 3.3% (3.0%) | 96.8% (5.9%) | 57.6% (10.8%) | 0.03 (0.11) | 567/330 |
| | | Support Vector Machine | 49.0% (2.0%) | 44.5% (12.7%) | 66.1% (46.0%) | 31.9% (46.7%) | 49.0% (2.0%) | -0.04 (0.1) | 567/330 |
| Voting | all | all | 51.3% (2.6%) | 63.4% (2.0%) | 5.2% (5.0%) | 97.4% (1.9%) | 64.7% (5.3%) | 0.05 (0.12) | 567/330 |
| Voting | all | Boosting Classifier | 52.0% (2.8%) | 63.0% (2.4%) | 10.3% (5.2%) | 93.7% (2.1%) | 60.3% (3.5%) | 0.07 (0.1) | 567/330 |
| | | k-Nearest Neighbours | 51.9% (3.1%) | 64.1% (2.3%) | 5.8% (6.0%) | 98.1% (1.8%) | 57.1% (7.8%) | 0.09 (0.12) | 567/330 |
| | | Logistic Regression | 58.8% (5.4%) | 65.4% (5.8%) | 33.6% (5.6%) | 84.0% (7.6%) | 64.4% (5.0%) | 0.21 (0.13) | 567/330 |
| | | Gaussian Naive Bayes | 58.4% (5.6%) | 64.2% (5.4%) | 36.4% (8.5%) | 80.4% (6.6%) | 60.2% (5.9%) | 0.19 (0.12) | 567/330 |
| | | Random Forest | 50.3% (1.0%) | 63.2% (0.7%) | 1.2% (2.1%) | 99.3% (0.9%) | 59.4% (6.1%) | 0.01 (0.07) | 567/330 |
| | | Support Vector Machine | 56.8% (2.7%) | 66.2% (2.5%) | 21.2% (6.1%) | 92.4% (4.1%) | 61.3% (6.8%) | 0.2 (0.07) | 567/330 |
| Voting | Freesurfer | all | 51.6% (3.4%) | 60.9% (2.8%) | 16.7% (7.2%) | 86.6% (3.6%) | 53.5% (6.2%) | 0.04 (0.09) | 567/330 |
| | VBM | | 51.8% (2.3%) | 62.0% (2.3%) | 13.0% (4.5%) | 90.5% (3.7%) | 52.1% (4.2%) | 0.06 (0.08) | 567/330 |
| | DTI FA | | 49.8% (1.9%) | 61.3% (2.3%) | 6.4% (1.7%) | 93.3% (3.3%) | 51.0% (7.3%) | 0.0 (0.07) | 567/330 |
| | DTI MD | | 49.5% (3.0%) | 61.6% (3.1%) | 3.6% (3.7%) | 95.4% (4.1%) | 47.8% (4.1%) | -0.02 (0.13) | 567/330 |

| | | | | | | | | | |
|---|---|---|---|---|---|---|---|---|---|
| | DTI Network Parameters | | 51.0% (4.1%) | 61.1% (3.7%) | 12.7% (6.0%) | 89.3% (3.2%) | 53.7% (3.2%) | 0.03 (0.12) | 567/330 |
| | Face Matching Task | | 53.9% (3.4%) | 63.4% (2.7%) | 17.9% (7.5%) | 89.9% (3.5%) | 57.2% (6.4%) | 0.11 (0.09) | 567/330 |
| | RS Connectivity | | 57.8% (4.5%) | 65.3% (3.8%) | 29.4% (7.7%) | 86.2% (3.6%) | 62.6% (8.0%) | 0.19 (0.11) | 567/330 |
| | ALFF | | 57.8% (4.9%) | 64.8% (4.2%) | 31.5% (8.6%) | 84.1% (3.8%) | 64.3% (5.9%) | 0.18 (0.11) | 567/330 |
| | fALFF | | 54.6% (3.4%) | 63.8% (3.3%) | 20.0% (5.9%) | 89.2% (4.2%) | 59.0% (3.1%) | 0.13 (0.1) | 567/330 |
| | LCOR | | 52.8% (5.1%) | 61.3% (4.5%) | 20.6% (9.9%) | 85.0% (5.6%) | 57.6% (6.2%) | 0.07 (0.13) | 567/330 |
| | RS Network Parameters | | 56.3% (2.5%) | 65.1% (2.1%) | 23.0% (4.8%) | 89.6% (1.9%) | 59.3% (5.3%) | 0.17 (0.06) | 567/330 |

Note: HC = Healthy Controls, MDD = Major Depressive Disorder, BACC = Balanced Accuracy, ACC = Accuracy, AUC = Area Under The Receiver Operating Characteristic Curve, MCC = Matthew's Correlation Coefficient.

**eTable 8. Classification accuracy based on structural MRI for HC vs recurrent MDD.**

| *eTable 8. Classification accuracy for multivariate biomarkers based on structural MRI for HC vs recurrent MDD.* | | | | | | | | |
|---|---|---|---|---|---|---|---|---|
| **Modality** | **Algorithm** | **BACC** | **ACC** | **Sensitivity** | **Specificity** | **AUC** | **MCC** | **n (HC/MDD)** |
| Freesurfer | Boosting Classifier | 51.1% (2.6%) | 70.9% (2.8%) | 13.8% (5.2%) | 88.4% (3.7%) | 55.1% (6.9%) | 0.03 (0.07) | 920/282 |
| | k-Nearest Neighbours | 51.5% (3.9%) | 73.0% (2.4%) | 11.0% (7.8%) | 92.0% (2.4%) | 55.0% (7.0%) | 0.04 (0.11) | 920/282 |
| | Logistic Regression | 60.2% (3.2%) | 60.9% (3.6%) | 58.8% (7.3%) | 61.5% (5.4%) | 63.3% (5.0%) | 0.17 (0.05) | 920/282 |
| | Gaussian Naive Bayes | 59.1% (5.8%) | 62.6% (4.0%) | 52.5% (11.2%) | 65.7% (4.3%) | 62.6% (5.5%) | 0.16 (0.1) | 920/282 |
| | Random Forest | 51.5% (2.0%) | 76.1% (1.1%) | 5.0% (4.2%) | 97.9% (0.8%) | 60.9% (5.9%) | 0.06 (0.09) | 920/282 |
| | Support Vector Machine | 53.8% (4.7%) | 66.1% (5.7%) | 30.5% (20.2%) | 77.1% (12.8%) | 53.8% (4.7%) | 0.07 (0.09) | 920/282 |
| VBM | Boosting Classifier | 55.8% (3.3%) | 73.5% (2.6%) | 22.1% (8.4%) | 89.4% (4.3%) | 61.0% (6.8%) | 0.14 (0.07) | 924/285 |
| | k-Nearest Neighbours | 50.7% (2.7%) | 73.9% (2.6%) | 6.7% (5.7%) | 94.7% (3.1%) | 52.8% (5.0%) | 0.02 (0.09) | 924/285 |
| | Logistic Regression | 54.8% (3.1%) | 49.7% (9.1%) | 64.3% (18.5%) | 45.3% (17.1%) | 52.7% (4.9%) | 0.09 (0.05) | 924/285 |
| | Gaussian Naive Bayes | 55.4% (4.1%) | 67.9% (6.5%) | 31.6% (20.4%) | 79.1% (14.3%) | 58.8% (6.4%) | 0.1 (0.09) | 924/285 |
| | Random Forest | 50.1% (1.3%) | 75.8% (1.0%) | 1.4% (3.0%) | 98.7% (1.1%) | 63.8% (5.8%) | -0.01 (0.08) | 924/285 |
| | Support Vector Machine | 51.6% (2.5%) | 64.6% (19.4%) | 27.0% (39.0%) | 76.2% (37.3%) | 51.6% (2.5%) | 0.06 (0.09) | 924/285 |
| DTI FA | Boosting Classifier | 49.9% (3.9%) | 70.5% (5.2%) | 12.9% (9.4%) | 86.9% (8.0%) | 49.8% (6.1%) | -0.0 (0.08) | 818/233 |
| | k-Nearest Neighbours | 51.4% (2.7%) | 74.0% (3.3%) | 10.7% (4.2%) | 92.0% (3.9%) | 51.9% (4.2%) | 0.05 (0.09) | 818/233 |
| | Logistic Regression | 50.4% (3.0%) | 69.8% (4.1%) | 15.5% (7.8%) | 85.3% (6.2%) | 51.8% (6.1%) | 0.01 (0.07) | 818/233 |
| | Gaussian Naive Bayes | 51.0% (6.1%) | 68.8% (4.8%) | 19.2% (14.3%) | 82.9% (7.3%) | 50.7% (7.1%) | 0.02 (0.14) | 818/233 |
| | Random Forest | 50.0% (0.0%) | 77.8% (0.4%) | 0.0% (0.0%) | 100.0% (0.0%) | 51.5% (5.7%) | 0.0 (0.0) | 818/233 |

| | | | | | | | | |
|---|---|---|---|---|---|---|---|---|
| | Support Vector Machine | 51.9% (3.2%) | 68.7% (6.5%) | 21.6% (13.7%) | 82.2% (11.7%) | 51.9% (3.2%) | 0.04 (0.08) | 818/233 |
| DTI MD | Boosting Classifier | 51.1% (3.8%) | 74.0% (4.4%) | 10.0% (8.0%) | 92.3% (6.2%) | 52.9% (6.1%) | 0.04 (0.13) | 818/233 |
| | k-Nearest Neighbours | 50.9% (4.5%) | 75.4% (2.7%) | 6.9% (9.8%) | 94.9% (3.9%) | 49.9% (9.3%) | 0.02 (0.14) | 818/233 |
| | Logistic Regression | 53.8% (5.3%) | 69.4% (5.6%) | 25.7% (7.0%) | 81.8% (6.8%) | 52.8% (8.6%) | 0.09 (0.12) | 818/233 |
| | Gaussian Naive Bayes | 49.5% (2.2%) | 73.9% (5.3%) | 5.6% (6.2%) | 93.4% (8.3%) | 50.7% (8.2%) | -0.01 (0.07) | 818/233 |
| | Random Forest | 50.0% (0.0%) | 77.8% (0.4%) | 0.0% (0.0%) | 100.0% (0.0%) | 56.7% (9.5%) | 0.0 (0.0) | 818/233 |
| | Support Vector Machine | 51.1% (7.1%) | 66.6% (9.0%) | 23.4% (17.8%) | 78.8% (14.0%) | 51.1% (7.1%) | 0.03 (0.16) | 818/233 |
| DTI Network Parameters | Boosting Classifier | 49.1% (3.0%) | 69.7% (3.4%) | 12.0% (6.3%) | 86.2% (4.7%) | 50.1% (6.1%) | -0.02 (0.08) | 818/233 |
| | k-Nearest Neighbours | 50.6% (2.0%) | 75.6% (1.5%) | 5.6% (5.4%) | 95.6% (2.7%) | 52.5% (4.7%) | 0.01 (0.07) | 818/233 |
| | Logistic Regression | 52.5% (4.2%) | 70.9% (3.8%) | 19.3% (6.0%) | 85.6% (3.9%) | 58.6% (6.5%) | 0.06 (0.1) | 818/233 |
| | Gaussian Naive Bayes | 50.1% (3.8%) | 63.9% (5.7%) | 25.3% (9.6%) | 74.9% (8.7%) | 50.1% (6.2%) | 0.0 (0.08) | 818/233 |
| | Random Forest | 50.2% (0.7%) | 77.9% (0.6%) | 0.4% (1.4%) | 100.0% (0.0%) | 54.8% (5.5%) | 0.02 (0.06) | 818/233 |
| | Support Vector Machine | 49.2% (5.7%) | 64.2% (5.2%) | 22.0% (14.8%) | 76.3% (8.4%) | 49.2% (5.7%) | -0.02 (0.11) | 818/233 |

Note: HC = Healthy Controls, MDD = Major Depressive Disorder, BACC = Balanced Accuracy, ACC = Accuracy, AUC = Area Under The Receiver Operating Characteristic Curve, MCC = Matthew's Correlation Coefficient.

**eTable 9. Classification accuracy based on functional MRI for HC vs recurrent MDD.**

| *eTable 9. Classification accuracy for multivariate biomarkers based on functional MRI for HC vs recurrent MDD.* | | | | | | | | |
|---|---|---|---|---|---|---|---|---|
| **Modality** | **Algorithm** | **BACC** | **ACC** | **Sensitivity** | **Specificity** | **AUC** | **MCC** | **n (HC/MDD)** |
| Face Matching Task | Boosting Classifier | 53.4% (5.3%) | 72.3% (3.0%) | 19.4% (12.2%) | 87.3% (4.0%) | 58.2% (5.3%) | 0.07 (0.12) | 654/185 |
| | k-Nearest Neighbours | 49.7% (2.9%) | 74.7% (2.4%) | 4.9% (5.4%) | 94.5% (2.8%) | 47.6% (7.6%) | -0.01 (0.1) | 654/185 |
| | Logistic Regression | 54.0% (6.6%) | 69.1% (3.5%) | 27.0% (13.1%) | 81.0% (3.5%) | 52.9% (9.6%) | 0.08 (0.14) | 654/185 |
| | Gaussian Naive Bayes | 57.4% (5.2%) | 68.3% (4.8%) | 37.9% (8.2%) | 76.9% (5.3%) | 58.2% (5.0%) | 0.14 (0.1) | 654/185 |
| | Random Forest | 49.9% (0.2%) | 77.8% (0.8%) | 0.0% (0.0%) | 99.8% (0.5%) | 55.4% (9.3%) | -0.01 (0.02) | 654/185 |
| | Support Vector Machine | 53.7% (4.7%) | 65.9% (5.1%) | 31.9% (12.5%) | 75.5% (8.3%) | 53.7% (4.7%) | 0.07 (0.09) | 654/185 |
| RS Connectivity | Boosting Classifier | 52.8% (3.7%) | 70.9% (2.1%) | 19.4% (9.1%) | 86.1% (3.4%) | 57.2% (8.3%) | 0.06 (0.09) | 700/207 |
| | k-Nearest Neighbours | 50.4% (3.7%) | 74.3% (3.7%) | 6.3% (5.2%) | 94.4% (4.3%) | 56.9% (9.6%) | 0.03 (0.14) | 700/207 |
| | Logistic Regression | 57.1% (6.0%) | 70.2% (4.6%) | 32.9% (10.6%) | 81.3% (5.1%) | 61.4% (6.7%) | 0.15 (0.12) | 700/207 |
| | Gaussian Naive Bayes | 61.7% (6.7%) | 66.8% (7.1%) | 52.2% (9.7%) | 71.1% (8.5%) | 62.4% (7.7%) | 0.21 (0.13) | 700/207 |
| | Random Forest | 50.6% (1.3%) | 77.3% (1.0%) | 1.5% (2.3%) | 99.7% (0.6%) | 62.3% (8.9%) | 0.05 (0.1) | 700/207 |
| | Support Vector Machine | 55.9% (6.6%) | 70.2% (4.0%) | 29.5% (14.0%) | 82.3% (4.8%) | 55.9% (6.6%) | 0.12 (0.13) | 700/207 |
| ALFF | Boosting Classifier | 55.8% (5.2%) | 72.6% (5.1%) | 25.1% (9.5%) | 86.6% (6.5%) | 60.2% (6.3%) | 0.14 (0.12) | 701/207 |
| | k-Nearest Neighbours | 55.1% (4.7%) | 77.0% (2.2%) | 14.9% (11.3%) | 95.3% (3.3%) | 63.2% (4.6%) | 0.16 (0.14) | 701/207 |
| | Logistic Regression | 61.3% (5.5%) | 67.6% (4.6%) | 49.7% (8.8%) | 72.9% (5.0%) | 66.8% (5.8%) | 0.2 (0.1) | 701/207 |
| | Gaussian Naive Bayes | 55.0% (6.0%) | 63.2% (4.8%) | 40.0% (11.9%) | 70.0% (6.0%) | 55.1% (6.8%) | 0.09 (0.11) | 701/207 |
| | Random Forest | 52.4% (2.3%) | 77.3% (1.5%) | 6.7% (4.6%) | 98.1% (1.8%) | 66.5% (7.2%) | 0.13 (0.11) | 701/207 |
| | Support Vector Machine | 56.1% (5.5%) | 72.4% (3.9%) | 26.1% (10.3%) | 86.0% (4.5%) | 56.1% (5.5%) | 0.14 (0.13) | 701/207 |
| fALFF | Boosting Classifier | 50.3% (7.1%) | 69.0% (5.4%) | 16.0% (12.6%) | 84.7% (5.2%) | 51.4% (12.2%) | 0.01 (0.16) | 701/207 |

| | | | | | | | | |
|---|---|---|---|---|---|---|---|---|
| | k-Nearest Neighbours | 50.9% (2.6%) | 76.8% (1.5%) | 3.3% (5.0%) | 98.4% (1.3%) | 53.8% (6.1%) | 0.04 (0.13) | 701/207 |
| | Logistic Regression | 53.8% (3.4%) | 66.7% (5.8%) | 30.0% (14.0%) | 77.6% (10.9%) | 56.2% (6.8%) | 0.08 (0.07) | 701/207 |
| | Gaussian Naive Bayes | 53.0% (10.2%) | 56.2% (8.2%) | 47.3% (16.2%) | 58.8% (7.9%) | 53.8% (10.5%) | 0.05 (0.18) | 701/207 |
| | Random Forest | 50.0% (0.0%) | 77.2% (0.5%) | 0.0% (0.0%) | 100.0% (0.0%) | 55.5% (8.2%) | 0.0 (0.0) | 701/207 |
| | Support Vector Machine | 52.9% (3.2%) | 69.4% (3.5%) | 22.7% (6.7%) | 83.2% (5.0%) | 52.9% (3.2%) | 0.07 (0.07) | 701/207 |
| LCOR | Boosting Classifier | 53.6% (4.8%) | 71.2% (3.3%) | 21.4% (9.0%) | 85.9% (3.0%) | 56.5% (8.4%) | 0.08 (0.11) | 701/207 |
| | k-Nearest Neighbours | 52.5% (2.8%) | 75.2% (1.9%) | 10.6% (6.4%) | 94.3% (2.4%) | 55.6% (8.1%) | 0.07 (0.09) | 701/207 |
| | Logistic Regression | 57.2% (3.7%) | 66.7% (4.2%) | 39.6% (13.2%) | 74.8% (8.5%) | 58.7% (5.0%) | 0.13 (0.06) | 701/207 |
| | Gaussian Naive Bayes | 58.9% (7.4%) | 62.0% (4.8%) | 53.2% (16.0%) | 64.6% (6.3%) | 58.9% (7.4%) | 0.15 (0.13) | 701/207 |
| | Random Forest | 50.2% (1.6%) | 76.7% (1.6%) | 1.5% (2.3%) | 98.9% (1.9%) | 58.6% (8.6%) | 0.02 (0.09) | 701/207 |
| | Support Vector Machine | 53.5% (5.5%) | 69.7% (3.9%) | 23.7% (11.3%) | 83.3% (4.7%) | 53.5% (5.5%) | 0.07 (0.13) | 701/207 |
| RS Network Parameters | Boosting Classifier | 52.2% (3.3%) | 72.2% (3.1%) | 15.4% (5.7%) | 89.0% (4.0%) | 55.1% (4.0%) | 0.06 (0.1) | 700/207 |
| | k-Nearest Neighbours | 53.1% (2.9%) | 73.5% (2.6%) | 15.5% (6.7%) | 90.7% (3.8%) | 57.3% (6.4%) | 0.08 (0.08) | 700/207 |
| | Logistic Regression | 52.2% (5.2%) | 69.0% (2.9%) | 21.2% (10.7%) | 83.1% (2.8%) | 57.5% (4.6%) | 0.04 (0.11) | 700/207 |
| | Gaussian Naive Bayes | 59.9% (4.5%) | 68.0% (4.7%) | 45.0% (6.9%) | 74.9% (5.8%) | 62.1% (5.3%) | 0.18 (0.09) | 700/207 |
| | Random Forest | 50.3% (0.8%) | 77.1% (0.4%) | 1.0% (2.0%) | 99.6% (0.7%) | 62.7% (6.0%) | 0.01 (0.05) | 700/207 |
| | Support Vector Machine | 54.8% (4.7%) | 69.2% (4.6%) | 28.1% (10.5%) | 81.4% (6.9%) | 54.8% (4.7%) | 0.1 (0.1) | 700/207 |

Note: HC = Healthy Controls, MDD = Major Depressive Disorder, BACC = Balanced Accuracy, ACC = Accuracy, AUC = Area Under The Receiver Operating Characteristic Curve, MCC = Matthew's Correlation Coefficient.

**eTable 10. Classification accuracy using neuroimaging modality integration for HC vs recurrent MDD.**

*eTable 10. Classification accuracy for multivariate biomarkers using neuroimaging modality integration for HC vs recurrent MDD.*

| Modality Integration | Modality | Algorithm | BACC | ACC | Sensitivity | Specificity | AUC | MCC | n (HC/MDD) |
|---|---|---|---|---|---|---|---|---|---|
| PCA-based | all | Boosting Classifier | 55.6% (5.6%) | 77.2% (3.2%) | 19.5% (12.6%) | 91.7% (4.1%) | 62.7% (10.0%) | 0.14 (0.14) | 567/143 |
| | | k-Nearest Neighbours | 50.3% (4.0%) | 77.2% (3.4%) | 5.5% (7.7%) | 95.2% (3.7%) | 48.4% (6.6%) | 0.01 (0.14) | 567/143 |
| | | Logistic Regression | 52.2% (4.9%) | 71.4% (17.7%) | 19.8% (27.8%) | 84.5% (28.5%) | 49.6% (8.0%) | 0.07 (0.15) | 567/143 |
| | | Gaussian Naive Bayes | 49.9% (3.3%) | 73.5% (3.4%) | 10.4% (8.2%) | 89.4% (5.2%) | 47.6% (7.5%) | -0.01 (0.09) | 567/143 |
| | | Random Forest | 50.4% (1.3%) | 79.3% (1.2%) | 2.0% (3.3%) | 98.8% (1.2%) | 49.8% (9.0%) | 0.01 (0.07) | 567/143 |
| | | Support Vector Machine | 51.2% (3.2%) | 40.4% (27.3%) | 69.3% (45.0%) | 33.2% (45.4%) | 51.2% (3.2%) | 0.04 (0.16) | 567/143 |
| Voting | all | all | 50.0% (0.0%) | 79.9% (0.7%) | 0.0% (0.0%) | 100.0% (0.0%) | 68.3% (6.7%) | 0.0 (0.0) | 567/143 |
| Voting | all | Boosting Classifier | 50.1% (1.0%) | 79.6% (1.0%) | 0.7% (2.3%) | 99.5% (0.9%) | 60.1% (7.2%) | 0.0 (0.05) | 567/143 |
| | | k-Nearest Neighbours | 50.0% (0.0%) | 79.9% (0.7%) | 0.0% (0.0%) | 100.0% (0.0%) | 59.6% (4.8%) | 0.0 (0.0) | 567/143 |
| | | Logistic Regression | 56.2% (5.9%) | 78.7% (3.3%) | 18.6% (11.7%) | 93.8% (3.9%) | 66.7% (7.9%) | 0.18 (0.17) | 567/143 |
| | | Gaussian Naive Bayes | 56.2% (5.4%) | 76.6% (3.5%) | 22.2% (10.4%) | 90.3% (3.5%) | 65.6% (8.5%) | 0.15 (0.13) | 567/143 |
| | | Random Forest | 50.0% (0.0%) | 79.9% (0.7%) | 0.0% (0.0%) | 100.0% (0.0%) | 53.5% (3.6%) | 0.0 (0.0) | 567/143 |
| | | Support Vector Machine | 51.5% (2.4%) | 79.0% (1.9%) | 5.6% (4.3%) | 97.5% (1.9%) | 61.6% (6.0%) | 0.07 (0.11) | 567/143 |
| Voting | Freesurfer | all | 51.1% (4.2%) | 75.9% (2.9%) | 9.7% (8.1%) | 92.6% (3.6%) | 57.3% (7.8%) | 0.03 (0.14) | 567/143 |
| | VBM | | 51.8% (2.7%) | 79.3% (2.1%) | 5.6% (4.5%) | 97.9% (2.0%) | 57.4% (6.4%) | 0.09 (0.13) | 567/143 |
| | DTI FA | | 50.5% (2.8%) | 79.0% (1.9%) | 2.8% (4.8%) | 98.2% (1.9%) | 52.9% (10.3%) | 0.03 (0.15) | 567/143 |
| | DTI MD | | 49.5% (0.7%) | 79.0% (1.6%) | 0.0% (0.0%) | 98.9% (1.5%) | 55.2% (8.3%) | -0.03 (0.04) | 567/143 |

| | | | | | | | | | |
|---|---|---|---|---|---|---|---|---|---|
| | DTI Network Parameters | | 49.3% (1.2%) | 78.3% (2.0%) | 0.7% (2.3%) | 97.9% (2.3%) | 50.4% (10.2%) | -0.04 (0.05) | 567/143 |
| | Face Matching Task | | 51.3% (2.6%) | 79.0% (2.9%) | 4.9% (3.4%) | 97.7% (2.9%) | 57.5% (6.6%) | 0.09 (0.14) | 567/143 |
| | RS Connectivity | | 53.2% (4.6%) | 79.2% (2.6%) | 9.8% (8.8%) | 96.6% (2.0%) | 63.7% (7.0%) | 0.11 (0.16) | 567/143 |
| | ALFF | | 54.7% (4.4%) | 80.0% (1.6%) | 12.4% (8.9%) | 97.0% (1.7%) | 65.8% (5.7%) | 0.16 (0.16) | 567/143 |
| | fALFF | | 50.7% (1.8%) | 78.5% (1.9%) | 4.1% (3.6%) | 97.2% (2.6%) | 54.8% (9.0%) | 0.04 (0.1) | 567/143 |
| | LCOR | | 52.2% (4.4%) | 78.0% (3.1%) | 8.9% (7.7%) | 95.4% (3.3%) | 59.4% (6.1%) | 0.08 (0.14) | 567/143 |
| | RS Network Parameters | | 50.7% (2.7%) | 78.5% (1.6%) | 4.2% (6.0%) | 97.2% (1.7%) | 62.9% (7.3%) | 0.01 (0.11) | 567/143 |

Note: HC = Healthy Controls, MDD = Major Depressive Disorder, BACC = Balanced Accuracy, ACC = Accuracy, AUC = Area Under The Receiver Operating Characteristic Curve, MCC = Matthew’s Correlation Coefficient.

### eTable 11. Classification accuracy based on structural MRI for HC vs MDD (male).

| *eTable 11. Classification accuracy for multivariate biomarkers based on structural MRI for HC vs MDD (male).* | | | | | | | | |
|---|---|---|---|---|---|---|---|---|
| **Modality** | **Algorithm** | **BACC** | **ACC** | **Sensitivity** | **Specificity** | **AUC** | **MCC** | **n (HC/MDD)** |
| Freesurfer | Boosting Classifier | 46.8% (4.4%) | 47.6% (4.8%) | 36.8% (5.8%) | 56.9% (11.1%) | 47.4% (3.5%) | -0.06 (0.09) | 327/280 |
| | k-Nearest Neighbours | 52.7% (5.0%) | 54.5% (5.0%) | 28.9% (6.8%) | 76.5% (7.5%) | 51.9% (7.1%) | 0.06 (0.11) | 327/280 |
| | Logistic Regression | 50.3% (6.9%) | 51.6% (7.3%) | 34.3% (8.3%) | 66.4% (14.0%) | 49.4% (5.8%) | 0.01 (0.14) | 327/280 |
| | Gaussian Naive Bayes | 50.1% (5.2%) | 50.9% (5.5%) | 38.9% (8.8%) | 61.2% (11.2%) | 50.3% (4.4%) | 0.0 (0.11) | 327/280 |
| | Random Forest | 48.3% (4.0%) | 49.9% (4.4%) | 27.5% (6.5%) | 69.1% (10.6%) | 50.6% (5.3%) | -0.03 (0.09) | 327/280 |
| | Support Vector Machine | 49.4% (3.8%) | 50.7% (3.9%) | 32.1% (8.2%) | 66.7% (8.7%) | 49.4% (3.8%) | -0.01 (0.08) | 327/280 |
| VBM | Boosting Classifier | 53.2% (5.2%) | 53.8% (5.3%) | 45.2% (8.6%) | 61.2% (9.6%) | 53.0% (6.1%) | 0.07 (0.11) | 330/281 |
| | k-Nearest Neighbours | 47.9% (4.1%) | 49.4% (4.3%) | 28.1% (5.0%) | 67.6% (8.1%) | 47.3% (6.9%) | -0.05 (0.09) | 330/281 |
| | Logistic Regression | 51.3% (5.5%) | 50.9% (5.8%) | 56.9% (15.9%) | 45.8% (17.2%) | 52.1% (5.5%) | 0.03 (0.12) | 330/281 |
| | Gaussian Naive Bayes | 50.1% (3.0%) | 52.7% (4.6%) | 17.5% (24.2%) | 82.7% (27.7%) | 51.5% (6.2%) | 0.02 (0.1) | 330/281 |
| | Random Forest | 54.1% (5.9%) | 56.1% (5.6%) | 29.2% (11.2%) | 79.1% (5.8%) | 56.5% (5.4%) | 0.09 (0.15) | 330/281 |
| | Support Vector Machine | 51.7% (3.2%) | 50.4% (4.4%) | 66.4% (42.2%) | 37.0% (41.7%) | 51.7% (3.2%) | 0.06 (0.12) | 330/281 |
| DTI FA | Boosting Classifier | 49.3% (6.5%) | 49.9% (6.9%) | 42.9% (7.5%) | 55.6% (11.8%) | 48.3% (7.8%) | -0.01 (0.13) | 291/238 |
| | k-Nearest Neighbours | 51.7% (9.8%) | 52.0% (9.6%) | 48.4% (13.3%) | 55.0% (9.5%) | 53.1% (7.4%) | 0.03 (0.2) | 291/238 |
| | Logistic Regression | 50.6% (6.6%) | 51.6% (6.5%) | 39.5% (10.9%) | 61.6% (9.2%) | 50.8% (6.8%) | 0.01 (0.13) | 291/238 |
| | Gaussian Naive Bayes | 55.4% (6.1%) | 55.4% (5.7%) | 55.1% (14.5%) | 55.7% (8.7%) | 55.7% (6.2%) | 0.11 (0.12) | 291/238 |
| | Random Forest | 55.5% (4.9%) | 57.7% (4.5%) | 33.6% (10.2%) | 77.4% (6.1%) | 57.9% (5.8%) | 0.12 (0.1) | 291/238 |
| | Support Vector Machine | 52.1% (5.4%) | 52.8% (5.8%) | 45.0% (5.1%) | 59.1% (10.7%) | 52.1% (5.4%) | 0.04 (0.11) | 291/238 |

| | | | | | | | | |
|---|---|---|---|---|---|---|---|---|
| DTI MD | Boosting Classifier | 49.5% (9.6%) | 50.3% (9.8%) | 41.2% (11.5%) | 57.7% (12.9%) | 51.3% (11.0%) | -0.01 (0.19) | 291/238 |
| | k-Nearest Neighbours | 53.2% (6.4%) | 54.3% (6.8%) | 42.1% (13.7%) | 64.3% (16.1%) | 55.8% (7.5%) | 0.07 (0.13) | 291/238 |
| | Logistic Regression | 49.7% (4.4%) | 51.2% (4.8%) | 33.3% (13.3%) | 66.0% (14.8%) | 49.3% (6.9%) | -0.0 (0.1) | 291/238 |
| | Gaussian Naive Bayes | 50.0% (6.5%) | 51.4% (7.3%) | 36.2% (13.1%) | 63.9% (19.1%) | 53.6% (8.3%) | 0.02 (0.16) | 291/238 |
| | Random Forest | 49.8% (4.2%) | 52.7% (4.4%) | 19.8% (6.7%) | 79.8% (8.7%) | 52.5% (5.9%) | -0.0 (0.1) | 291/238 |
| | Support Vector Machine | 51.0% (6.3%) | 52.2% (6.4%) | 39.9% (10.1%) | 62.2% (10.7%) | 51.0% (6.3%) | 0.02 (0.13) | 291/238 |
| DTI Network Parameters | Boosting Classifier | 47.0% (6.9%) | 47.6% (7.1%) | 39.9% (12.6%) | 54.0% (14.1%) | 45.9% (8.1%) | -0.06 (0.14) | 291/238 |
| | k-Nearest Neighbours | 47.9% (6.5%) | 49.9% (6.2%) | 28.1% (11.3%) | 67.7% (9.1%) | 50.3% (4.9%) | -0.05 (0.15) | 291/238 |
| | Logistic Regression | 48.0% (5.7%) | 49.1% (5.4%) | 36.6% (10.8%) | 59.5% (6.7%) | 47.7% (7.8%) | -0.04 (0.12) | 291/238 |
| | Gaussian Naive Bayes | 50.1% (5.6%) | 50.7% (5.2%) | 44.2% (12.6%) | 56.0% (8.3%) | 51.7% (7.1%) | 0.0 (0.11) | 291/238 |
| | Random Forest | 48.9% (4.7%) | 51.2% (5.4%) | 25.6% (12.0%) | 72.1% (15.1%) | 46.9% (8.1%) | -0.02 (0.12) | 291/238 |
| | Support Vector Machine | 49.8% (6.0%) | 50.3% (5.7%) | 44.6% (12.5%) | 55.0% (8.5%) | 49.8% (6.0%) | -0.01 (0.12) | 291/238 |

Note: HC = Healthy Controls, MDD = Major Depressive Disorder, BACC = Balanced Accuracy, ACC = Accuracy, AUC = Area Under The Receiver Operating Characteristic Curve, MCC = Matthew's Correlation Coefficient.

### eTable 12. Classification accuracy based on functional MRI for HC vs MDD (male).

| *eTable 12. Classification accuracy for multivariate biomarkers based on functional MRI for HC vs MDD (male).* | | | | | | | | |
|---|---|---|---|---|---|---|---|---|
| **Modality** | **Algorithm** | **BACC** | **ACC** | **Sensitivity** | **Specificity** | **AUC** | **MCC** | **n (HC/MDD)** |
| Face Matching Task | Boosting Classifier | 52.2% (8.8%) | 52.6% (8.8%) | 47.1% (12.0%) | 57.3% (11.2%) | 51.1% (9.5%) | 0.05 (0.18) | 227/195 |
| | k-Nearest Neighbours | 48.4% (9.7%) | 49.3% (9.8%) | 34.5% (10.9%) | 62.3% (16.3%) | 48.2% (8.8%) | -0.03 (0.21) | 227/195 |
| | Logistic Regression | 53.7% (7.2%) | 53.6% (7.3%) | 54.0% (9.9%) | 53.4% (10.8%) | 55.0% (10.7%) | 0.07 (0.15) | 227/195 |
| | Gaussian Naive Bayes | 51.6% (9.5%) | 51.5% (9.5%) | 52.0% (17.9%) | 51.2% (16.6%) | 52.6% (12.0%) | 0.04 (0.2) | 227/195 |
| | Random Forest | 50.5% (5.4%) | 52.2% (5.3%) | 27.2% (15.8%) | 73.8% (16.4%) | 52.8% (9.3%) | 0.01 (0.12) | 227/195 |
| | Support Vector Machine | 48.3% (7.8%) | 47.6% (7.5%) | 58.3% (25.0%) | 38.3% (22.2%) | 48.3% (7.8%) | -0.05 (0.18) | 227/195 |
| RS Connectivity | Boosting Classifier | 52.9% (8.2%) | 53.5% (8.4%) | 46.4% (12.9%) | 59.4% (13.3%) | 51.9% (10.7%) | 0.06 (0.17) | 249/209 |
| | k-Nearest Neighbours | 54.3% (7.8%) | 55.4% (7.7%) | 41.1% (13.2%) | 67.4% (8.3%) | 55.6% (9.3%) | 0.09 (0.16) | 249/209 |
| | Logistic Regression | 57.6% (8.6%) | 58.1% (8.4%) | 52.1% (13.1%) | 63.0% (8.4%) | 60.2% (9.0%) | 0.15 (0.17) | 249/209 |
| | Gaussian Naive Bayes | 57.8% (5.9%) | 58.3% (6.0%) | 53.5% (14.6%) | 62.2% (13.9%) | 59.5% (7.9%) | 0.16 (0.13) | 249/209 |
| | Random Forest | 58.9% (5.7%) | 60.3% (5.3%) | 44.4% (13.1%) | 73.4% (6.8%) | 60.6% (9.0%) | 0.19 (0.11) | 249/209 |
| | Support Vector Machine | 60.3% (8.1%) | 61.1% (7.6%) | 51.2% (17.9%) | 69.4% (9.7%) | 60.3% (8.1%) | 0.21 (0.17) | 249/209 |
| ALFF | Boosting Classifier | 55.6% (6.6%) | 55.9% (6.3%) | 51.7% (12.9%) | 59.5% (7.7%) | 58.1% (8.6%) | 0.11 (0.13) | 249/209 |
| | k-Nearest Neighbours | 57.3% (3.5%) | 58.3% (3.8%) | 46.4% (7.7%) | 68.2% (8.5%) | 59.8% (4.9%) | 0.15 (0.08) | 249/209 |
| | Logistic Regression | 60.4% (6.7%) | 61.2% (6.7%) | 52.2% (8.2%) | 68.7% (7.9%) | 62.5% (10.2%) | 0.21 (0.14) | 249/209 |
| | Gaussian Naive Bayes | 59.8% (5.7%) | 60.1% (5.9%) | 57.8% (11.2%) | 61.8% (11.9%) | 59.5% (5.6%) | 0.2 (0.12) | 249/209 |
| | Random Forest | 59.0% (4.9%) | 60.5% (4.7%) | 42.6% (10.1%) | 75.4% (7.0%) | 62.5% (9.5%) | 0.19 (0.1) | 249/209 |
| | Support Vector Machine | 59.1% (8.6%) | 60.1% (8.8%) | 47.4% (11.0%) | 70.7% (13.8%) | 59.1% (8.6%) | 0.19 (0.18) | 249/209 |
| fALFF | Boosting Classifier | 52.7% (7.6%) | 53.3% (7.6%) | 46.9% (13.4%) | 58.6% (12.9%) | 51.7% (9.1%) | 0.06 (0.16) | 249/209 |

| | | | | | | | | |
|---|---|---|---|---|---|---|---|---|
| | k-Nearest Neighbours | 50.0% (8.6%) | 51.1% (8.8%) | 37.3% (10.5%) | 62.7% (13.8%) | 52.2% (8.7%) | 0.0 (0.18) | 249/209 |
| | Logistic Regression | 54.2% (7.2%) | 55.0% (7.2%) | 45.0% (9.8%) | 63.5% (9.6%) | 56.3% (11.5%) | 0.09 (0.15) | 249/209 |
| | Gaussian Naive Bayes | 54.8% (7.0%) | 55.0% (6.9%) | 52.6% (9.8%) | 57.0% (7.0%) | 55.7% (7.6%) | 0.1 (0.14) | 249/209 |
| | Random Forest | 53.5% (8.8%) | 55.2% (8.8%) | 34.0% (12.3%) | 73.1% (11.8%) | 55.7% (9.4%) | 0.08 (0.19) | 249/209 |
| | Support Vector Machine | 53.6% (7.7%) | 55.0% (7.6%) | 36.9% (13.5%) | 70.3% (11.6%) | 53.6% (7.7%) | 0.08 (0.17) | 249/209 |
| LCOR | Boosting Classifier | 51.0% (5.5%) | 51.5% (6.0%) | 45.0% (13.1%) | 57.0% (16.1%) | 51.2% (3.2%) | 0.02 (0.11) | 249/209 |
| | k-Nearest Neighbours | 56.9% (7.4%) | 57.4% (7.0%) | 50.7% (13.7%) | 63.0% (6.7%) | 59.0% (6.6%) | 0.14 (0.15) | 249/209 |
| | Logistic Regression | 52.2% (7.1%) | 52.8% (6.7%) | 45.5% (17.1%) | 59.0% (12.1%) | 55.0% (4.1%) | 0.04 (0.15) | 249/209 |
| | Gaussian Naive Bayes | 53.5% (5.6%) | 53.5% (5.4%) | 53.5% (10.3%) | 53.4% (6.7%) | 53.0% (5.5%) | 0.07 (0.11) | 249/209 |
| | Random Forest | 51.8% (6.2%) | 53.3% (5.8%) | 34.9% (16.0%) | 68.6% (10.3%) | 54.9% (6.7%) | 0.04 (0.13) | 249/209 |
| | Support Vector Machine | 52.8% (7.4%) | 53.7% (6.8%) | 42.1% (18.5%) | 63.4% (10.8%) | 52.8% (7.4%) | 0.05 (0.15) | 249/209 |
| RS Network Parameters | Boosting Classifier | 52.4% (6.9%) | 53.3% (6.9%) | 43.5% (10.6%) | 61.4% (10.2%) | 53.6% (7.1%) | 0.05 (0.14) | 249/209 |
| | k-Nearest Neighbours | 51.8% (7.1%) | 53.3% (6.7%) | 35.3% (15.7%) | 68.2% (11.8%) | 53.2% (11.4%) | 0.03 (0.16) | 249/209 |
| | Logistic Regression | 52.5% (6.9%) | 54.1% (7.0%) | 33.9% (7.7%) | 71.0% (9.0%) | 56.0% (9.1%) | 0.05 (0.15) | 249/209 |
| | Gaussian Naive Bayes | 56.6% (6.9%) | 56.8% (6.4%) | 54.9% (13.6%) | 58.2% (6.8%) | 58.1% (7.5%) | 0.13 (0.14) | 249/209 |
| | Random Forest | 56.0% (6.1%) | 57.6% (6.1%) | 36.8% (11.5%) | 75.1% (9.3%) | 58.7% (8.7%) | 0.13 (0.13) | 249/209 |
| | Support Vector Machine | 53.5% (5.5%) | 54.6% (5.6%) | 42.0% (17.9%) | 65.1% (17.8%) | 53.5% (5.5%) | 0.08 (0.12) | 249/209 |

Note: HC = Healthy Controls, MDD = Major Depressive Disorder, BACC = Balanced Accuracy, ACC = Accuracy, AUC = Area Under The Receiver Operating Characteristic Curve, MCC = Matthew's Correlation Coefficient.

### eTable 13. Classification accuracy using neuroimaging modality integration for HC vs MDD (male).

*eTable 13. Classification accuracy for multivariate biomarkers using neuroimaging modality integration for HC vs MDD (male).*

| Modality Integration | Modality | Algorithm | BACC | ACC | Sensitivity | Specificity | AUC | MCC | n (HC/MDD) |
|---|---|---|---|---|---|---|---|---|---|
| PCA-based | all | Boosting Classifier | 49.7% (11.0%) | 50.6% (10.8%) | 40.6% (17.0%) | 58.7% (14.3%) | 48.9% (14.5%) | -0.01 (0.24) | 196/158 |
| | | k-Nearest Neighbours | 48.1% (9.3%) | 50.0% (9.7%) | 29.2% (12.7%) | 66.9% (17.3%) | 48.7% (10.6%) | -0.04 (0.2) | 196/158 |
| | | Logistic Regression | 49.1% (4.1%) | 48.3% (7.2%) | 56.7% (44.1%) | 41.5% (46.6%) | 53.0% (9.0%) | -0.05 (0.15) | 196/158 |
| | | Gaussian Naive Bayes | 51.1% (7.6%) | 50.8% (7.9%) | 51.0% (34.9%) | 51.2% (31.8%) | 51.2% (8.9%) | 0.01 (0.18) | 196/158 |
| | | Random Forest | 53.5% (6.9%) | 56.8% (7.3%) | 23.5% (15.4%) | 83.6% (16.0%) | 55.6% (9.6%) | 0.09 (0.18) | 196/158 |
| | | Support Vector Machine | 48.9% (4.3%) | 46.6% (5.0%) | 67.3% (38.5%) | 30.4% (36.3%) | 48.9% (4.3%) | -0.03 (0.14) | 196/158 |
| Voting | all | all | 58.2% (6.3%) | 61.6% (5.8%) | 27.7% (10.5%) | 88.7% (6.0%) | 64.6% (8.8%) | 0.21 (0.16) | 196/158 |
| Voting | all | Boosting Classifier | 54.8% (6.4%) | 56.5% (6.2%) | 39.2% (10.5%) | 70.4% (9.5%) | 54.6% (10.1%) | 0.1 (0.14) | 196/158 |
| | | k-Nearest Neighbours | 55.5% (5.3%) | 59.1% (5.0%) | 22.7% (10.0%) | 88.3% (5.8%) | 59.8% (6.3%) | 0.14 (0.14) | 196/158 |
| | | Logistic Regression | 55.4% (6.1%) | 57.6% (6.3%) | 35.3% (9.5%) | 75.4% (11.7%) | 62.7% (9.8%) | 0.13 (0.14) | 196/158 |
| | | Gaussian Naive Bayes | 59.4% (7.9%) | 61.3% (7.3%) | 42.2% (13.6%) | 76.5% (8.1%) | 63.5% (6.3%) | 0.2 (0.17) | 196/158 |
| | | Random Forest | 53.1% (3.3%) | 57.3% (3.4%) | 13.3% (6.2%) | 92.8% (4.9%) | 64.5% (12.8%) | 0.11 (0.11) | 196/158 |
| | | Support Vector Machine | 57.2% (9.2%) | 59.0% (9.3%) | 39.9% (10.7%) | 74.4% (10.4%) | 61.1% (7.9%) | 0.15 (0.2) | 196/158 |
| Voting | Freesurfer | all | 52.4% (6.8%) | 55.1% (7.3%) | 27.8% (9.4%) | 76.9% (12.8%) | 52.6% (9.9%) | 0.06 (0.16) | 196/158 |
| | VBM | | 51.5% (5.1%) | 55.3% (4.9%) | 16.3% (10.7%) | 86.7% (10.3%) | 56.2% (7.2%) | 0.05 (0.18) | 196/158 |
| | DTI FA | | 54.0% (5.6%) | 56.5% (5.7%) | 31.0% (5.1%) | 77.1% (7.8%) | 56.2% (7.0%) | 0.09 (0.13) | 196/158 |
| | DTI MD | | 52.1% (7.1%) | 55.7% (7.3%) | 20.2% (8.1%) | 84.1% (12.6%) | 54.1% (11.4%) | 0.07 (0.19) | 196/158 |

| | | | | | | | | | |
|---|---|---|---|---|---|---|---|---|---|
| | DTI Network Parameters | | 49.5% (6.0%) | 52.0% (6.2%) | 27.1% (8.1%) | 71.9% (8.3%) | 47.4% (9.5%) | -0.01 (0.13) | 196/158 |
| | Face Matching Task | | 49.9% (6.8%) | 52.3% (6.8%) | 27.8% (11.0%) | 71.9% (12.2%) | 52.3% (9.2%) | 0.0 (0.17) | 196/158 |
| | RS Connectivity | | 61.3% (7.3%) | 63.0% (7.6%) | 45.0% (9.0%) | 77.5% (11.3%) | 63.3% (10.8%) | 0.24 (0.16) | 196/158 |
| | ALFF | | 63.0% (8.4%) | 64.9% (8.1%) | 45.5% (12.7%) | 80.5% (8.3%) | 65.2% (9.5%) | 0.28 (0.18) | 196/158 |
| | fALFF | | 53.2% (7.0%) | 55.6% (7.5%) | 29.8% (9.1%) | 76.5% (11.1%) | 54.1% (7.0%) | 0.08 (0.17) | 196/158 |
| | LCOR | | 52.6% (7.7%) | 54.2% (7.7%) | 36.8% (9.4%) | 68.4% (8.4%) | 52.3% (7.8%) | 0.05 (0.16) | 196/158 |
| | RS Network Parameters | | 54.4% (6.4%) | 56.8% (6.3%) | 32.8% (9.8%) | 76.1% (10.2%) | 56.6% (4.7%) | 0.1 (0.14) | 196/158 |

Note: HC = Healthy Controls, MDD = Major Depressive Disorder, BACC = Balanced Accuracy, ACC = Accuracy, AUC = Area Under The Receiver Operating Characteristic Curve, MCC = Matthew's Correlation Coefficient.

**eTable 14. Classification accuracy based on structural MRI for HC vs MDD (female).**

| *eTable 14. Classification accuracy for multivariate biomarkers based on structural MRI for HC vs MDD (female).* | | | | | | | | |
|---|---|---|---|---|---|---|---|---|
| **Modality** | **Algorithm** | **BACC** | **ACC** | **Sensitivity** | **Specificity** | **AUC** | **MCC** | **n (HC/MDD)** |
| Freesurfer | Boosting Classifier | 54.3% (6.1%) | 54.6% (5.9%) | 48.0% (11.4%) | 60.6% (7.3%) | 55.6% (6.4%) | 0.09 (0.12) | 593/533 |
| | k-Nearest Neighbours | 51.4% (4.8%) | 51.7% (4.8%) | 46.9% (9.1%) | 56.0% (8.2%) | 51.6% (5.2%) | 0.03 (0.1) | 593/533 |
| | Logistic Regression | 53.5% (3.7%) | 54.0% (3.8%) | 45.0% (8.7%) | 62.1% (9.5%) | 56.0% (3.0%) | 0.07 (0.08) | 593/533 |
| | Gaussian Naive Bayes | 53.9% (3.5%) | 53.8% (3.6%) | 54.4% (5.9%) | 53.3% (6.8%) | 56.4% (3.5%) | 0.08 (0.07) | 593/533 |
| | Random Forest | 53.1% (5.2%) | 53.8% (5.1%) | 38.8% (8.4%) | 67.3% (6.7%) | 55.6% (3.9%) | 0.06 (0.11) | 593/533 |
| | Support Vector Machine | 54.9% (2.9%) | 55.6% (2.8%) | 42.0% (13.8%) | 67.8% (12.6%) | 54.9% (2.9%) | 0.11 (0.06) | 593/533 |
| VBM | Boosting Classifier | 53.1% (2.5%) | 53.1% (2.4%) | 52.2% (6.2%) | 53.9% (4.1%) | 53.1% (3.3%) | 0.06 (0.05) | 594/536 |
| | k-Nearest Neighbours | 49.6% (4.2%) | 50.4% (4.3%) | 33.8% (7.9%) | 65.5% (10.7%) | 51.1% (5.9%) | -0.01 (0.09) | 594/536 |
| | Logistic Regression | 51.8% (4.4%) | 50.9% (4.7%) | 67.8% (16.6%) | 35.8% (18.0%) | 52.0% (4.8%) | 0.04 (0.09) | 594/536 |
| | Gaussian Naive Bayes | 51.0% (3.8%) | 51.9% (3.4%) | 31.5% (24.6%) | 70.4% (21.5%) | 50.7% (6.2%) | 0.02 (0.09) | 594/536 |
| | Random Forest | 54.5% (3.4%) | 55.0% (3.4%) | 42.9% (5.5%) | 66.0% (6.0%) | 57.2% (4.6%) | 0.09 (0.07) | 594/536 |
| | Support Vector Machine | 53.5% (4.1%) | 53.3% (4.3%) | 58.0% (15.1%) | 49.0% (17.2%) | 53.5% (4.1%) | 0.07 (0.08) | 594/536 |
| DTI FA | Boosting Classifier | 54.4% (6.9%) | 54.7% (6.7%) | 51.2% (11.7%) | 57.7% (8.1%) | 56.3% (7.2%) | 0.09 (0.14) | 527/447 |
| | k-Nearest Neighbours | 50.8% (3.2%) | 51.4% (3.1%) | 43.2% (8.3%) | 58.5% (7.6%) | 50.7% (3.9%) | 0.02 (0.06) | 527/447 |
| | Logistic Regression | 52.8% (5.3%) | 53.3% (5.1%) | 46.6% (9.0%) | 59.0% (5.8%) | 54.5% (5.3%) | 0.06 (0.11) | 527/447 |
| | Gaussian Naive Bayes | 49.5% (3.6%) | 49.3% (4.5%) | 51.9% (21.3%) | 47.1% (23.7%) | 49.8% (2.6%) | -0.01 (0.08) | 527/447 |
| | Random Forest | 55.2% (5.1%) | 56.9% (5.2%) | 34.0% (6.8%) | 76.3% (8.0%) | 58.0% (5.3%) | 0.12 (0.11) | 527/447 |

| | | | | | | | | |
|---|---|---|---|---|---|---|---|---|
| | Support Vector Machine | 54.6% (5.3%) | 55.3% (5.2%) | 45.4% (13.0%) | 63.7% (11.0%) | 54.6% (5.3%) | 0.09 (0.11) | 527/447 |
| DTI MD | Boosting Classifier | 55.8% (4.9%) | 56.4% (4.8%) | 48.4% (9.9%) | 63.2% (7.9%) | 55.6% (5.3%) | 0.12 (0.1) | 527/447 |
| | k-Nearest Neighbours | 52.4% (4.4%) | 53.5% (4.2%) | 39.5% (14.9%) | 65.2% (13.4%) | 51.7% (2.9%) | 0.05 (0.1) | 527/447 |
| | Logistic Regression | 52.8% (3.3%) | 53.4% (3.3%) | 45.2% (7.0%) | 60.3% (5.4%) | 53.5% (3.8%) | 0.06 (0.07) | 527/447 |
| | Gaussian Naive Bayes | 53.5% (4.8%) | 53.0% (4.8%) | 60.2% (8.6%) | 46.9% (7.2%) | 56.6% (4.2%) | 0.07 (0.1) | 527/447 |
| | Random Forest | 55.0% (3.0%) | 56.9% (3.0%) | 31.6% (7.3%) | 78.4% (7.6%) | 56.6% (3.8%) | 0.11 (0.07) | 527/447 |
| | Support Vector Machine | 53.0% (5.2%) | 53.6% (4.9%) | 46.3% (10.2%) | 59.8% (3.0%) | 53.0% (5.2%) | 0.06 (0.11) | 527/447 |
| DTI Network Parameters | Boosting Classifier | 49.2% (5.0%) | 49.6% (5.2%) | 45.0% (8.3%) | 53.5% (11.0%) | 49.8% (4.4%) | -0.01 (0.1) | 527/447 |
| | k-Nearest Neighbours | 50.7% (4.0%) | 52.0% (4.1%) | 35.8% (9.4%) | 65.7% (9.3%) | 51.2% (6.3%) | 0.02 (0.09) | 527/447 |
| | Logistic Regression | 53.7% (4.8%) | 54.4% (4.9%) | 44.9% (7.3%) | 62.4% (8.8%) | 54.2% (5.1%) | 0.08 (0.1) | 527/447 |
| | Gaussian Naive Bayes | 52.4% (3.2%) | 52.5% (3.5%) | 51.3% (9.2%) | 53.5% (10.7%) | 54.4% (3.2%) | 0.05 (0.07) | 527/447 |
| | Random Forest | 50.1% (3.8%) | 52.4% (3.7%) | 23.0% (11.6%) | 77.2% (9.9%) | 53.6% (7.3%) | -0.0 (0.09) | 527/447 |
| | Support Vector Machine | 52.2% (4.1%) | 52.9% (4.1%) | 43.8% (7.2%) | 60.6% (6.7%) | 52.2% (4.1%) | 0.04 (0.08) | 527/447 |

Note: HC = Healthy Controls, MDD = Major Depressive Disorder, BACC = Balanced Accuracy, ACC = Accuracy, AUC = Area Under The Receiver Operating Characteristic Curve, MCC = Matthew's Correlation Coefficient.

**eTable 15. Classification accuracy based on functional MRI for HC vs MDD (female).**

| *eTable 15. Classification accuracy for multivariate biomarkers based on functional MRI for HC vs MDD (female).* | | | | | | | | |
|---|---|---|---|---|---|---|---|---|
| **Modality** | **Algorithm** | **BACC** | **ACC** | **Sensitivity** | **Specificity** | **AUC** | **MCC** | **n (HC/MDD)** |
| Face Matching Task | Boosting Classifier | 52.8% (5.6%) | 53.0% (5.6%) | 49.1% (5.2%) | 56.4% (7.6%) | 53.0% (4.7%) | 0.06 (0.11) | 427/385 |
| | k-Nearest Neighbours | 51.2% (5.9%) | 52.1% (5.9%) | 33.8% (13.7%) | 68.7% (12.4%) | 52.5% (5.7%) | 0.02 (0.13) | 427/385 |
| | Logistic Regression | 50.5% (6.2%) | 50.7% (5.9%) | 45.2% (16.2%) | 55.8% (9.6%) | 51.7% (7.0%) | 0.01 (0.13) | 427/385 |
| | Gaussian Naive Bayes | 56.0% (3.9%) | 55.2% (4.3%) | 71.0% (11.1%) | 41.0% (13.3%) | 57.5% (4.2%) | 0.13 (0.08) | 427/385 |
| | Random Forest | 55.6% (5.9%) | 56.5% (5.8%) | 37.4% (7.3%) | 73.8% (7.7%) | 57.3% (6.6%) | 0.12 (0.13) | 427/385 |
| | Support Vector Machine | 51.4% (6.4%) | 51.6% (6.6%) | 47.8% (11.8%) | 55.0% (16.2%) | 51.4% (6.4%) | 0.03 (0.13) | 427/385 |
| RS Connectivity | Boosting Classifier | 53.2% (4.1%) | 53.3% (4.1%) | 50.9% (5.0%) | 55.4% (8.1%) | 55.2% (4.2%) | 0.06 (0.08) | 451/422 |
| | k-Nearest Neighbours | 55.6% (5.1%) | 56.0% (5.2%) | 42.9% (5.7%) | 68.3% (8.8%) | 58.6% (7.0%) | 0.12 (0.11) | 451/422 |
| | Logistic Regression | 58.4% (4.2%) | 58.5% (4.3%) | 54.3% (5.7%) | 62.5% (8.7%) | 62.3% (4.6%) | 0.17 (0.09) | 451/422 |
| | Gaussian Naive Bayes | 57.4% (5.1%) | 57.6% (5.0%) | 51.9% (7.6%) | 63.0% (5.5%) | 59.7% (6.2%) | 0.15 (0.1) | 451/422 |
| | Random Forest | 56.9% (4.5%) | 57.2% (4.5%) | 47.9% (5.9%) | 65.8% (6.4%) | 61.0% (4.4%) | 0.14 (0.09) | 451/422 |
| | Support Vector Machine | 58.1% (3.6%) | 58.3% (3.6%) | 51.5% (6.8%) | 64.7% (7.8%) | 58.1% (3.6%) | 0.16 (0.07) | 451/422 |
| ALFF | Boosting Classifier | 57.4% (5.6%) | 57.5% (5.7%) | 55.3% (7.7%) | 59.6% (8.7%) | 59.3% (7.0%) | 0.15 (0.11) | 452/422 |
| | k-Nearest Neighbours | 59.2% (4.8%) | 59.5% (4.8%) | 51.4% (6.3%) | 67.0% (6.7%) | 60.9% (6.9%) | 0.19 (0.1) | 452/422 |
| | Logistic Regression | 59.1% (5.9%) | 59.3% (5.7%) | 54.3% (11.9%) | 64.0% (5.7%) | 62.4% (6.0%) | 0.19 (0.12) | 452/422 |
| | Gaussian Naive Bayes | 60.3% (4.5%) | 60.4% (4.5%) | 54.8% (7.6%) | 65.8% (7.7%) | 60.1% (4.3%) | 0.21 (0.09) | 452/422 |
| | Random Forest | 60.5% (6.8%) | 60.7% (6.7%) | 55.7% (10.1%) | 65.3% (6.0%) | 63.3% (5.4%) | 0.21 (0.14) | 452/422 |
| | Support Vector Machine | 61.6% (4.5%) | 61.8% (4.5%) | 56.9% (7.7%) | 66.4% (5.1%) | 61.6% (4.5%) | 0.23 (0.09) | 452/422 |
| fALFF | Boosting Classifier | 54.4% (6.0%) | 54.5% (6.1%) | 53.3% (9.1%) | 55.6% (9.9%) | 55.6% (4.8%) | 0.09 (0.12) | 452/422 |

| | | | | | | | | |
|---|---|---|---|---|---|---|---|---|
| | k-Nearest Neighbours | 56.0% (5.1%) | 56.6% (5.0%) | 35.8% (11.2%) | 76.1% (6.5%) | 58.7% (6.0%) | 0.13 (0.11) | 452/422 |
| | Logistic Regression | 57.9% (4.1%) | 58.0% (4.1%) | 53.3% (6.1%) | 62.4% (7.5%) | 59.8% (3.9%) | 0.16 (0.08) | 452/422 |
| | Gaussian Naive Bayes | 55.3% (2.7%) | 55.4% (2.6%) | 53.1% (6.0%) | 57.5% (4.4%) | 56.1% (3.3%) | 0.11 (0.05) | 452/422 |
| | Random Forest | 55.4% (4.7%) | 55.8% (4.7%) | 43.1% (8.3%) | 67.7% (9.2%) | 58.9% (6.1%) | 0.11 (0.1) | 452/422 |
| | Support Vector Machine | 58.6% (3.9%) | 58.8% (3.9%) | 53.1% (5.0%) | 64.2% (6.9%) | 58.6% (3.9%) | 0.17 (0.08) | 452/422 |
| LCOR | Boosting Classifier | 50.8% (4.9%) | 51.0% (4.7%) | 45.5% (11.8%) | 56.2% (6.7%) | 51.8% (8.1%) | 0.01 (0.1) | 452/422 |
| | k-Nearest Neighbours | 56.2% (6.5%) | 56.5% (6.6%) | 46.7% (9.2%) | 65.7% (10.7%) | 59.7% (4.6%) | 0.13 (0.14) | 452/422 |
| | Logistic Regression | 57.7% (2.7%) | 57.8% (2.7%) | 54.5% (5.5%) | 60.8% (4.1%) | 59.3% (4.8%) | 0.15 (0.05) | 452/422 |
| | Gaussian Naive Bayes | 55.9% (4.4%) | 56.1% (4.3%) | 52.8% (9.7%) | 59.1% (5.0%) | 55.8% (4.3%) | 0.12 (0.09) | 452/422 |
| | Random Forest | 54.5% (5.7%) | 54.8% (5.5%) | 47.4% (11.6%) | 61.7% (3.1%) | 59.4% (6.4%) | 0.09 (0.11) | 452/422 |
| | Support Vector Machine | 56.5% (3.8%) | 56.6% (3.7%) | 51.4% (8.2%) | 61.5% (7.3%) | 56.5% (3.8%) | 0.13 (0.08) | 452/422 |
| RS Network Parameters | Boosting Classifier | 50.5% (4.9%) | 50.7% (4.8%) | 41.3% (10.9%) | 59.6% (5.1%) | 52.6% (5.3%) | 0.01 (0.1) | 451/422 |
| | k-Nearest Neighbours | 52.9% (3.8%) | 53.4% (3.9%) | 38.6% (7.6%) | 67.2% (8.9%) | 55.6% (3.8%) | 0.06 (0.08) | 451/422 |
| | Logistic Regression | 57.8% (5.0%) | 58.0% (5.0%) | 52.6% (7.2%) | 63.0% (6.2%) | 61.6% (6.0%) | 0.16 (0.1) | 451/422 |
| | Gaussian Naive Bayes | 53.1% (6.8%) | 53.2% (6.8%) | 50.9% (7.4%) | 55.2% (7.9%) | 54.5% (7.9%) | 0.06 (0.14) | 451/422 |
| | Random Forest | 56.2% (5.0%) | 56.5% (5.1%) | 47.2% (7.2%) | 65.2% (8.5%) | 57.8% (6.4%) | 0.13 (0.1) | 451/422 |
| | Support Vector Machine | 60.3% (6.9%) | 60.5% (7.0%) | 55.4% (7.1%) | 65.2% (9.2%) | 60.3% (6.9%) | 0.21 (0.14) | 451/422 |

Note: HC = Healthy Controls, MDD = Major Depressive Disorder, BACC = Balanced Accuracy, ACC = Accuracy, AUC = Area Under The Receiver Operating Characteristic Curve, MCC = Matthew's Correlation Coefficient.

### eTable 16. Classification accuracy using neuroimaging modality integration for HC vs MDD (female).

*eTable 16. Classification accuracy for multivariate biomarkers using neuroimaging modality integration for HC vs MDD (female).*

| Modality Integration | Modality | Algorithm | BACC | ACC | Sensitivity | Specificity | AUC | MCC | n (HC/MDD) |
|---|---|---|---|---|---|---|---|---|---|
| PCA-based | all | Boosting Classifier | 57.9% (4.4%) | 58.6% (4.3%) | 49.6% (10.6%) | 66.3% (7.7%) | 56.3% (5.0%) | 0.16 (0.09) | 371/313 |
| | | k-Nearest Neighbours | 51.7% (4.6%) | 52.9% (4.7%) | 37.4% (9.9%) | 66.0% (9.8%) | 53.6% (7.5%) | 0.04 (0.1) | 371/313 |
| | | Logistic Regression | 50.7% (2.3%) | 53.7% (3.1%) | 15.7% (20.8%) | 85.7% (21.0%) | 50.8% (5.4%) | 0.03 (0.08) | 371/313 |
| | | Gaussian Naive Bayes | 50.5% (2.9%) | 53.4% (3.2%) | 18.1% (25.0%) | 83.0% (24.0%) | 53.4% (5.2%) | 0.03 (0.11) | 371/313 |
| | | Random Forest | 54.1% (5.9%) | 56.6% (5.8%) | 25.2% (10.0%) | 83.0% (8.5%) | 58.2% (9.3%) | 0.11 (0.16) | 371/313 |
| | | Support Vector Machine | 49.0% (2.4%) | 49.1% (5.6%) | 46.8% (48.4%) | 51.3% (50.3%) | 49.0% (2.4%) | -0.03 (0.1) | 371/313 |
| Voting | all | all | 61.5% (5.9%) | 63.0% (5.3%) | 43.8% (13.7%) | 79.2% (4.0%) | 65.9% (5.6%) | 0.25 (0.12) | 371/313 |
| Voting | all | Boosting Classifier | 56.6% (6.1%) | 57.6% (6.0%) | 44.1% (10.7%) | 69.0% (6.5%) | 59.8% (5.3%) | 0.14 (0.13) | 371/313 |
| | | k-Nearest Neighbours | 57.4% (4.8%) | 59.5% (4.5%) | 32.0% (10.1%) | 82.7% (6.7%) | 60.1% (6.9%) | 0.17 (0.11) | 371/313 |
| | | Logistic Regression | 60.2% (4.4%) | 61.0% (4.1%) | 50.5% (10.4%) | 69.8% (6.3%) | 63.1% (5.8%) | 0.21 (0.09) | 371/313 |
| | | Gaussian Naive Bayes | 60.8% (5.9%) | 61.3% (5.7%) | 54.7% (10.9%) | 66.8% (6.1%) | 62.6% (7.7%) | 0.22 (0.12) | 371/313 |
| | | Random Forest | 59.0% (5.4%) | 61.1% (4.9%) | 34.5% (11.6%) | 83.6% (4.4%) | 64.1% (5.5%) | 0.21 (0.11) | 371/313 |
| | | Support Vector Machine | 61.3% (6.1%) | 62.4% (5.9%) | 48.2% (10.1%) | 74.4% (5.9%) | 65.3% (6.0%) | 0.23 (0.12) | 371/313 |
| Voting | Freesurfer | all | 49.7% (7.7%) | 51.0% (7.7%) | 34.2% (8.5%) | 65.2% (8.9%) | 52.6% (9.8%) | -0.0 (0.16) | 371/313 |
| | VBM | | 52.6% (5.0%) | 53.8% (4.6%) | 38.7% (10.0%) | 66.6% (5.1%) | 53.8% (6.4%) | 0.05 (0.1) | 371/313 |
| | DTI FA | | 53.8% (4.6%) | 55.7% (4.5%) | 31.3% (8.3%) | 76.3% (7.0%) | 55.6% (6.2%) | 0.09 (0.1) | 371/313 |
| | DTI MD | | 56.1% (5.8%) | 58.0% (5.7%) | 33.2% (8.5%) | 79.0% (7.6%) | 56.4% (3.9%) | 0.14 (0.13) | 371/313 |

| | | | | | | | | | |
|---|---|---|---|---|---|---|---|---|---|
| | DTI Network Parameters | | 51.5% (7.6%) | 53.8% (7.8%) | 24.3% (8.0%) | 78.7% (9.8%) | 52.9% (5.0%) | 0.04 (0.17) | 371/313 |
| | Face Matching Task | | 54.6% (3.8%) | 56.0% (3.5%) | 37.7% (10.7%) | 71.4% (7.6%) | 56.8% (6.0%) | 0.1 (0.08) | 371/313 |
| | RS Connectivity | | 58.0% (4.5%) | 59.2% (4.2%) | 43.1% (8.8%) | 72.8% (4.8%) | 61.7% (5.7%) | 0.17 (0.09) | 371/313 |
| | ALFF | | 62.7% (5.5%) | 63.6% (5.4%) | 52.7% (9.3%) | 72.8% (6.7%) | 64.8% (6.0%) | 0.26 (0.11) | 371/313 |
| | fALFF | | 59.5% (5.4%) | 60.7% (5.2%) | 45.4% (8.8%) | 73.6% (5.7%) | 60.6% (6.0%) | 0.2 (0.11) | 371/313 |
| | LCOR | | 56.8% (4.7%) | 58.0% (4.7%) | 42.5% (7.2%) | 71.2% (7.1%) | 59.5% (6.3%) | 0.14 (0.1) | 371/313 |
| | RS Network Parameters | | 58.4% (7.1%) | 59.8% (6.9%) | 42.5% (9.9%) | 74.4% (5.4%) | 59.8% (7.9%) | 0.18 (0.15) | 371/313 |

Note: HC = Healthy Controls, MDD = Major Depressive Disorder, BACC = Balanced Accuracy, ACC = Accuracy, AUC = Area Under The Receiver Operating Characteristic Curve, MCC = Matthew's Correlation Coefficient.

**eTable 17. Classification accuracy based on structural MRI for HC vs MDD (age 24 - 28).**

| *eTable 17. Classification accuracy for multivariate biomarkers based on structural MRI for HC vs MDD (age 24 -28).* | | | | | | | | |
|---|---|---|---|---|---|---|---|---|
| **Modality** | **Algorithm** | **BACC** | **ACC** | **Sensitivity** | **Specificity** | **AUC** | **MCC** | **n (HC/MDD)** |
| Freesurfer | Boosting Classifier | 53.5% (8.5%) | 55.1% (8.7%) | 41.8% (14.0%) | 65.3% (14.0%) | 51.5% (12.9%) | 0.07 (0.17) | 224/166 |
| | k-Nearest Neighbours | 50.0% (5.9%) | 52.8% (6.5%) | 30.8% (8.2%) | 69.1% (9.8%) | 51.6% (5.6%) | 0.0 (0.13) | 224/166 |
| | Logistic Regression | 52.4% (6.9%) | 55.9% (6.9%) | 28.9% (11.7%) | 75.9% (11.6%) | 57.6% (6.0%) | 0.06 (0.16) | 224/166 |
| | Gaussian Naive Bayes | 52.8% (7.8%) | 54.6% (8.0%) | 39.4% (15.2%) | 66.1% (13.8%) | 56.2% (9.6%) | 0.06 (0.16) | 224/166 |
| | Random Forest | 56.3% (6.2%) | 59.2% (5.7%) | 36.3% (14.6%) | 76.3% (8.4%) | 56.8% (11.1%) | 0.13 (0.14) | 224/166 |
| | Support Vector Machine | 47.9% (8.3%) | 50.3% (9.2%) | 33.1% (17.3%) | 62.8% (20.7%) | 47.9% (8.3%) | -0.03 (0.2) | 224/166 |
| VBM | Boosting Classifier | 49.0% (8.3%) | 49.9% (7.9%) | 42.9% (12.7%) | 55.0% (6.9%) | 49.0% (7.3%) | -0.02 (0.17) | 225/166 |
| | k-Nearest Neighbours | 50.9% (7.5%) | 52.2% (7.7%) | 42.8% (9.8%) | 59.0% (10.0%) | 49.9% (10.7%) | 0.02 (0.15) | 225/166 |
| | Logistic Regression | 49.6% (6.6%) | 46.8% (7.0%) | 67.0% (32.7%) | 32.2% (29.6%) | 45.4% (6.0%) | 0.0 (0.16) | 225/166 |
| | Gaussian Naive Bayes | 48.5% (5.6%) | 51.1% (10.5%) | 29.8% (32.7%) | 67.3% (40.6%) | 50.5% (10.3%) | -0.04 (0.17) | 225/166 |
| | Random Forest | 49.6% (6.7%) | 55.0% (6.4%) | 13.9% (10.8%) | 85.4% (7.7%) | 51.5% (13.5%) | -0.01 (0.19) | 225/166 |
| | Support Vector Machine | 48.3% (5.4%) | 46.0% (9.0%) | 63.5% (42.2%) | 33.1% (43.4%) | 48.3% (5.4%) | -0.05 (0.19) | 225/166 |
| DTI FA | Boosting Classifier | 46.7% (9.2%) | 47.7% (9.5%) | 40.4% (11.4%) | 52.9% (14.2%) | 47.2% (11.1%) | -0.07 (0.18) | 204/144 |
| | k-Nearest Neighbours | 47.0% (8.4%) | 50.2% (8.8%) | 29.0% (9.8%) | 65.0% (11.3%) | 47.9% (9.4%) | -0.06 (0.18) | 204/144 |
| | Logistic Regression | 46.5% (4.6%) | 50.3% (6.8%) | 24.4% (14.1%) | 68.6% (19.2%) | 48.6% (4.1%) | -0.07 (0.1) | 204/144 |
| | Gaussian Naive Bayes | 52.7% (8.1%) | 54.3% (8.0%) | 43.0% (13.4%) | 62.4% (11.6%) | 52.2% (9.1%) | 0.06 (0.17) | 204/144 |
| | Random Forest | 51.5% (4.9%) | 58.4% (5.6%) | 11.9% (7.6%) | 91.2% (7.6%) | 46.7% (8.1%) | 0.06 (0.17) | 204/144 |
| | Support Vector Machine | 52.0% (6.1%) | 54.3% (7.1%) | 39.0% (7.0%) | 65.1% (13.0%) | 52.0% (6.1%) | 0.05 (0.13) | 204/144 |

| | | | | | | | | |
|---|---|---|---|---|---|---|---|---|
| DTI MD | Boosting Classifier | 52.6% (6.5%) | 55.2% (6.4%) | 37.4% (11.6%) | 67.8% (11.0%) | 51.7% (9.6%) | 0.05 (0.14) | 204/144 |
| | k-Nearest Neighbours | 50.0% (6.6%) | 54.3% (6.7%) | 25.5% (13.0%) | 74.5% (11.8%) | 49.1% (10.4%) | -0.0 (0.15) | 204/144 |
| | Logistic Regression | 47.5% (4.3%) | 50.3% (4.0%) | 31.3% (14.4%) | 63.8% (11.4%) | 45.6% (7.6%) | -0.06 (0.1) | 204/144 |
| | Gaussian Naive Bayes | 52.6% (4.6%) | 54.0% (5.0%) | 44.0% (9.6%) | 61.3% (9.4%) | 53.9% (5.3%) | 0.05 (0.1) | 204/144 |
| | Random Forest | 48.0% (5.8%) | 54.6% (6.6%) | 10.3% (8.0%) | 85.8% (11.7%) | 50.2% (12.5%) | -0.05 (0.18) | 204/144 |
| | Support Vector Machine | 46.8% (7.2%) | 49.1% (7.2%) | 33.3% (15.7%) | 60.3% (14.0%) | 46.8% (7.2%) | -0.07 (0.15) | 204/144 |
| DTI Network Parameters | Boosting Classifier | 51.5% (10.6%) | 53.4% (10.3%) | 40.8% (15.4%) | 62.2% (11.5%) | 51.8% (12.1%) | 0.03 (0.21) | 204/144 |
| | k-Nearest Neighbours | 49.5% (7.2%) | 53.8% (7.8%) | 24.4% (14.2%) | 74.5% (15.3%) | 47.2% (8.0%) | -0.01 (0.18) | 204/144 |
| | Logistic Regression | 47.6% (4.7%) | 54.3% (6.7%) | 7.8% (12.4%) | 87.4% (18.2%) | 43.7% (10.2%) | -0.05 (0.11) | 204/144 |
| | Gaussian Naive Bayes | 48.2% (8.3%) | 52.3% (8.8%) | 25.4% (15.2%) | 71.0% (16.5%) | 44.8% (11.5%) | -0.03 (0.18) | 204/144 |
| | Random Forest | 50.2% (5.7%) | 55.5% (5.2%) | 19.4% (14.3%) | 81.0% (10.0%) | 45.3% (7.8%) | 0.01 (0.14) | 204/144 |
| | Support Vector Machine | 46.5% (7.5%) | 49.4% (8.1%) | 30.3% (17.1%) | 62.7% (17.7%) | 46.5% (7.5%) | -0.07 (0.15) | 204/144 |

Note: HC = Healthy Controls, MDD = Major Depressive Disorder, BACC = Balanced Accuracy, ACC = Accuracy, AUC = Area Under The Receiver Operating Characteristic Curve, MCC = Matthew's Correlation Coefficient.

**eTable 18. Classification accuracy based on functional MRI for HC vs MDD (age 24 - 28).**

| *eTable 18. Classification accuracy for multivariate biomarkers based on functional MRI for HC vs MDD (age 24 - 28).* | | | | | | | | |
|---|---|---|---|---|---|---|---|---|
| **Modality** | **Algorithm** | **BACC** | **ACC** | **Sensitivity** | **Specificity** | **AUC** | **MCC** | **n (HC/MDD)** |
| Face Matching Task | Boosting Classifier | 49.1% (10.9%) | 50.0% (10.7%) | 40.2% (17.5%) | 58.1% (13.2%) | 47.2% (11.6%) | -0.02 (0.22) | 148/116 |
| | k-Nearest Neighbours | 50.3% (7.8%) | 53.1% (7.6%) | 28.4% (15.6%) | 72.3% (11.5%) | 49.5% (7.1%) | -0.0 (0.18) | 148/116 |
| | Logistic Regression | 46.2% (12.0%) | 48.1% (12.5%) | 31.3% (10.1%) | 61.2% (17.7%) | 46.0% (12.0%) | -0.08 (0.24) | 148/116 |
| | Gaussian Naive Bayes | 52.5% (6.6%) | 52.7% (7.0%) | 49.3% (12.2%) | 55.7% (15.7%) | 52.4% (6.5%) | 0.05 (0.14) | 148/116 |
| | Random Forest | 51.6% (4.7%) | 54.6% (4.7%) | 25.3% (11.0%) | 77.9% (9.8%) | 52.0% (10.6%) | 0.04 (0.11) | 148/116 |
| | Support Vector Machine | 48.5% (8.0%) | 50.0% (9.5%) | 36.4% (31.4%) | 60.6% (32.8%) | 48.5% (8.0%) | -0.01 (0.19) | 148/116 |
| RS Connectivity | Boosting Classifier | 57.4% (7.1%) | 58.7% (6.7%) | 45.3% (14.5%) | 69.6% (8.2%) | 59.7% (9.0%) | 0.15 (0.15) | 155/126 |
| | k-Nearest Neighbours | 56.3% (7.0%) | 58.0% (7.1%) | 38.9% (13.0%) | 73.7% (15.8%) | 60.2% (9.6%) | 0.15 (0.16) | 155/126 |
| | Logistic Regression | 58.8% (12.3%) | 59.8% (12.3%) | 49.2% (20.2%) | 68.5% (19.5%) | 60.0% (16.8%) | 0.19 (0.25) | 155/126 |
| | Gaussian Naive Bayes | 56.8% (12.2%) | 57.6% (12.2%) | 49.8% (12.6%) | 63.9% (14.1%) | 59.0% (12.6%) | 0.14 (0.25) | 155/126 |
| | Random Forest | 58.5% (6.2%) | 60.5% (6.3%) | 38.9% (8.9%) | 78.0% (8.3%) | 58.2% (10.3%) | 0.19 (0.13) | 155/126 |
| | Support Vector Machine | 54.5% (7.6%) | 55.1% (8.1%) | 47.1% (13.8%) | 62.0% (16.8%) | 54.5% (7.6%) | 0.1 (0.16) | 155/126 |
| ALFF | Boosting Classifier | 50.5% (9.9%) | 51.6% (9.4%) | 39.7% (16.0%) | 61.2% (8.2%) | 46.4% (12.7%) | 0.01 (0.2) | 155/126 |
| | k-Nearest Neighbours | 52.4% (9.9%) | 54.1% (9.7%) | 38.5% (16.7%) | 66.3% (14.5%) | 52.5% (11.8%) | 0.05 (0.21) | 155/126 |
| | Logistic Regression | 53.3% (9.4%) | 54.1% (9.0%) | 45.1% (13.7%) | 61.5% (10.2%) | 56.9% (11.7%) | 0.07 (0.19) | 155/126 |
| | Gaussian Naive Bayes | 48.4% (12.8%) | 48.4% (13.1%) | 49.7% (18.2%) | 47.1% (20.6%) | 49.1% (11.8%) | -0.04 (0.27) | 155/126 |
| | Random Forest | 51.3% (7.7%) | 54.1% (8.0%) | 25.3% (10.9%) | 77.3% (11.3%) | 53.9% (11.8%) | 0.03 (0.19) | 155/126 |
| | Support Vector Machine | 49.2% (7.5%) | 50.9% (7.1%) | 33.3% (11.4%) | 65.2% (8.0%) | 49.2% (7.5%) | -0.02 (0.16) | 155/126 |
| fALFF | Boosting Classifier | 50.6% (11.0%) | 50.9% (10.5%) | 48.2% (18.4%) | 52.9% (11.6%) | 52.5% (13.3%) | 0.01 (0.23) | 155/126 |

| | | | | | | | | |
|---|---|---|---|---|---|---|---|---|
| | k-Nearest Neighbours | 47.7% (5.8%) | 50.2% (6.5%) | 23.7% (9.4%) | 71.7% (15.7%) | 45.5% (8.7%) | -0.04 (0.16) | 155/126 |
| | Logistic Regression | 51.1% (7.6%) | 51.9% (8.1%) | 43.5% (15.3%) | 58.7% (20.6%) | 50.5% (9.7%) | 0.03 (0.17) | 155/126 |
| | Gaussian Naive Bayes | 52.9% (7.0%) | 53.4% (7.9%) | 50.5% (13.7%) | 55.2% (21.6%) | 52.0% (9.7%) | 0.05 (0.16) | 155/126 |
| | Random Forest | 51.3% (7.0%) | 54.1% (7.1%) | 24.6% (12.1%) | 78.1% (14.3%) | 56.5% (8.1%) | 0.03 (0.18) | 155/126 |
| | Support Vector Machine | 49.0% (6.3%) | 51.6% (7.0%) | 24.6% (14.5%) | 73.4% (18.4%) | 49.0% (6.3%) | -0.02 (0.16) | 155/126 |
| LCOR | Boosting Classifier | 52.9% (12.2%) | 54.1% (12.1%) | 42.1% (19.1%) | 63.7% (16.3%) | 53.8% (13.9%) | 0.06 (0.26) | 155/126 |
| | k-Nearest Neighbours | 54.5% (6.9%) | 55.8% (6.7%) | 41.9% (11.7%) | 67.0% (9.6%) | 55.8% (9.2%) | 0.09 (0.14) | 155/126 |
| | Logistic Regression | 51.9% (11.2%) | 53.3% (11.6%) | 38.1% (9.6%) | 65.6% (14.8%) | 49.9% (11.3%) | 0.04 (0.23) | 155/126 |
| | Gaussian Naive Bayes | 52.7% (4.6%) | 53.0% (4.8%) | 52.1% (15.7%) | 53.4% (15.0%) | 52.0% (5.1%) | 0.06 (0.1) | 155/126 |
| | Random Forest | 53.6% (10.1%) | 55.5% (10.1%) | 35.6% (13.0%) | 71.6% (12.7%) | 53.6% (9.5%) | 0.08 (0.21) | 155/126 |
| | Support Vector Machine | 49.9% (7.7%) | 51.6% (8.3%) | 33.3% (4.9%) | 66.4% (14.4%) | 49.9% (7.7%) | 0.0 (0.16) | 155/126 |
| RS Network Parameters | Boosting Classifier | 59.2% (10.7%) | 59.8% (10.5%) | 52.5% (16.0%) | 65.9% (13.4%) | 61.3% (14.0%) | 0.19 (0.22) | 155/126 |
| | k-Nearest Neighbours | 53.0% (5.1%) | 54.8% (5.3%) | 35.0% (12.0%) | 71.0% (12.1%) | 51.4% (5.1%) | 0.07 (0.11) | 155/126 |
| | Logistic Regression | 52.4% (11.2%) | 53.7% (11.4%) | 41.0% (11.9%) | 63.7% (16.5%) | 54.7% (13.0%) | 0.05 (0.24) | 155/126 |
| | Gaussian Naive Bayes | 53.6% (6.7%) | 54.1% (6.8%) | 48.5% (12.1%) | 58.7% (13.6%) | 53.4% (9.1%) | 0.08 (0.14) | 155/126 |
| | Random Forest | 62.4% (9.4%) | 64.1% (9.5%) | 45.4% (12.1%) | 79.3% (9.6%) | 61.3% (12.8%) | 0.27 (0.21) | 155/126 |
| | Support Vector Machine | 58.5% (9.9%) | 60.1% (9.8%) | 43.5% (12.2%) | 73.5% (12.4%) | 58.5% (9.9%) | 0.18 (0.21) | 155/126 |

Note: HC = Healthy Controls, MDD = Major Depressive Disorder, BACC = Balanced Accuracy, ACC = Accuracy, AUC = Area Under The Receiver Operating Characteristic Curve, MCC = Matthew’s Correlation Coefficient.

**eTable 19. Classification accuracy using neuroimaging modality integration for HC vs MDD (age 24 - 28).**

***eTable 19. Classification accuracy for multivariate biomarkers using neuroimaging modality integration for HC vs MDD (age 24 - 28).***

| Modality Integration | Modality | Algorithm | BACC | ACC | Sensitivity | Specificity | AUC | MCC | n (HC/MDD) |
|---|---|---|---|---|---|---|---|---|---|
| PCA-based | all | Boosting Classifier | 53.6% (10.6%) | 55.4% (11.3%) | 42.6% (20.3%) | 64.7% (23.0%) | 50.6% (12.1%) | 0.08 (0.24) | 133/94 |
| | | k-Nearest Neighbours | 48.2% (10.5%) | 52.9% (10.6%) | 22.9% (16.3%) | 73.5% (15.0%) | 54.7% (8.5%) | -0.04 (0.26) | 133/94 |
| | | Logistic Regression | 48.6% (7.5%) | 48.6% (12.1%) | 50.3% (33.0%) | 46.9% (40.9%) | 41.9% (13.9%) | -0.04 (0.21) | 133/94 |
| | | Gaussian Naive Bayes | 46.9% (10.4%) | 49.7% (9.9%) | 29.4% (24.1%) | 64.3% (19.1%) | 48.2% (17.2%) | -0.08 (0.23) | 133/94 |
| | | Random Forest | 53.2% (9.2%) | 58.1% (8.5%) | 26.1% (23.7%) | 80.2% (17.2%) | 55.8% (12.7%) | 0.06 (0.24) | 133/94 |
| | | Support Vector Machine | 52.0% (3.3%) | 53.7% (7.2%) | 43.1% (49.1%) | 60.8% (46.0%) | 52.0% (3.3%) | 0.08 (0.14) | 133/94 |
| Voting | all | all | 49.9% (8.4%) | 56.3% (8.9%) | 12.7% (9.9%) | 87.1% (12.5%) | 56.5% (14.8%) | 0.03 (0.23) | 133/94 |
| Voting | all | Boosting Classifier | 55.1% (10.2%) | 58.5% (8.3%) | 34.9% (23.6%) | 75.2% (7.7%) | 58.3% (11.5%) | 0.09 (0.23) | 133/94 |
| | | k-Nearest Neighbours | 49.9% (5.6%) | 56.8% (5.6%) | 10.4% (9.7%) | 89.5% (8.0%) | 52.8% (13.1%) | -0.0 (0.18) | 133/94 |
| | | Logistic Regression | 50.6% (8.5%) | 54.6% (8.7%) | 26.8% (15.2%) | 74.3% (15.2%) | 50.4% (12.9%) | 0.01 (0.19) | 133/94 |
| | | Gaussian Naive Bayes | 54.1% (10.3%) | 56.7% (9.8%) | 38.3% (17.8%) | 69.8% (13.1%) | 56.6% (11.8%) | 0.09 (0.21) | 133/94 |
| | | Random Forest | 50.8% (3.4%) | 58.2% (3.5%) | 7.6% (7.4%) | 94.0% (5.9%) | 59.5% (15.8%) | 0.03 (0.15) | 133/94 |
| | | Support Vector Machine | 48.7% (9.4%) | 52.8% (8.3%) | 24.4% (16.4%) | 73.0% (8.7%) | 49.0% (14.8%) | -0.03 (0.22) | 133/94 |
| Voting | Freesurfer | all | 53.5% (7.9%) | 59.0% (7.5%) | 22.1% (12.8%) | 84.9% (7.1%) | 51.1% (14.7%) | 0.09 (0.2) | 133/94 |
| | VBM | | 48.3% (8.1%) | 53.4% (8.9%) | 19.3% (8.6%) | 77.4% (10.3%) | 48.8% (9.3%) | -0.04 (0.19) | 133/94 |
| | DTI FA | | 48.8% (9.6%) | 54.5% (9.4%) | 14.8% (13.0%) | 82.8% (13.1%) | 47.2% (15.6%) | -0.04 (0.24) | 133/94 |
| | DTI MD | | 47.7% (8.9%) | 52.8% (8.6%) | 17.9% (11.2%) | 77.4% (12.5%) | 48.2% (10.4%) | -0.05 (0.24) | 133/94 |

| | | | | | | | | | |
|---|---|---|---|---|---|---|---|---|---|
| | DTI Network Parameters | | 48.9% (6.7%) | 56.3% (6.7%) | 5.2% (7.6%) | 92.5% (10.2%) | 48.8% (12.6%) | -0.02 (0.21) | 133/94 |
| | Face Matching Task | | 51.1% (10.8%) | 56.8% (10.0%) | 18.0% (17.6%) | 84.2% (5.7%) | 50.2% (13.5%) | 0.0 (0.29) | 133/94 |
| | RS Connectivity | | 53.3% (6.7%) | 56.8% (6.4%) | 33.9% (14.7%) | 72.8% (12.7%) | 59.1% (9.3%) | 0.07 (0.17) | 133/94 |
| | ALFF | | 51.6% (9.7%) | 54.6% (9.7%) | 35.0% (17.0%) | 68.1% (13.0%) | 51.2% (10.1%) | 0.04 (0.21) | 133/94 |
| | fALFF | | 50.9% (9.5%) | 54.6% (9.0%) | 29.0% (16.6%) | 72.9% (9.9%) | 51.9% (13.7%) | 0.01 (0.21) | 133/94 |
| | LCOR | | 53.1% (9.7%) | 56.0% (8.5%) | 36.2% (18.1%) | 70.0% (6.7%) | 53.8% (11.3%) | 0.06 (0.21) | 133/94 |
| | RS Network Parameters | | 56.9% (10.9%) | 60.3% (9.6%) | 36.2% (19.7%) | 77.5% (8.3%) | 60.4% (13.2%) | 0.14 (0.23) | 133/94 |

Note: HC = Healthy Controls, MDD = Major Depressive Disorder, BACC = Balanced Accuracy, ACC = Accuracy, AUC = Area Under The Receiver Operating Characteristic Curve, MCC = Matthew's Correlation Coefficient.

19 Consortium E. Structural image processing protocols. http://enigma.ini.usc.edu/protocols/imaging-protocols/.

20 Whitfield-Gabrieli S, Nieto-Castanon A. Conn: A Functional Connectivity Toolbox for Correlated and Anticorrelated Brain Networks. *Brain Connectivity* **2**012; 2: 125–41.

21 Muschelli J, Nebel MB, Caffo BS, Barber AD, Pekar JJ, Mostofsky SH. Reduction of motion-related artifacts in resting state fMRI using aCompCor. Neuroimage 2014; **96**: 22–35.

22 Yang H, Long X-Y, Yang Y, et al. Amplitude of low frequency fluctuation within visual areas revealed by resting-state functional MRI. Neuroimage 2007; **36**: 144–**52**.

23 Deshpande G, LaConte S, Peltier S, Hu X. Integrated local correlation: A new measure of local coherence in fMRI data. *Hum Brain Mapp* 2009; **30**: 13–23.

24 Hariri AR, Tessitore A, Mattay VS, Fera F, Weinberger DR. The Amygdala Response to Emotional Stimuli: A Comparison of Faces and Scenes. Neuroimage 2002; 17: 317–23.

25 Kessler R, Schmitt S, Sauder T, et al. Long-Term Neuroanatomical Consequences of Childhood Maltreatment: Reduced Amygdala Inhibition by Medial Prefrontal Cortex. *Frontiers Syst Neurosci* 2020; **14**: 28.

26 Repple J, Mauritz M, Meinert S, et al. Severity of current depression and remission status are associated with

structural connectome alterations in major depressive disorder. *Mol Psychiatr* 2020; **25**: 1550–8.
27 Heuvel MP van den, Sporns O, Collin G, et al. Abnormal Rich Club Organization and Functional Brain Dynamics in Schizophrenia. Jama Psychiat 2013; **70**: 783–92.
28 Andersson JLR, Skare S. A Model-Based Method for Retrospective Correction of Geometric Distortions in Diffusion-Weighted EPI. Neuroimage 2002; **16**: 177–99.
29 Chang L, Walker L, Pierpaoli C. Informed RESTORE: A method for robust estimation of diffusion tensor from low redundancy datasets in the presence of physiological noise artifacts. *Magnet Reson Med* 2012; **68**: 1654–63.
30 Chang L, Jones DK, Pierpaoli C. RESTORE: Robust estimation of tensors by outlier rejection. Magnet Reson Med 2005; **53**: 1088–95.
31 Lange SC de, Heuvel MP van den. Structural and functional connectivity reconstruction with CATO - A Connectivity Analysis TOolbox. *Biorxiv* 2021; : 2021.05.31.446012.
32 Cammoun L, Gigandet X, Meskaldji D, et al. Mapping the human connectome at multiple scales with diffusion spectrum MRI. *J Neurosci Meth* 2012; **203**: 386–97.
33 Hagmann P, Cammoun L, Gigandet X, et al. Mapping the Structural Core of Human Cerebral Cortex. *Plos Biol* 2008; **6**: e159.
34 Lange SC de, Scholtens LH, Initiative ADN, et al. Shared vulnerability for connectome alterations across psychiatric and neurological brain disorders. *Nat Hum Behav* 2019; **3**: 988–98.
35 Rubinov M, Sporns O. Complex network measures of brain connectivity: Uses and interpretations. Neuroimage 2010; 52: 1059–69.
36 Chicco D, Tötsch N, Jurman G. The Matthews correlation coefficient (MCC) is more reliable than balanced accuracy, bookmaker informedness, and markedness in two-class confusion matrix evaluation. *Biodata Min* 2021; **14**: 13.
